# Long-read genome sequencing and multi-omics in aging and neurodegeneration

**DOI:** 10.1101/2025.10.10.25337775

**Authors:** Tanner D Jensen, Yann Le Guen, Lia Talozzi, Sherry Yang, John Gorzynski, Andrés Peña-Tauber, Ilaria Stewart, Alexis Ferrasse, Daniel Nachun, Maggie T Arriaga, Justin Lee, Rafael Catoia Pulgrossi, Junyoung Park, Jingyu Zhang, Anthony D. Wagner, Elizabeth C. Mormino, Kathleen L. Poston, Victor W. Henderson, Zihuai He, Tony Wyss-Coray, Stephen B Montgomery, Euan A Ashley, Michael D. Greicius

## Abstract

Structural variants (SVs) are a major source of genetic variation yet remain underexplored in healthy aging and neurodegenerative diseases. We performed nanopore long-read genome sequencing (lrGS) on 551 deeply-phenotyped individuals from Stanford’s Aging and Memory Study and Alzheimer’s Disease Research Center, generating a comprehensive SV map integrated with matched methylation, transcriptomic, and proteomic data. Over 60% of SVs identified by lrGS were not detected with short-read WGS, including many poorly tagged by single-nucleotide variants (SNVs). We discovered >60,000 SV-QTLs across molecular traits and showed that SVs were more likely than SNVs to be fine-mapped as causal. Colocalization with Alzheimer’s and Parkinson’s disease GWAS implicated SVs at multiple loci, including *TMEM106B*, *BIN3*, and *NBEAL1*. Multi-omic outlier enrichment and Bayesian modeling prioritized rare functional SVs near known risk genes. Combined, these data reveal widespread regulatory SVs in healthy aging and neurodegeneration, underscoring the importance of lrGS in deciphering complex genetic architecture.

## Introduction

Structural variants (SVs), defined as genomic alterations ≥50 base pairs including deletions, insertions, inversions, and complex rearrangements, represent a major source of genetic diversity and disease risk. SVs span larger genomic segments than single-nucleotide variants (SNVs) and are disproportionately enriched for functional effects on gene regulation, dosage, and chromatin structure (Collins et al. 2020; Eichler 2019). Yet much of their contribution to human disease remains uncharacterized, largely due to historical limitations in their detection by short-read sequencing as previously reported (Chaisson et al. 2019; Ebert et al. 2021).

Neurodegenerative disorders, such as Alzheimer’s disease (AD) and Parkinson’s disease (PD), exemplify these challenges. Recent genome-wide association studies (GWAS) have greatly expanded the list of loci associated with risk: the largest AD meta-analysis to date identified 75 loci implicating endocytic and immune pathways (Bellenguez et al. 2022), while a recent multi-ancestry meta-analysis of PD revealed 90 independent risk variants across diverse populations (Kim et al. 2024). A limited number of studies have examined SV associations with PD (Billingsley et al. 2023), AD (Vialle et al. 2025; Wang et al. 2025), and related dementias (Xu et al. 2024) using short-read whole-genome sequencing (srGS). In AD, one of the strongest examples of an SV tagged by the lead GWAS single-nucleotide variant (SNV) is a 316 bp Alu insertion in the 3’ UTR of *TMEM106B* (Chemparathy et al. 2024; Vialle et al. 2025; Bellenguez et al. 2022). Despite these initial insights, the degree to which SVs contribute to GWAS associations remains unknown.

Long-read genome sequencing (lrGS) is beginning to provide enhanced understanding of the contribution of SVs to human diseases. Recent projects, including the Human Genome Structural Variation Consortium (HGSVC) (Ebert et al. 2021, Logsdon et al. 2025), the Telemore-to-telomere and Human Pangenome Reference Consortium (HPRC) (Nurk et al. 2022, Liao et al 2025), long-read sequencing of 1000 Genomes Project samples (Gustafson et al. 2024, Schloissnig et al. 2025), and All of Us (Mahmoud et al. 2024), have shown that short-read pipelines miss more than half of SVs, especially in GC-rich, duplicated, or complex loci. These efforts have begun to reveal the spectrum of SV diversity and demonstrate the relevance of SVs detected with lrGS to both population genomics and disease biology (Logsdon, Vollger, and Eichler 2020).

Here, we present a population-scale long-read sequencing and multi-omics dataset from aged individuals with and without neurodegenerative disease. By integrating structural variant calls with methylation, single-cell transcriptomics, and proteomics, we uncover SVs that influence molecular traits and underlie unresolved GWAS signals in aging and neurodegeneration.

## Results

### Long-read sequencing of a deeply phenotyped neurodegenerative disease cohort shows SVs at GWAS loci

We performed nanopore-based lrGS on 551 individuals recruited as part of the Stanford Aging and Memory Study (SAMS only; n=125), the Stanford Alzheimer’s Disease Research Center (ADRC) (n=391), or co-enrolled in both (n=35). Participants were characterized as cognitively normal seniors or grouped based on clinical neurodegenerative disease diagnoses (mild cognitive impairment MCI, Alzheimer’s disease, Parkinson’s-MCI, Parkinson’s disease, Lewy body dementia) supported by biomarker data. Participants spanned an age range of 36-88 years old at recruitment (mean = 70.7; std = 8.0), with 261 males and 290 females (**Figure 1a, Extended Data Table 1**). Participants underwent extensive cognitive assessments, with multiple biomarker data types collected from plasma, cerebrospinal fluid (CSF), brain MRI, and amyloid PET to help support diagnoses. A subset of participants also had short-read whole genome (srGS) using MGI-sequencing (n=366), as well as SomaLogic proteomics from plasma (n=495) and/or CSF (n=226), and single-cell RNA sequencing from peripheral blood mononuclear cells (PBMCs) (n=275). For our study, DNA extracted from PBMCs was sequenced on the Oxford Nanopore PromethION to generate lrGS data, achieving a median coverage of 16×, read-length N50 of 18 kb, and median Quality score of 12.3 (corresponding to roughly 93.6% identity) after sample quality control (**Figure 1b, Supplementary figure 1**). Structural variants (SVs) and methylation were subsequently called from the long-read genome data (**Methods**). Population stratification analysis of common SVs (MAF > 2%) showed expected clustering by ancestry group as self-reported by participants (**Extended Data Figure 1**). Many SVs overlapped 85 known Alzheimer’s (Bellenguez et al. 2022) and 102 known Parkinson’s GWAS loci (Nalls et al. 2019; Kim et al. 2024). While a higher burden of SVs was observed within 100Kb of AD loci (mean = 19.5) relative to a large-scale control GWAS for BMI (Huang et al. 2022; mean = 14.1; p = 0.025), the overall pattern of SV burden near GWAS loci was broadly similar (**Figure 1c**).

**Figure 1.**
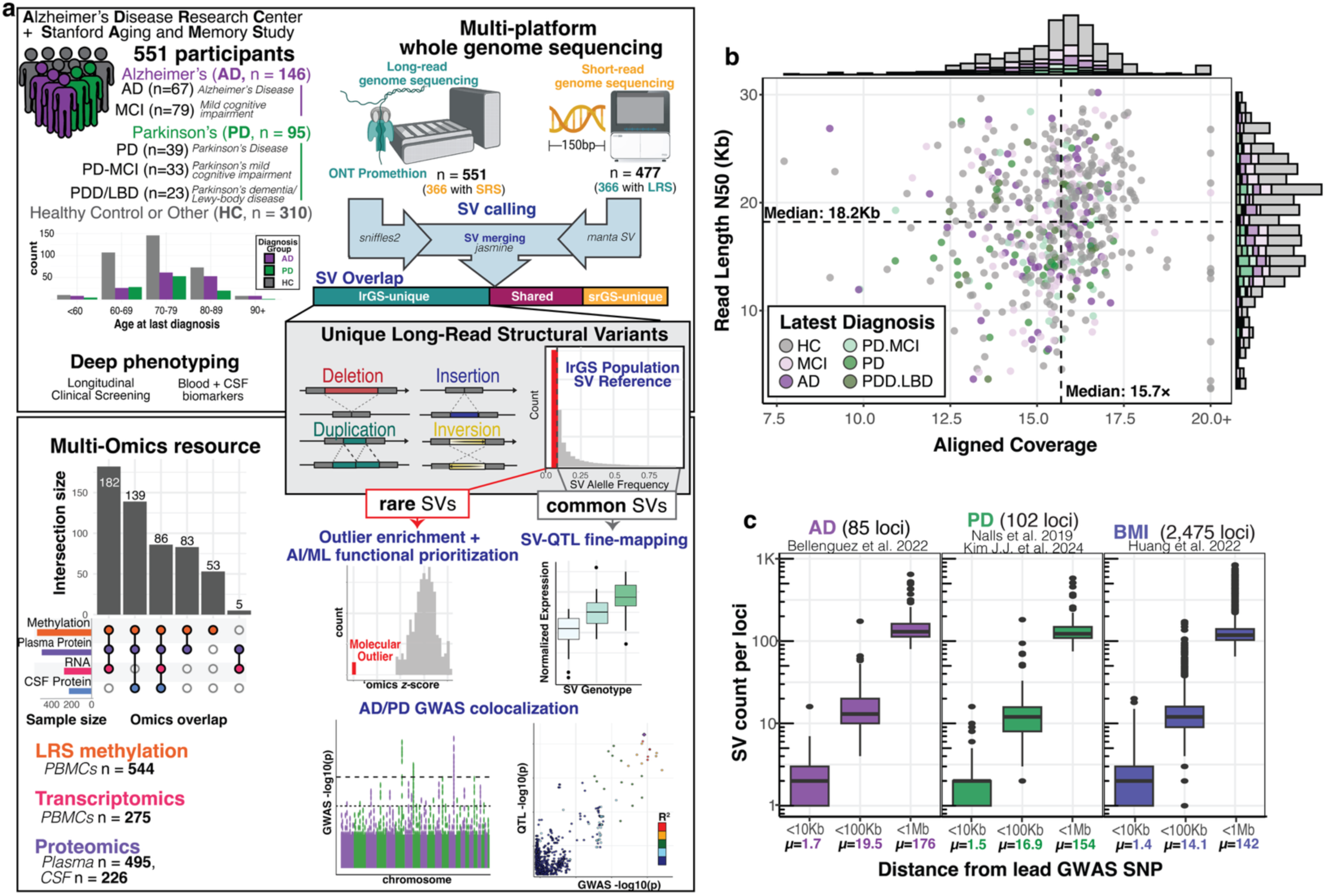
Long-read sequencing and multi-omics in healthy aging and neurodegenerative disease. **a**, Illustrative outline of paper analyses. Data from aging individuals from the Stanford Alzheimer’s Disease Research Center (ADRC) and the Stanford Aging and Memory Study (SAMS) were analyzed using long-read sequencing. ADRC and SAMS cohorts contain a wealth of deep phenotype data, including initial neuropsychological assessment and, for ADRC, yearly clinical visits, along with MRI and PET scans, proteomics from plasma and CSF, and transcriptomics from PBMCs. Long-read sequencing revealed structural variants (SVs) (many not detected with short-read sequencing) that we interpreted using matched multi-omics data. Functional rare SVs were prioritized with WatershedSV, and functional common SVs were identified through QTL fine-mapping. Colocalizing QTL associations with Alzheimer’s and Parkinson’s disease GWAS reveals functional SVs that are likely causal at neurodegeneration risk loci. **b**, Sequencing summary metrics of 551 long-read genomes sequenced on R9.4 flowcells on the PromethION device. Colored by diagnosis group, HC=healthy control, AD=Alzheimer’s disease, MCI=mild cognitive impairment, LBD=Lewy body dementia, PD=Parkinson’s disease. **c**, Burden of SVs from Sniffles2 joint genotyping near GWAS lead SNVs at various distance thresholds for 3 different GWAS phenotypes. For PD, SNV-union between Nalls et al. and Kim JJ et al was used. BMI=body mass index.

### Most SVs discovered by long reads remain undetected by conventional short-read pipelines

Among the 366 individuals with both srGS and lrGS data, we observed that the majority of lrGS insertions (85%) were unique to long-read, while 70% of srGS insertions were detected in long-read (**Figure 2a**). For deletions, the difference was more modest; 66% of lrGS deletions were unique to long-read, while 59% of srGS deletions were unique (**Figure 2a**). While most SVs are insertions and deletions, we also detected duplications and inversions. For these variants, srGS identified higher counts, particularly for duplications under 1kb (**Extended Data Figure 1**). The increased calling of inversions and duplications in srGS may be related to the higher coverage srGS to identify read-depth changes and break points, false positives which are more prevalent in srGS, or the long-read call set splitting some duplications as insertions.

**Figure 2.**
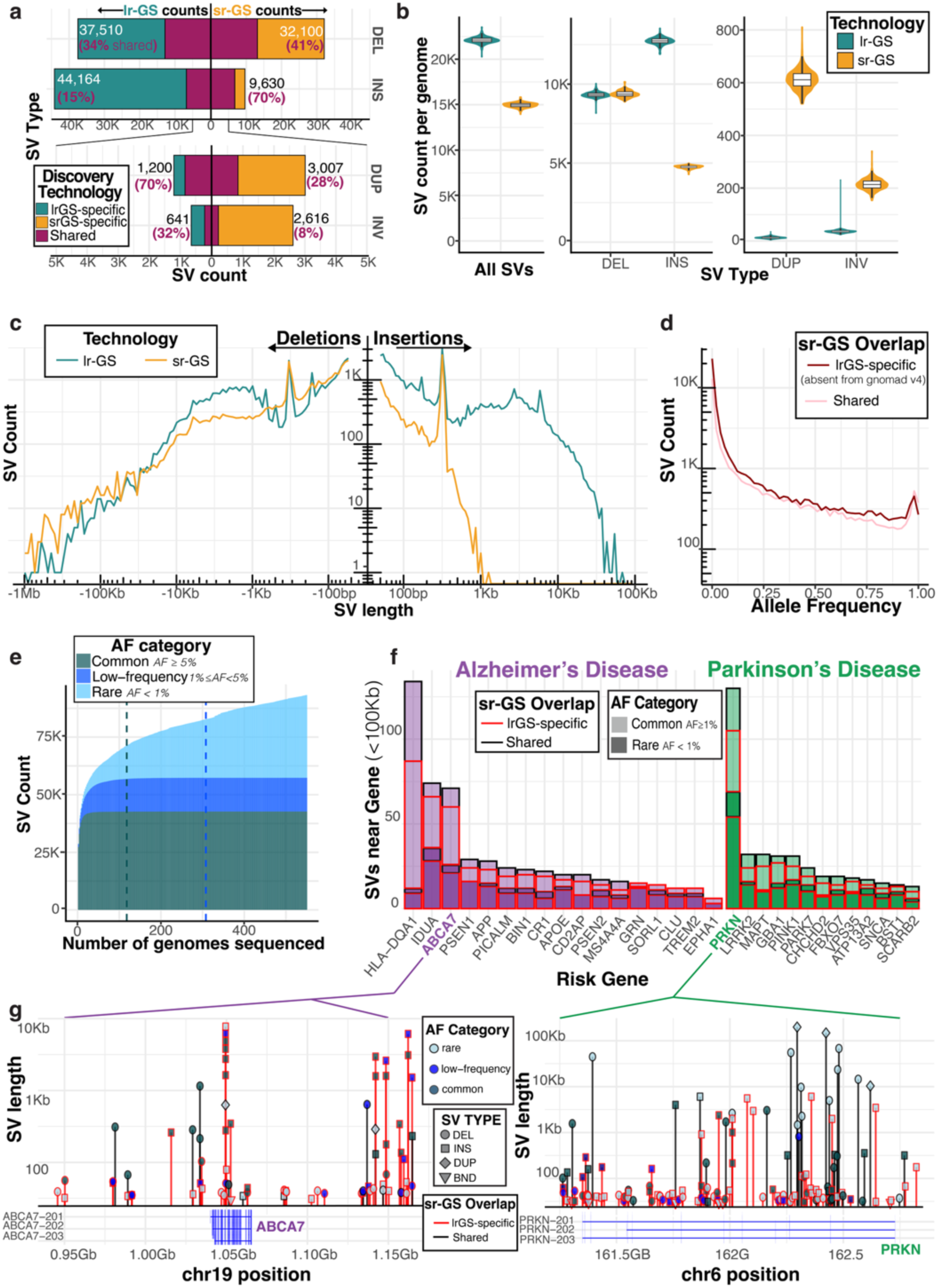
Long-read-specific structural variants across the frequency spectrum are found near neurodegeneration risk genes. **a,** In 366 matched individuals with both LRS and SRS genomes, count of structural variants identified by each technology after merging SVs with Jasmine. Percentages of shared SVs found in each technology that were also detected by the other technology are highlighted. **b,** Burden of SVs per genome comparing LRS Sniffles2 call set and SRS manta-paragraph call set. **c,** In the 366 matched individuals, count of deletions and insertions identified by each technology across variant length spectrum. **d,** Sniffles2 joint genotyping was used to ascertain allele frequencies for SVs. Counts of SVs shown across frequency spectrum, stratified by whether variant was specific to LRS (absent from short-read call set and additionally not found in SRS-based population resources gnomad v4), or also detected by SRS. **e,** Number of total SVs identified as a function of how many genomes were sequenced. Vertical lines show saturation point for common variants (AF ≥ 0.05) and for low-frequency variants (0.01 ≤ AF < 0.05). **f,** Count of SVs within 100Kb of a selection of well-known Alzheimer’s and Parkinson’s disease genes, stratified both by whether variant is rare or common and whether it was detected by short-read sequencing or not. **g,** Structural variant landscape in 100kb window around AD gene *ABCA7*, and PD gene *PRKN*. X-axis displays left break-point of SV and Y-axis displays variant length. Lollipops are shaped by SV type and colored by allele frequency, long-read-unique variants absent from SRS population references colored in red.

On average, lrGS yielded ∼22,400 SVs per genome, compared to ∼15,000 from srGS (**Figure 2b**). Individuals with non-European ancestry showed a higher median number of SVs per genome compared to European participants (**Extended Data Figure 1**). The length distribution of long-read SVs exhibited pronounced peaks at ∼300 bp, ∼2.5 kb, and ∼6 kb, corresponding to Alu, SVA, and LINE-1 mobile elements (Audano et al 2019), while short-read detection dropped sharply for insertions >1 kb (**Figure 2c**).

Upon intersecting our lrGS call set with short-read-derived SVs in gnomAD v4 (Collins et al 2020), we found that over 50% of lrGS SVs were absent from srGS population references; those SVs shared with reference datasets exhibited a similar frequency distribution but were primarily composed of deletions and short insertions (<1 kb). Among low-frequency and common SVs with minor allele frequency (MAF) >2%, we observed that short insertions and deletions (<150 bp) frequently exhibit low linkage disequilibrium (LD) (r² < 0.3) with nearby SNVs, indicating poor tagging. Mid-sized SVs (150–1000 bp), particularly insertions, exhibited stronger LD with adjacent SNVs (**Extended Data Figure 2**). Furthermore, SVs with higher overlap fractions (OFP) with gnomAD v4 short-read SVs demonstrated improved SNV tagging (SNVs with highest LD with SVs), especially among mid-sized insertions (**Extended Data Figure 3**).

To further characterize differences between shared variants and those detected uniquely by lrGS, we classified repeat context of each insertion and deletion sequence using RepeatMasker (Smits et al 2015). Consistent with prior observations, a majority of detected SVs represent repetitive sequence (**Supplementary figure 2c**). Compared to srGS-shared variants, lrGS-specific variants were more frequently located within tandem repeat and satellite regions, whereas canonical retrotransposon-derived events (e.g., LINEs and SINE/Alus), were more often shared between lrGS and srGS datasets (**Supplementary figure 2d**). To localize sources of long-read vs short-read discordance, we also stratified SVs using GIAB region annotations (segmental duplications, tandem repeats, sequence, low-mappability, and “not in all difficult regions” (ie., “easy”) sequence) (Dwarshuis et al 2024). LrGS–specific SVs were enriched in segmental duplications and other difficult regions, whereas SVs supported by both technologies were strongly enriched in GIAB “easy” regions (easy: 61.7% for shared vs 35.6% for lrGS–unique), consistent with known limitations of short-read SV discovery in repetitive/low-mappability sequence (**Supplementary figure 2a**).

To establish the biological validity of our lrGS–specific SVs, we queried ascertainment in orthogonal long-read reference databases. A majority of lrGS–unique SVs matched external long-read resources (55.7% matched HGSVC3 (65 samples) (Logsdon et al 2025); 61.1% matched a 1000 Genomes ONT dataset (500 samples) (Gustafson et al 2024)), with substantially higher concordance for more common frequency SVs (minor allele frequency (MAF) > 10%: 85.1–95.2%) than for rarer frequency SVs (MAF < 1%: 33.3–38.1%), as expected given reference incompleteness for rare variants (**Supplementary figure 2b**). These analyses clarify that many lrGS–unique SVs represent genuine variation missed by short reads.

### Population-scale long-read sequencing saturates the discovery of common SVs in European populations

Population sequencing of 551 nanopore genomes enabled estimation of structural variant allele frequency. We used *Sniffles2* to jointly genotype SVs across the cohort and calculate allele frequencies (AFs). Additionally, we queried existing srGS-based population references (i.e. gnomAD v4) to examine the extent of overlap with our lrGS calls (**Methods**). We define lrGS-specific variants as those absent from these srGS resources and not detected in our cohort’s matched short-read call set. We found 54% of common SVs (minor allele frequency ≥ 5%) were specific to lrGS; this increased to 64% for rare and low-frequency variants (MAF < 5%). Overall, 40% of SVs in our cohort were rare (MAF < 1%) (**Figure 2d**), demonstrating the necessity of population-scale lrGS sequencing to establish the AF of novel lrGS variants. We share allele frequencies and coordinates of the SVs we discovered as a resource to help identify and filter out SVs that are tolerated in healthy aging (**Table S1**, see **Data Availability**).

To determine if the size of our cohort was sufficiently large to detect most common SV alleles, we performed a down-sampling analysis. By calculating the number of SVs detected as a function of genomes sampled, we found that counts of common variants (AF ≥ 5%) saturated at 67 genomes. For low-frequency variants (1% ≤ AF < 5%), this saturation occurred after 272 genomes. As expected, counts of rare variants continued to increase with each genome sequenced at a rate of about 45 variants per genome (**Figure 2e**). Extrapolating from these saturation points, we expect we would need to sequence 12,000 genomes to achieve saturation of variants above an allele frequency of 0.1% (**Extended Data Figure 1e**). Together, these data indicate that with 551 genomes, we have achieved comprehensive coverage of common and low-frequency SV alleles in our mostly-European population sample.

Having discovered lrGS-specific variants across the frequency spectrum, we investigated whether these variants could be impacting risk genes for neurodegenerative disease. We intersected our SV calls with a selection of 30 well-known Alzheimer’s and Parkinson’s disease genes, and found these genes had a median of 20 SVs (range 6-134) in a 100Kb window; overall 46.4% were rare and 71% were novel to lrGS (**Figure 2f**). The structural variant landscape of two genes of interest, *ABCA7* for AD, which is known to harbor complex structural rearrangements, and *PRKN* for PD, a long gene with known deletion and duplication alleles, depicts the density of lrGS-discovered variants across both length and frequency spectrum, highlighting the discovery power of performing long-read sequencing to find novel variants near disease genes (**Figure 2g**).

### Integration of multi-omics and long-read sequencing reveals outliers enriched near rare SVs

To interpret the functional consequences of the rare variants we discovered with lrGS, we leveraged existing multi-omics data available for ADRC and SAMS participants. We identified molecular outliers from these data using covariate-adjusted z-scores, with normalization parameters selected to maximize cross-omic outlier concordance (Methods) (Ferraro et al. 2020; Li et al. 2023; Chui et al. 2025). We discovered 9,211 expression and 38,074 protein outliers (|*z*| > 2) across 4,659 genes and 6,004 proteins, respectively. Using lrGS, we called CpG methylation and used METAFORA to detect a total of 14,462 methylation outliers across 6,641 unique regions (**Extended Data Figures 4a**) (Jensen et al. 2026). Summarizing this outlier information, we found that a typical genome has a median burden of 79 genes with a nearby (<100Kb) rare SV, 25 genes with methylation outliers within 10Kb, 354 genes with RNA expression outliers, and 229 and 92 proteins with abundance outliers in plasma and CSF, respectively (**Figure 3a**).

**Figure 3.**
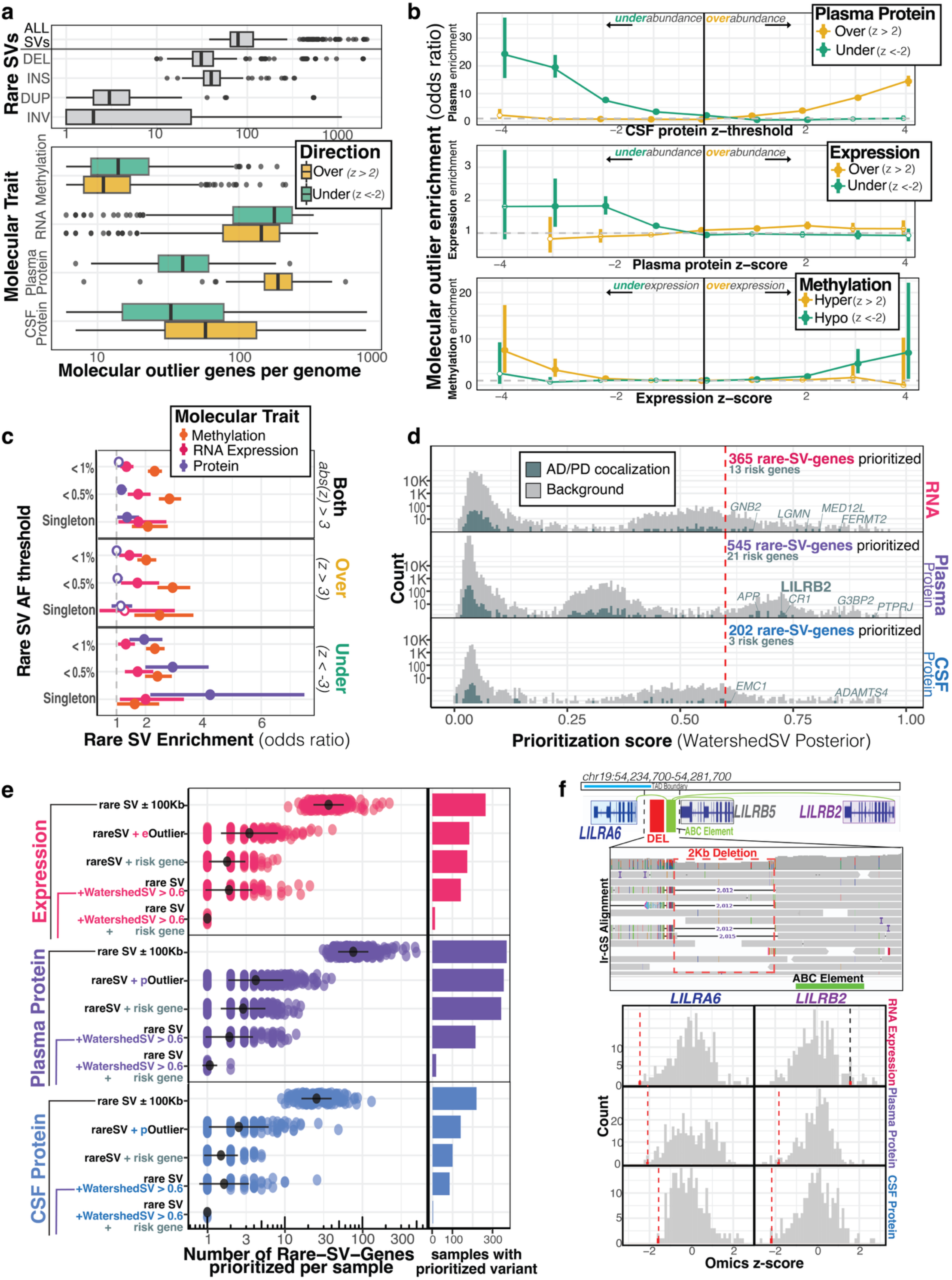
Molecular outlier prioritization of functional rare structural variants impacting neurodegenerative disease genes. **a,** Burden of molecular outliers per individual across molecular traits with |*z*| > 2. Genes defined as outlier for SV type if they have an SV within 100Kb and for methylation if they have a METAFORA methylation outliers within 10Kb. **b,** Pairwise multi-omic outlier enrichments showing the concordance of outliers (|z| > 2) across tissues and layers of regulation. X-axis depicts the z-score threshold to define outliers--negative values for under-outliers (z-score < -x), positive values for over-outliers (z-score > x). Y-axis displays Fisher’s exact test odds ratio of observing an outlier in a comparison ome (y-axis label) given an outlier was observed in a discovery ome (x-axis label). Solid points represent nominally significant enrichments, white-filled points are not significant. **c,** Enrichment from Fisher’s exact test of rare SVs within 10Kb of molecular outlier (|*z*| > 3) genes at various AF thresholds. Singleton SVs are variants only identified in a single genome. Y-axis stratified by outlier direction. “Both” represents outliers in either tail, while “under” and “over” represent outliers in negative and positive direction, respectively. White-filled points represent enrichments that were not nominally significant. **d,** Distribution of WatershedSV posteriors across RNA expression, plasma protein, and CSF protein. Log-scaled y-axis shows the histogram of rare-SV-genes scored across posterior threshold. Line at posterior = 0.6 represents the prioritization cut-off used to label functional rare variants. Risk genes based on colocalizations of AD and PD GWAS are highlighted. **e,** Prioritization scheme for identifying disease-relevant functional rare variants. Number of SV-gene pairs per genome are triaged down based on additional filters including, if a gene is an outlier abs(z-score)>2 (rare SV + outlier), whether a gene is in a risk gene list based on AD/PD GWAS colocalizations (rare SV + risk gene), whether a gene was highly prioritized with Watershed (rare SV + WatershedSV > 0.6), and a combination of both Watershed prioritization and colocalization evidence (rare SV + WatershedSV > 0.6 + risk gene). Bar plot shows the number of individuals with at least 1 rare-SV-Gene pair prioritized with a given filter. **f,** Example of a rare, non-coding deletion prioritized by WatershedSV. A rare 2 kb deletion in the LILRB gene cluster on chr19 was called in a single SAMS individual as seen in the IGV screenshot showing read alignment pile-up. The deletion was proximal to an ABC element linked to nearby *LILRA6* and *LILRB2* genes. *LILRB2* is a known Alzheimer’s risk gene based on GTEx eQTL colocalizations. Across multiple ‘omes, the deletion-carrier sample was an under-outlier in both LILRA6 and LILRB2 as seen in omics z-score histograms. Vertical line represents the deletion-carrier’s z-score, colored in red if the carrier is in the bottom 1% tail of the distribution.

Many of these molecular outliers were shared across various tissues and layers of regulation. We found that over-expression outlier genes were strongly enriched for having hypomethylation outliers (odds-ratio (OR)=5.21, p-value=9.9E-8); conversely, under-expression outliers were enriched for hypermethylation outliers (OR=2.82, p-value=1.1E-3), supporting the canonical model that promoter methylation is associated with silencing of genes. Comparing expression to protein levels, we observed enrichment of under-expression outliers with under-abundance outliers of plasma protein (OR=1.6, p-value=1.3E-8), and weaker enrichment for over-expression with protein over-abundance (OR=1.15, p-value=6.6E-3), indicating that under-expression was a stronger predictor of protein abundance than overexpression. Finally, we demonstrate replicability across two different tissues, CSF and plasma; CSF outliers were strongly enriched near plasma outliers in concordant directions (under-under: OR=20.3, p-value<1E-32; over-over: OR=8.7, p-value<1E-32) (**Figure 3b**).

We assessed if molecular outliers were enriched near rare SVs (<10Kb) as reported in prior SV studies (Chiang et al. 2017; Jensen et al. 2024). Here, we replicated the expression outlier enrichment of rare SVs in our ADRC cohort (|*z*| > 3, OR=1.34, p-value=7.9E-4) and further show rare SVs are also enriched near methylation and protein outliers (methylation OR=2.32, p-value<1E-32; plasma protein |*z*| > 3, OR=1.35, p-value=0.034). When stratifying enrichments by outlier direction, we observe that expression and methylation were enriched in both directions, whereas plasma protein was only enriched for under-abundance (*z* < -3, OR = 1.96, p-value = 1.92E-5), as was previously observed for the enrichment of rare SNVs (Li et al. 2023). We saw the strongest enrichments at lower allele frequency thresholds for rare SVs, particularly singleton variants, where rare SVs were 4x more likely to be seen near protein under-expression outliers compared to inliers (**Figure 3c**). Importantly, restricting this analysis to a high-confidence subset of rare SVs supported by orthogonal evidence recapitulated the same enrichment patterns across all molecular traits, albeit with modestly reduced effect sizes (**Supplementary figure 3**). When calculating type-specific enrichments for rare SVs, we observed that insertions were most enriched for driving hypermethylation outliers, and deletions were particularly enriched for under-abundance protein outliers (**Extended Data Figure 4b**). In general, we observed higher enrichments at higher absolute z-score thresholds (**Extended Data Figure 4c**) and for SVs overlapping the gene body. However, we detected long-range enrichment of rare SVs on methylation and expression as far out as 500Kb from the gene (**Extended Data Figure 5d**).

#### Outliers prioritize functional SVs near neurodegeneration risk genes

We applied WatershedSV, a hierarchical Bayesian model that integrates outlier omics signals with genomic annotations, to prioritize potentially functional rare SVs (Jensen et al. 2025). Variants were scored separately using three WatershedSV models for RNA expression, plasma protein levels, and CSF protein levels as each model prioritizes different annotations (**Supplementary Figures 4-6**) We identified 365, 545, and 202 prioritized rare SV-gene pairs from RNA expression, plasma protein levels, and CSF protein levels, respectively (Watershed posterior probability > 0.6) (**Table S2**, **Figure 3d**).

We next applied additional filters to the WatershedSV-prioritized variants to prioritize those most likely contributing to AD or PD risk. SVs were categorized based on outlier status of proximal genes, WatershedSV prioritization score, and whether gene had eQTL or pQTL colocalization evidence from AD or PD GWAS (coloc PP4 > 0.5). Among colocalized risk genes, we found 13, 21, and 3 prioritized rare SV-gene pairs for expression, plasma protein, and CSF protein, respectively (**Figure 3e**). Notably, one such prioritized rare SV near *LILRB2*— an Alzheimer’s risk gene identified through GTEx eQTL colocalizations—was detected in plasma protein data. This rare SV deletes a 2kb non-coding region of the *LILRB* gene cluster, overlapping a TAD boundary and adjacent to an ABC regulatory element linked to both *LILRA6* and *LILRB2*. The deletion was identified in a single individual from the SAMS cohort, and the deletion-carrier sample was a strong under-outlier in both *LILRA6* and *LILRB2* expression across multiple omics layers (**Figure 3f)**.

#### SV-QTL mapping across multi-omics discovers novel associations

To prioritize low-frequency and common structural variants (MAF > 2%) that may be functional, we used SV genotypes from lrGS and the matched omics data to map molecular QTLs. At an FDR threshold of 0.05, we discovered 60,177 SV-QTLs for 16,201 unique features (molQTL targets) over our 4 molecular traits (**Figure 4a, Extended Data Figure 6a, Table S3**) (methylation: 53,112 SV-QTLs in 13,397 regions; expression: 3,202 SV-QTLs in 1,216 genes; plasma: 2,904 SV-QTLs in 1,149 proteins; CSF: 959 SV-QTLs in 439 proteins). While most QTLs we found were for insertions and deletions, when a QTL was detected for an inversion or duplication, it exhibited larger effect sizes. A typical QTL-associated SV affected 1.51 genes for expression and protein, and 2.36 regions for methylation. Similarly, a typical molQTL target had 2.5 associated SVs across expression or protein, and an average of 4 SV-QTLs for methylation (**Extended Data Figure 6b**). By overlapping QTL-associated SVs across molecular traits, we discovered SVs with effects across multiple molecular traits, including over 100 SVs which had a significant QTL in all 4 traits. (**Extended Data Figure 6c**) 62.1% of our SV-QTLs included a lrGS-specific SV, particularly for insertions where a majority (76%) of QTLs were lrGS-specific (**Extended Data Figure 6d**). Given the sample size of our lrGS dataset, we were able to ascertain low-frequency variants and include them in QTL mapping. When stratifying SV-QTL effect size by MAF, we observed that low-frequency SVs (MAF 2-5%) had significantly higher effect sizes compared to more common SVs (beta=0.26, p-value<1E-32) (**Figure 4b**). In addition to allele frequency, we also observed that longer variants had larger absolute effect sizes (Length>10Kb, beta=0.11, p-value<1E-32) (**Extended Data Figure 6e**). Since SVs can have long-range effects, we tested all SVs within a 1Mb window of the TSS/methylation region and observed that 16% of SV-QTLs were within 10Kb and 58% were within 100Kb. QTLs for methylation had closer-range effects compared to the other omes (p-value=9.4E-18), and across all molecular traits, closer SVs had significantly higher effects (log-distance slope=-0.022, p-value<1E-32), particularly those for SVs directly overlapping their molecular target (**Extended Data Figure 6f,g**).

**Figure 4.**
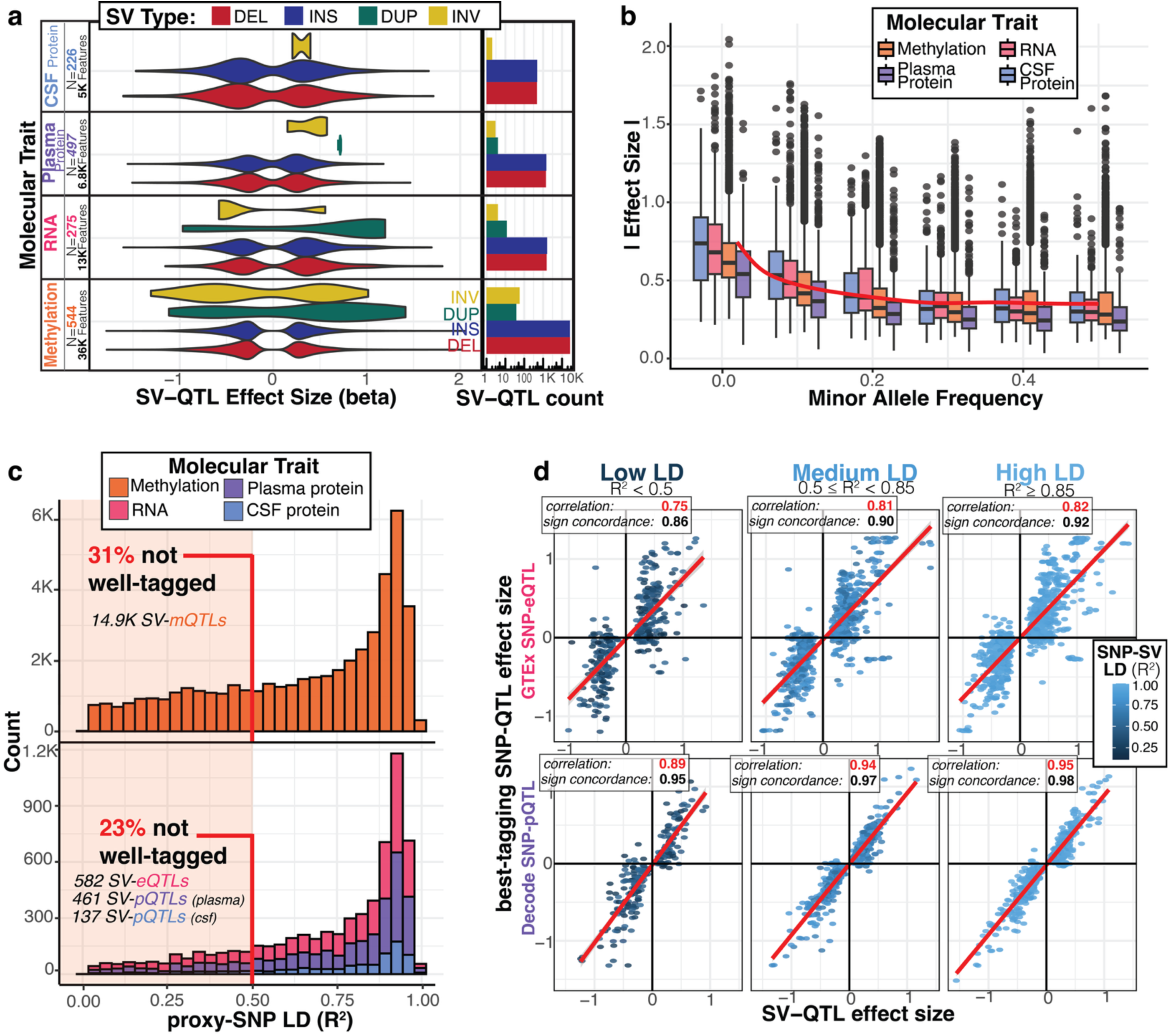
SV-QTL mapping identifies widespread regulatory structural variants not well-tagged by SNVs. **a,** Count and effect size distribution of significant SV-QTL at an FDR threshold of 0.05. Number of samples and features per ome meeting expression and variance thresholds to be tested are also annotated. **b,** Absolute effect size of SV-QTLs stratified by molecular trait and minor allele frequency bins. Red trend line shows loess fit of the relationship between MAF and effect size across all traits. **c,** Histogram of R^2^ correlation between SV-QTL and its best-tagging SNV genotypes; SVs that are poorly tagged (R^2^ < 0.5) are highlighted. Methylation SV-QTLs plotted on separate axes due to the increased number of effects. **d,** Scatter plot comparing SV-QTL effect size to the best-tagging SNV-QTL effect size from two public QTL resources: GTEx eQTLs from whole blood compared to ADRC eQTLs and deCODE pQTLs from plasma to compare to ADRC plasma pQTLs. Scatter plots are split by the strength of LD between SV-QTL and best-tagging SNV. Correlation of SNV and SV betas and sign concordance of betas calculated for each LD bucket.

To investigate the extent to which SV-QTLs represent novel associations, we investigated the LD of these SVs with nearby SNVs. For each SV-QTL, we defined an LD-proxy as the SNV with the highest genotype correlation with the SV. While most SVs were in high LD with their proxy SNV, we found that 32% of methylation SV-QTLs and 23% of expression and protein QTLs had *R*^2^ less than 0.5, representing over 16K unique SV-QTLs. Further, when comparing SV-QTL effect sizes from our QTL study to SNV-QTLs from GTEx v8 whole blood for eQTLs and deCODE genomics for plasma pQTLs, we observed higher correlation and sign concordance between SV-QTL effect sizes with proxy-SNV QTL effect sizes as a function of increasing LD (**Figure 4d**). This indicates that many previously reported QTL associations that relied only on SNV genotypes may be driven by an SV in high-LD.

### Fine-mapped structural variants underlie many QTL associations and exert larger effects than SNVs

To further identify which QTLs are likely driven by SVs instead of SNVs, we combined SV and SNV genotypes and performed statistical fine-mapping of QTLs with Sum of Single Effects (SuSiE) (Wang et al. 2020, McCreight et al. 2025). We labelled each association as to whether an SV was contained in the credible set and, additionally, if it had the highest Posterior Inclusion Probability (PIP) score. Prior work in GTEx using srGS estimated that SVs are likely causal at 3.5% to 6.8% of eQTLs, with SVs as the lead variant in 3.5% of eQTLs (Chiang et al. 2017). With our lrGS SV calls, we observed 12% of QTL associations having an SVs in the credible set, with 6% having an SV as the lead variant (**Figure 5a, Table S4**). 54% of these SVs contained in credible sets were novel to lrGS (**Extended Data Figure 7a**).

**Figure 5.**
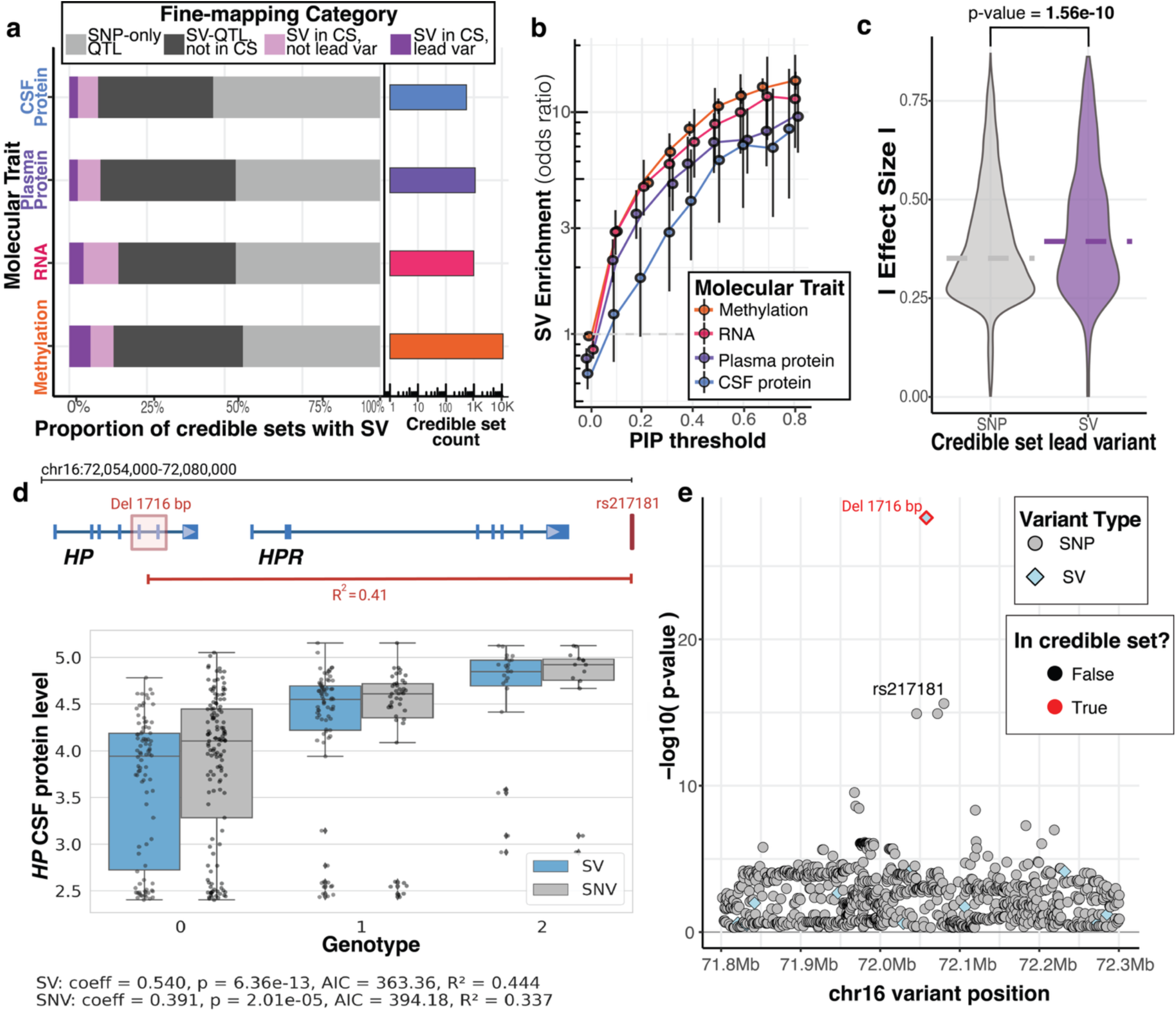
Fine-mapping reveals causal structural variants at QTL loci. **a,** Count of credible sets for each molecular trait, as well as proportion of those credible sets that were fine-mapped to contain an SV. “SNV-only QTL” traits contained no significant SV-QTLs., “SV-QTL not in CS” traits had a significant SV-QTL, but the SV was not included in the credible set. “SV in CS, not lead var” traits contained an SV in one of their credible sets, but the variant with the highest PIP (lead variant) was an SNV. Finally, “SV in CS, lead var” traits were those where an SV was included in the credible set and an SV had the highest PIP. **b,** Enrichment of an SV being a causal variant compared to an SNV across multiple molecular traits. Causality is defined by having high PIP from SuSiE’s fine-mapping at various PIP thresholds. Odds ratio and confidence intervals from Fisher’s exact test are plotted. **c,** Absolute effect size distributions of SV-QTL credible sets stratified by whether an SV or an SNV was the highest pip lead variant. **d,** Schematic of the HP locus with 1.7 kb deletion marked as well as its best-tagging SNV rs217181. Box plot demonstrates SV genotype has a stronger association than SNV with *HP* protein levels in CSF**. e,** Fine-mapping of the *HP* locus confidently puts the deletion variant in the credible set by itself

We observed that SVs were significantly enriched in fine-mapped credible variant sets as a function of PIP threshold (**Figure 5b**). At PIP thresholds >0.8, SVs were enriched in credible sets across each molecular QTL at rates up to 12x higher than SNVs. Across SV types, deletions and duplications were most enriched to be fine-mapped in credible sets (**Extended Data Figure 7b**), and longer SVs that disrupt more sequence were also more likely to be included (**Extended Data Figure 7c**). We also observed significant enrichment of TE-derived SVs, with the strongest signal coming from SINE/Alu and SVA elements, suggesting that specific classes of retrotransposon insertions are disproportionately likely to contribute to regulatory variation captured by QTL fine-mapping (**Supplementary figure 7**).

When the fine-mapped credible set had an SV as the lead variant, we observed a significant increase in the effect size of the QTL (p-value=1.56E-10; **Figure 5c**). Credible sets with a lead SV tended to be smaller than those containing a lead SNV (**Extended Data Figure 7d**). Comparing fine-mapped SVs to other SV-QTLs not included in credible sets, we found lead SVs tended to be in lower LD with nearby SNVs (p-value<1E-32, **Extended Data Figure 7e**). Fine-mapped SVs in QTL credible sets were also more likely to disrupt functional regions compared to background SVs, as evidenced by a higher non-coding constraint score from Gnocchi for these variants (Chen et al. 2024) (**Extended Data Figure 7f**).

We also characterized variant pleiotropy in our QTL data by counting the number of unique features for which variants were fine-mapped. We found that, particularly for methylation, SVs tended to be more pleiotropic than SNVs, being fine-mapped more often into multiple credible sets (**Supplementary figure 8a**). To help explain this pleiotropy, we further explored if SVs could be disrupting 3D genome organization. We found that while controlling for SV length, SVs that disrupted CTCF ChIP-seq peaks or TAD boundaries derived from Hi-C data were more likely to be pleiotropic, being included in significantly more credible sets (TADboundary beta=0.16, p-value=5.12E-6; CTCFpeak beta=0.33, p-value=0.016) (**Supplementary figure 8b**). Most pleiotropic SVs were fine-mapped across multiple features in the same molecular trait, though we did observe 78 SVs fine-mapped across multiple omics layers (**Supplementary figure 8c**).

One AD-relevant fine-mapped variant is a large deletion in HP (chr16:72057133–72058849; 1,716 bp deletion), which spans two exons and results in a different protein isoform: a dimeric form (HP1) when both exons are deleted, showing reduced binding affinity to APOE and hemoglobin (Boettger et al. 2016), in contrast to the tetrameric form when no deletion is present, or the trimeric form when only one allele is affected. In our findings, the 1,716 bp deletion emerged as the lead variant regulating HP protein levels, surpassing the signals from nearby SNVs (**Figure 5d,e**).

### Colocalization with neurodegeneration GWAS nominates structural variants that may confer disease risk

To assess the relevance of these molecular trait associations to neurodegenerative disease, we performed LD and colocalization analyses with Alzheimer’s and Parkinson’s disease GWAS for all our ADRC and SAMS QTLs. High-tagging SVs were identified using a threshold of R² ≥ 0.6 with AD and PD GWAS lead SNVs (**Table S6**). Overall, 10.5% of AD GWAS lead SNVs have at least one high-tagging SV, compared to 6.9% of PD GWAS lead SNVs. We also identified 146 and 67 colocalizations (PP H4 > 0.5) from 46 and 14 unique loci for AD (Bellenguez et al. 2022) and PD (Kim et al. 2024), respectively, across our four molecular traits (**Figure 6a, Table S7, Table S8**). 20.2% of these colocalizations contained a fine-mapped SV in the QTL credible set, with 8 examples of the SV being the lead variant. All together, these represent 8 unique risk loci for AD and 4 for PD that are colocalized with a fine-mapped SV association, representing candidate GWAS signals that are driven by an SV (**Extended Data Table 2)**. To further investigate if these fine-mapped SVs with colocalization evidence could be influencing disease risk, we ran an association of SV genotype on plasma PTau217 levels, a strong blood-based marker for brain amyloid pathology. We found colocalized SVs for AD risk were 10.6 times more likely to be nominally significant (alpha=0.1) compared to background non-colocalized SVs (p-value=8.21E-9) (**Supplementary figure 10a**). Several of these colocalized SVs appear to be enriched in European populations and are substantially rarer in non-European ancestries, suggesting that a subset of these associations may be population-specific (**Supplementary figure 10b**).

**Figure 6.**
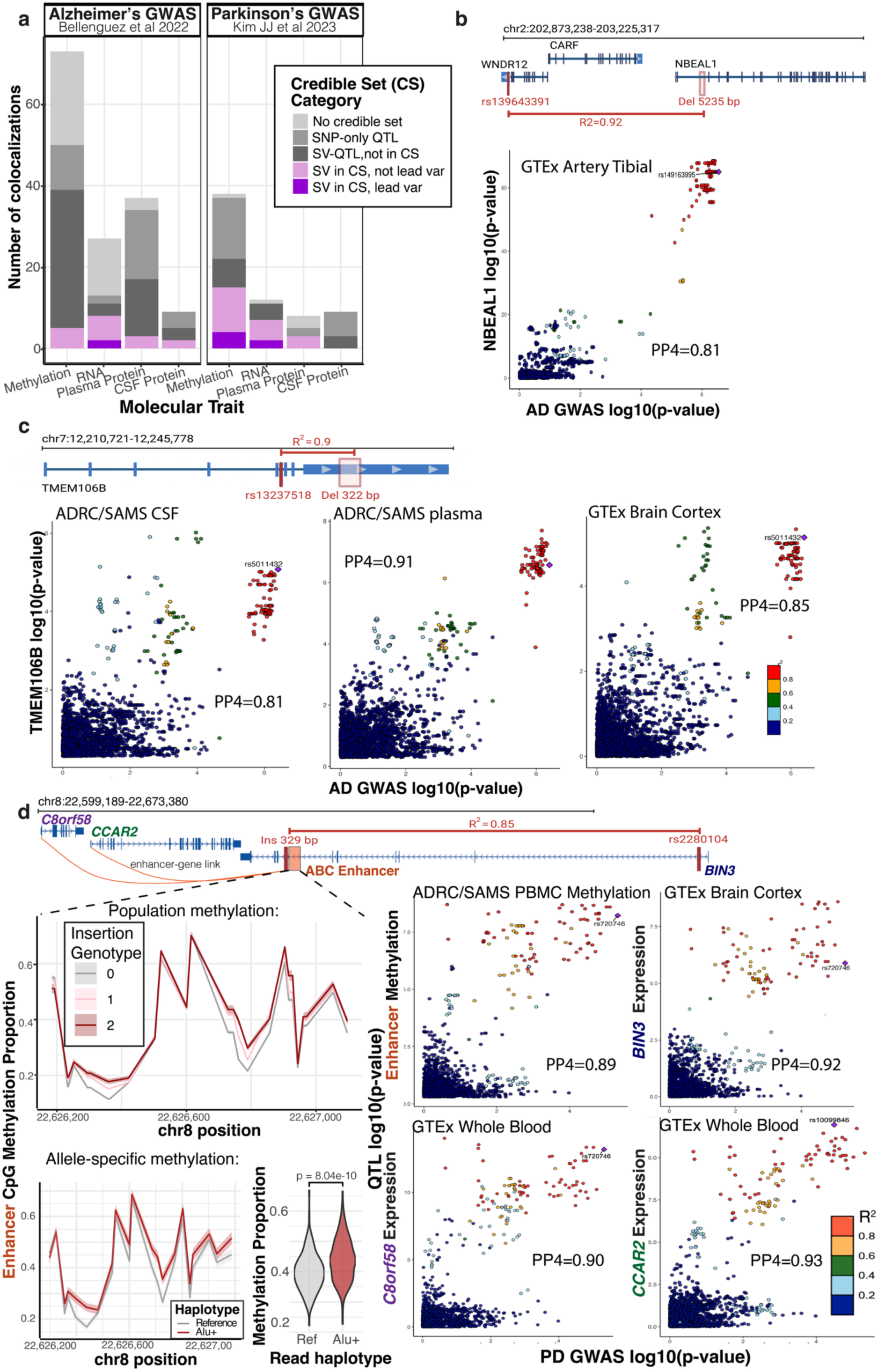
Colocalization across Alzheimer’s and Parkinson’s GWAS in loci with SVs tagging GWAS SNVs. Colocalization plots were generated showing PP4 values and highlighting the top GWAS SNVs at the intersection of the evaluated SNVs with ADRC molecular traits and GTEx expression data. **a,** Number of molecular traits colocalizing with neurodegenerative Alzheimer’s and Parkinson’s GWAS (posterior probability H4 > 0.5). **b,** Intronic deletion in *NBEAL1* in high LD with AD-associated SNV, shown alongside GTEx colocalization with *NBEAL1* expression in the tibial artery and AD GWAS. **c,** Alu element deletion in the 3′ UTR of *TMEM106B* in high LD with an AD-GWAS SNV, colocalizing with ADRC plasma and CSF *TMEM106B* protein levels and for expression in brain cortex from GTEx. **d,** Intronic Alu element insertion in *BIN3* in high LD with a PD-associated SNV, with locus-level colocalization involving ADRC methylation and GTEx brain expression brain cortex. Top line plot shows mean and standard error estimate of methylation of CpGs within the enhancer across the 3 different genotype groups, showing individuals with higher copies of the *Alu* insertion display higher methylation and silencing of the regulatory element. Below, line plot shows allele-specific methylation in 201 individuals heterozygous for *Alu* insertion, plotting methylation at each CpG across enhancer for reads that support either *Alu* or reference haplotype, or averaged across whole enhancer in violin plots. Acitivity-by-contact (ABC) links enhancer to *BIN3*, and nearby genes *C8orf58* and *CCAR2,* whose expression both colocalized with PD risk from Kim JJ et al.

We additionally colocalized GTEx-QTLs with high-tagging SVs to capture associations in tissues not profiled in this study (**Supplementary figure 11-12**). An interesting locus emerged from these AD–GTEx colocalizations for the gene NBEAL1, a protein missing from both aptamer-based and Olink proteomics platforms. We found a large intronic deletion in NBEAL1 (chr2:203034349–203039584; 5,235 bp) that is in high LD with SNVs previously reported in AD (Bellenguez et al. 2022, rs139643391 on WDR12, R^2^ =0.92); small vessel disease (Chung et al. 2019; rs72932727 on ICAIL, R^2^=0.89); and ischemic stroke and white matter hyperintensities (Traylor et al. 2021; rs7934535 on NBEAL1, R^2^=0.82) (**Figure 6b**). QTL variants for NBEAL1 expression in the GTEx Tibial Artery tissue colocalized with Alzheimer’s GWAS (PP4=0.81) and across all neurovascular GWASs (small vessel disease PP4=0.74, stroke and white matter hyperintensities PP4=0.84) (**Extended Data Figure 8**).

We found additional candidate risk SVs from our ADRC molecular trait colocalizations that contained SVs in QTL credible sets. Of particular relevance is an Alu element insertion in TMEM106B (chr7:12242079– 12242401; 322 bp insertion), well tagged by AD GWAS loci (rs13237518 R^2^=0.91) and previously reported (Chemparathy et al. 2024). Also present in the credible sets is an Alu element insertion in ACE (chr17:63488532; 291 bp insertion) (**Supplementary figure 13**), which has been shown to affect isoform splicing (Wu et al. 2013) and may be a relevant locus for AD (R^2^=0.4 with rs4277405 reported in AD-GWAS, R^2^=0.62 with rs4309 in CSF proteomics GWAS) (Cruchaga et al. 2023). The SAMS/ADRC CSF protein levels of the corresponding SNVs in both genes showed strong colocalization with AD GWAS signals (PP4=0.96 ACE CSF protein levels, **Supplementary figure 13**), TMEM106B protein levels (PP4= 0.81 CSF, PP4= 0.91 plasma), as well as GTEx expression (PP4=0.85 brain cortex, and **Figure 6c**).

An ADRC methylation signal colocalized with the *BIN3* Parkinson’s disease locus, where an intronic Alu insertion (chr8:22625890; 329 bp) acts as a methylation QTL for a nearby variable methylation segment (PP4 = 0.89) and is included in the fine-mapped credible set. This methylation segment overlaps a strong ABC enhancer, and individuals with more copies of the insertion exhibited higher rates of methylation across the enhancer CpGs (Fulco et al. 2019). To investigate allele-specific methylation, we assigned reads in insertion heterozygotes based on whether they supported the Alu or reference allele and found that Alu-supporting reads showed significantly higher enhancer methylation. Varying methylation at this regulatory element may modulate how this enhancer regulates its predicted target genes, including CCAR2 and C8orf58. We further observed the expression of these genes in GTEx whole blood, as well as BIN3 expression in cortex and cerebellum, also colocalized strongly with PD GWAS (**Figure 6d**). Using CSF biomarker data in our cohort, individuals with the Alu insertion were more likely to be positive for alpha-synuclein seeding assay (beta=0.162, p-value=0.04), providing orthogonal support for the association of this variant with Parkinson’s disease risk.

High colocalization signals also emerged at the large ∼1 Mbp inversion region spanning *MAPT* and neighboring genes. This region is primarily known for its role in PSP (rs62057121) (Wang et al. 2024), but also contains GWAS hits for both AD and PD. Within the inverted region, numerous SNVs are in high LD (Bowles et al. 2022); however, we also identified SVs encompassing *LINC02210-CRHR1*, *MAPT*, and *KANSL1*, as well as intergenic variants such as an SVA_67 (17:46237502–46238226, 724 bp deletion) located downstream of *LRRC37A*. Interestingly, across ADRC molecular traits, methylation regions and the expression of LRRC37 family genes show high colocalizations with disease risk. SVs are also present within the fine-mapped credible set—for example, LRRC37A2 (ENSG00000238083) shows strong colocalization with PP4 = 0.79 for AD and PP4 = 0.98 for PD (**Supplementary Figures 14**), with a lead insertion (37 bp) in the LRRC37A2 eQTL fine-mapped credible set (**Supplementary Figures 15)**. However, the MAPT inversion region remains a highly complex genetic locus to disentangle, partly due to its structural variability and extended linkage disequilibrium.

## Discussion

In this study, we sequenced 551 nanopore long-read genomes from healthy older controls and patients with AD and PD. The resulting genomes and SV calls are publicly available, enabling assessment of SV allele frequencies and functional impacts for clinical filtering. Coordinates are provided as a downloadable BED file and through an online browser, facilitating identification of variants tolerated in an aging population and prioritization of rare, potentially pathogenic SVs.

Our SV calls are consistent with prior estimates of genome-wide burden, with a median of ∼22,000 SVs per genome (Audano et al. 2019; Chaisson et al. 2019). Our cohort size was sufficient to saturate discovery of common SVs above 1% allele frequency. However, the predominantly European ancestry of our cohort limits the generalizability of these allele frequencies, and additional long-read sequencing across globally diverse populations will be essential to capture the full spectrum of SV diversity (Billingsley et al. 2024; Gustafson et al. 2024; Mahmoud et al. 2024).

As expected, long-read sequencing enabled improved detection of insertions, including repetitive mobile element insertions such as Alu elements. We observed more duplications and inversions by short-read data, likely reflecting both limitations of alignment-based long-read SV callers and differences in sensitivity for read-depth–based approaches. While assembly-based approaches could improve recovery of complex SVs, they were not feasible here given sequencing depth constraints. For fine-mapping analyses, we relied on short-read genotypes for small variants due to higher coverage and accuracy, whereas SV discovery and genotyping were performed using long-read data. Together, this highlights a complementary design in which low-depth long-read sequencing provides improved SV resolution while short-read sequencing remains advantageous for SNV and indel genotyping, although ongoing improvements in long-read accuracy and depth may enable full replacement in future studies. For instance, high-depth ONT genomes sequenced with R10 chemistry and PacBio HiFi have comparable SNP-calling performance to srGS (Olson et al 2022), suggesting population-scale data of these modalities could be used to jointly fine-map SVs and SNPs from a single technology, with the additional benefit of phasing to detect allele-specific affects.

We sought to define the contribution of structural variants (SVs) to neurodegenerative disease risk. Through fine-mapping and colocalization analyses, we found 11 candidate loci where an SV may underlie molecular trait associations that colocalize with AD or PD GWAS signals. In addition, LD-based analyses revealed up to 20 loci where lead GWAS SNPs strongly tag nearby SVs, suggesting that SVs may contribute to disease risk even at loci where they were not directly fine-mapped. These loci may reflect regulatory mechanisms not captured in our study, including effects on splicing or activity in disease-relevant tissues or contexts not profiled here. Notably, we found up to 33% of SV-QTLs are poorly tagged by nearby SNVs, indicating that a substantial fraction of SV-mediated regulatory variation may also be missed by conventional GWAS. At the HP locus, for example, we fine-mapped a 1.7 kb deletion spanning two exons—absent from srGS and poorly tagged by SNVs—as the causal variant driving differences in HP protein levels in both plasma and CSF. This previously identified deletion is known to influence HP isoform structure, which has been implicated in AD and PD risk (Bai et al. 2023; Costa-Mallen et al. 2015), though is not a GWAS loci potentially due to the lack of tagging SNPs. As population-scale long-read datasets expand, direct association testing of SVs or imputation from SNP haplotypes will be important for evaluating their contribution to disease risk. Importantly, while LD and QTL fine-mapping help prioritize candidate SVs, functional validation of SVs will still be required to distinguish true causal variants from those just tagging broader risk haplotypes.

A notable feature of our dataset is the prominent role of transposable element (TE)–derived SVs, particularly Alu and SVA insertions, in regulatory variation. We observed enrichment of TE-derived SVs within fine-mapped QTL credible sets, supporting their contribution to functional regulatory variation. Further, many of the candidate risk SVs we prioritized were *Alu* elements including those in *ACE, TMEM106B,* and *BIN3.* Beyond simple disruption of existing regulatory elements, TEs may introduce novel regulatory activity, including new transcription factor binding sites, CpG-rich regions influencing methylation, or altered chromatin interactions. To support this, recent work has demonstrated that Alu elements can contribute to enhancer–promoter connectivity and chromatin looping (Liang et al. 2023), highlighting the potential for SVs to create new regulatory architecture rather than solely perturb existing elements. At the *BIN3* locus, an Alu insertion was associated with higher methylation at a nearby enhancer, including allele-specific hypermethylation of Alu-containing haplotypes. We speculate that this may reflect TE silencing and the allele-specific spread of methylation into the nearby enhancer. Further, at the *ACE* locus, an intronic *Alu* insertions has been reported to lead to splicing and isoform changes, including exonization of the element (Lei et al 2025). Future work integrating orthogonal assays—including ChIP-seq, chromatin conformation mapping, and cell-type–resolved transcriptomic and epigenomic profiling—will be needed to systematically map the diverse regulatory effects of TE-derived SVs across molecular layers.

Our study also highlights several limitations. First, RNA-seq data were generated using a 3′ single-cell protocol and aggregated to pseudobulk profiles to maximize power, which limits resolution of cell-type-specific and splicing-related effects. Although targeted cell-type–specific analyses identified additional SV-QTLs and context-specific regulatory effects, these analyses were underpowered for less abundant cell types, and comprehensive assessment of splicing consequences will require full-length or long-read transcriptomic data. Second, while long-read sequencing enables phasing, limited coverage restricted our ability to perform genome-wide allele-specific methylation analyses, although targeted analyses at specific loci support haplotype-specific regulatory mechanisms. Third, current functional annotation frameworks for SVs remain less developed than for SNVs, particularly for variants that introduce or reconfigure regulatory elements—such as transposable element insertions or structural rearrangements—potentially limiting sensitivity of computational prioritization approaches. Finally, our analyses are largely based on blood-derived molecular phenotypes, which may not fully capture regulatory effects occurring in disease-relevant brain cell types.

In conclusion, by combining long-read genome sequencing with multi-omic functional data, we identified thousands of SVs missed by short-read sequencing and fine-mapped a subset with potential regulatory and disease relevance. Our results highlight the importance of including SVs, particularly those detectable only with long-read technologies, in models of genetic architecture for complex diseases like AD and PD.

## Methods

### Ethics statement

All ADRC and SAMS participants provided written informed consent under Stanford IRB–approved protocols for the use of their data in IRB-approved research.

### Cohorts description

Participants in the Stanford Alzheimer’s Disease Research Center (ADRC) were enrolled through the NIA-funded Stanford ADRC, a longitudinal cohort comprising individuals diagnosed with dementia and cognitively normal elder controls. Diagnostic status was assigned in a consensus conference of board-certified neurologists and neuropsychologists using the Clinical Dementia Rating scale and NACC procedures. Stanford ADRC participants receive neurological exams, comprehensive neuropsychological evaluations, MR and amyloid PET imaging, and donate blood and CSF samples. They are invited back annually for follow-up assessments.

The Stanford Aging and Memory Study (SAMS) is an ongoing NIA-funded longitudinal investigation of healthy aging, enrolling participants who demonstrate no cognitive impairment (Trelle et al. 2021, Sheng et al. 2025). All blood collection, processing protocols, and neuropsychological assessments mirrored those used in the ADRC to ensure methodological consistency. At baseline, each SAMS participant exhibited a Clinical Dementia Rating of 0 and achieved age-adjusted normative scores on a standardized neuropsychological battery; cognitive status was confirmed by the same multidisciplinary consensus panel.

Of note, some participants were enrolled in both ADRC and SAMS. In the Stanford ADRC and SAMS cohorts, comprehensive biomarker assessments were conducted to evaluate amyloid and tau pathology. Cerebrospinal fluid (CSF) levels of Aβ42, Aβ40, p-tau181, and total tau were measured using Lumipulse G assays, with a CSF Aβ42/Aβ40 ratio below 0.091 indicating amyloid positivity. Plasma p-tau181 and Aβ42/Aβ40 ratios were also assessed using modified Lumipulse assays, demonstrating high concordance with CSF and PET measures. Amyloid PET imaging with 18F-Florbetaben was performed on a subset of participants, and standardized uptake value ratios (SUVRs) were calculated using a global cortical composite relative to the whole cerebellum.

### Defining clinical diagnoses groups

To define neurodegenerative disease groupings of the lrGS sequenced ADRC and SAMS participants, we relied on the comprehensive biomarker data and cognitive assessments as described in a previous study (Winer et al. 2025). In brief, participants underwent diagnostic adjudication at multidisciplinary consensus meetings, which included a panel of neurologists, neuropsychologists, and research staff through their involvement in ADRC or SAMS study. Participants diagnosed clinically with mild cognitive impairment due to AD (MCI) or Alzheimer’s Disease (AD) met criteria for National Institutes of Health Alzheimer’s Disease Diagnostic guidelines and were confirmed to have biomarker evidence of AD using Amyloid-PET and CSF Aβ42/40. Parkinson’s was also diagnosed using the UK Brain Bank criteria and required bradykinesia with muscle rigidity and/or rest tremor. Parkinson’s diagnoses were further split into PD with MCI (PD-MCI) if they had cognitive complaint and objective impairment on cognitive testing, without substantial impact on functional activities, or Parkinson’s disease dementia / Lewy body dementia (PDD/LBD) as cognitive impairment severe enough to interfere with activities of daily living as determined by clinical history and Clinical Dementia Rating.

### Long-read sequencing protocol

Long-read genome sequencing (lrGS) was performed on 575 participants. We obtained genomic DNA from NCRAD whole blood samples or extracted high molecular weight DNA from primary blood mononuclear cells stored at -80 °C using a Puregene kit (Qiagen, Germany). DNA was then sheared using a G-tube (Covaris LLC, Massachusetts). Sequencing libraries were prepared using Nanopore LSK-110 and sequenced on the PromethION48 following standard protocol (Oxford Nanopore Technologies, United Kingdom). We aimed for 60 GB of estimated bases sequenced per sample during sequencing, at which point the run was stopped and the flow cell was washed and then used with the Nanopore WASH-003 kit to be used on an additional sample. We sequenced an average of 1.8 samples per flow cell. After sequencing QC, an average of 50.4 gigabases were sequenced per sample, with a read length N50 of 18 kb. Sequencing data were basecalled using Guppy (High Accuracy, version 6.3) and aligned to hg38 using Minimap2.15. Structural variants were called using Sniffles216 in population mode. Clair3 was used to call SNVs and short indels from lrGS bams and 24 samples were excluded when IBD analysis with srGS variant calls revealed sample swaps or cryptic nonmatching between the lrGS and srGS sequencing.

### Short-read sequencing protocol

Short-read whole-genome sequencing (sWGS) was performed on 588 participants. The majority (n=554) were sequenced using MGI high-throughput sequencing on DNBSEQ-T10 (BaseNum > 90G, Q30 > 85). The rest (n=34) were sequenced as part of the Stanford Extreme Phenotypes in Alzheimer’s Disease project, with sequencing performed at the Uniformed Services University of the Health Sciences (USUHS) on an Illumina HiSeq platform. The Genome Analysis Toolkit (GATK) workflow Germline short variant discovery was used to map genome sequencing data to the reference genome (GRCh38) and to produce high-confidence variant calls using joint-calling (Poplin et al., 2017). Six individuals were excluded from further srGS analysis due to discordance between their reported sex and genetic sex.

### Transcriptomics and proteomics normalization

Single-cell RNA sequencing data were generated for ADRC and SAMS individuals using DNBelab C4 system from MGI, as previously described (Grandke et al. 2025). To map QTLs and call outliers, these single-cell data were aggregated into pseudobulk profiles by summing gene read counts across all barcodes per individual. Library size was estimated from aggregated counts, and for individuals sequenced at multiple time points, we retained the sample with the largest library size. Genes with low variance were excluded using coefficient of variation with the modelGeneCV2 function from scran package and filterByExpr from edgeR leaving 13K genes (Lun, McCarthy, and Marioni 2016; Robinson, McCarthy, and Smyth 2010). Counts were log-transformed, and library-size normalized using edgeR package. The resulting sample-by-gene matrix was then rank-normalized for QTL mapping, and principal component analysis (PCA) was applied to estimate global hidden factors.

For outlier detection, normalized counts were instead scaled and centered to z-scores. PCA was performed on this z-score matrix, and top 120 PCs were regressed out along with age and sex. Residuals were re-scaled to generate final expression z-scores. Individuals with an excessive burden of outlier genes (|z-score| > 3) were removed as global outliers.

Proteomics data were generated using the SomaScan v4.1 platform. Following Soma<u>L</u>ogic-recommended normalization, calibration, and QC, raw aptamer intensities were log-transformed and normalized as previously described (Oh et al. 2023). For plasma samples, where most individuals had multiple time points, the sample with the highest connectivity z-score was retained. Low-expression aptamers were removed using edgeR *filterByExpr*, leaving 6.8K aptamers for plasma and 5K for CSF. The normalized matrices were rank-normalized for QTL mapping, and PCA was performed to capture phenotype structure. Outlier detection was performed analogously to RNA: log-transformed protein values were scaled, residualized for age, sex, and top PCs, re-scaled to z-scores, and global outlier samples (excess |z-score| > 3 aptamers) were excluded. SomaScan aptamers were mapped to genomic coordinates by first mapping associated UniProt IDs to corresponding Ensembl gene ids and coordinates using BioMart.

### Rare variant enrichment

For rare variant enrichment analyses, protein abundance and RNA expression were residualized for age, sex, disease status, batch, and principal components (120 for RNA, 30 for plasma proteomics, and 40 for CSF proteomics), selected to maximize cross-omic outlier concordance. Residuals were standardized as z-scores, and measurements with |z| > 3 were classified as outliers. For overall summaries of outlier burden per genome, we used a lower threshold of |z| > 2 to capture lower-magnitude events that may still reflect aberrant regulation.

Methylation outliers were identified using METAFORA with |z| > 2.5 and an absolute methylation difference >0.25, as recommended by default METAFORA parameters (Jensen et al. 2026). Genes were assigned expression or protein outlier status from their corresponding measurements and methylation outlier status when a methylation outlier occurred within 10 kb. Genes were classified as rare-SV carriers when an SV with AF < 0.01 occurred within 10 kb. For each pair of molecular traits, we restricted analysis to genes with at least one outlier measured in both datasets and tested enrichment of concordant or discordant outlier states using Fisher’s exact test. We further varied the z-score, SV distance, and allele-frequency thresholds used to define outliers and rare SVs-genes to assess their effects on enrichment strength.

### Methylation data processing and normalization

5mC CpG methylation was called from Oxford Nanopore data using nanopolish/f5c on aligned BAM files (Simpson et al. 2017; Gamaarachchi et al. 2020). Nanopolish methylation calls were summarized with *calculate_methylation_frequency.py*, applying a log-likelihood ratio threshold of 1 to generate per-CpG methylation proportions (beta values) and coverage. These calls were reformatted and indexed for input into METAFORA.

METAFORA aggregated methylation profiles across all samples to generate a tissue mean, then segmented CpG methylation on autosomes into 303K correlated clusters. Per-sample methylation betas were aggregated within segments, producing a sample-by-segment beta matrix. Segments were filtered for variability by transforming betas as *logit(1 – |0.5 – β|)*, which symmetrizes hyper/hypomethylation, mapping values near extremes of 0 and 1 to values close to zero and mapping high-variance segments with mean betas near 0.5 to higher positive values. The coefficient of variation (CV²) of transformed values was compared against a loess-estimated mean–variance trend, and 36K segments with residuals >3 standard deviations were defined as variable segments. This subset was rank-normalized for QTL input, and PCA was performed to estimate hidden factors.

Methylation outliers were called with METAFORA using minimum absolute z-score threshold of 2.5, and minimum outlier delta of 0.25. Global outlier samples who had above the tukey outlier limit of total methylation outlier counts per genome were excluded from enrichment analyses.

### QTL mapping

Structural variant (SV) genotypes were obtained from Sniffles2 joint-genotyping VCFs. SVs missing in >50% of individuals were excluded. Minor allele frequencies (MAF) were computed within ADRC, and only common SVs (MAF ≥ 0.02) were retained. Missing genotypes were imputed using missMDA package *imputePCA*, and PCA on the imputed matrix provided genotype PCs, which recapitulated self-reported ancestry.

QTL mapping was performed with *fastLLM* (R), chosen because tensorQTL cannot handle multi-base SV intervals when overlapping cis-windows. A 1 Mb cis-window around each phenotype coordinate (gene TSS, protein-coding gene, or methylation segment) was intersected with SV positions. For each overlap, linear models were fit with phenotype rank-normalized values as response, SV genotype as predictor, and covariates including sex, age, AD/PD diagnosis, and an optimized number of phenotype PCs. The optimal PC number was chosen by maximizing discovery yield (RNA = 30, plasma = 40, CSF = 30, methylation = 7), (Supp. Fig. 4d). From the final model, effect sizes and standard errors were reported. P-values were corrected across tests using the Benjamini-Hochberg procedure, and associations with adjusted p < 0.05 were retained.

### SuSiE fine-mapping

Fine-mapping of SV QTLs was performed jointly with short-read SNVs/indels using individual-level SuSiE (Wang et al. 2020, McCreight et al. 2025). The SV genotype matrix was subset to individuals with matching srGS data. For each phenotype, a 1 Mb cis-window (around TSS for expression/protein, or methylation segment coordinates) was intersected with lrGS SV and srGS SNV/indel calls (via plink). Genotypes were combined, imputed using missMDA *imputePCA*, then scaled and centered. Phenotype residuals (after covariate regression from the SV-QTL model) were likewise scaled.

SuSiE was run with *L = 10* and *min_abs_corr = 0.25*. For each credible set, the variant with highest PIP was designated the lead, and sets were annotated as SV-led or SNV-led. Credible sets were also annotated as containing an SV in credible set even if it was not lead. Enrichment for SVs among credible sets was assessed by applying PIP thresholds and testing enrichment of SVs compared to SNPs using Fisher’s exact test.

### GWAS colocalization

To colocalize ADRC/SAMS QTLs with neurodegenerative GWAS, we obtained summary statistics from Alzheimer’s disease (Bellenguez et al. 2022) and Parkinson’s disease (Nalls et al. 2019; Kim et al. 2024), as well as QTL resources (GTEx eQTLs, deCODE and UKBB plasma pQTLs, and Knight-ADRC CSF pQTLs) (GTEx Consortium 2013; Ferkingstad et al. 2021; Sun et al. 2023; Western et al. 2024). SNP summary statistics were harmonized, sorted, and indexed across each data set. For ADRC/SAMS QTLs, we used SNP beta and p-value summary stats from srGS from the susieR fine-mapping for methylation, RNA, plasma protein, and CSF protein phenotypes.

Significant loci were defined using windows of 100 kb around variants surpassing significance threshold of p < 1e-5 for GWAS data sets and p < 1e-4 for QTL data sets. Overlapping windows containing both a GWAS and QTL signal were tested with *coloc.abf* using p-value-only method (Rasooly, Peloso, and Giambartolomei 2022). We report the posterior probability (PP.H4) of a shared causal variant, retaining colocalizations with PP.H4 > 0.5. Selected loci were visualized using locusCompareR (Liu et al. 2019). For colocalizations with ADRC/SAMS QTL, the colocalizations were annotated based on SV fine-mapping of the corresponding ADRC/SAMS QTL. We highlight colocalizations where the SV was contained in the credible set of the QTL as a proxy for SVs that are also influencing risk in the GWAS.

Risk genes were defined from eQTL and pQTL colocalizations with public QTL resources (GTEx, deCODe, UKBB, Knight-ADRC): for each gene, we retained the maximum PP.H4 across all tissues, and genes with PP.H4 > 0.5 for AD or PD GWAS were included in the prioritized set for rare SV interpretation.

### WatershedSV rare variant annotation collection pipeline

Raw molecular measurements from each omic dataset, including RNA expression, plasma proteomics, and CSF proteomics, were first processed and normalized. To enhance concordance between RNA and protein outliers, RNA reads and protein abundance values were further corrected using principal components: 105 for RNA expression data, and 25 for plasma proteomics, and 25 for CSF proteomics. The data were then scaled and centered, and only genes with an absolute z-score greater than 2 were retained and input into WatershedSV. Z-scores were converted to binary variables to indicate outlier status.

The WatershedSV method was adapted from Jensen et al. (2024) to identify rare SVs with potential functional impact. Whole genome sequencing (WGS) data were processed according to the WatershedSV pipeline previously described. Briefly, rare variants which were annotated with nearby genes within a 100 kb window were identified. These gene-variant pairs were then paired with corresponding genomic annotations, which were used as input for the WatershedSV model, along with outlier data computed above.

WatershedSV was applied separately to generate RNA expression, plasma proteomics, and CSF proteomics models using the dataset from the ADRC cohort. For each model, model weights were learned and stratified by annotation categories, including “Coding”, “Conservation track”, “Encode chromHMM 25 states,” “Noncoding,” “Regulatory,” and “SV”. For each omic layer, the model integrated genomic annotations with molecular outlier status to compute posterior probabilities for each rare SV. Variants with high posterior scores were considered likely to have a functional molecular impact.

The WatershedSV pipeline outputs posterior probabilities ranging from 0 to 1 for each rare SV. Downstream model evaluation involved N2 pair prediction and performance benchmarking against a baseline Genome Annotation Model using the area under the precision-recall curve (AUPRC).

### Repeat class annotation

We used RepeatMasker (Smit et al. 2025) to annotate repeat classes of lrGS-discovered insertions and deletions. From the lrGS merged VCF we extracted all insertion sequences and used *bedtools* getFasta to convert deletion coordinates to reference sequence from GRCh38. The resulting fasta was input to RepeatMasker. Indels were assigned as repeats if over 70% of their sequence were annotated/masked by RepeatMasker. In the case that multiple repeats were discovered in an indel, each variant was assigned to the repeat with the longest aligned sequence match. To calculate lrGS-specific variant enrichment among different repeat classes, we used a fisher’s exact test on whether variant is lrGS-specific or srGS-shared and whether variant is in given repeat class or not (for each repeat class) to estimate odds ratios, confidence intervals, and p-values. Similarly, we used a fisher’s exact test to calcualte enrichment of repeat classes among credible set. For each molecular trait and repeat class, we used a fisher’s exact test on whether variant was fine-mapped in at least one credible set across all features and if variant was in given repeat class.

### Genome stratification enrichment

To assess regions of the genome where variants are more likely to be shared with srGS or lrGS, we used GRCh38 bed file genome stratifications from Genome in a bottle (GIAB) (Dwarshuis et al. 2024) downlaoded from (https://github.com/usnistgov/giab-stratifications). We used four bed file stratifications including tandem repeats, homopolymers, and low complexity sequences (tandem; GIAB/LowComplexity/GRCh38_AllTandemRepeatsandHomopolymers_slop5.bed.gz), segmental duplications (segdup; GIAB/SegmentalDuplications/GRCh38_segdups.bed.gz), and low-mappability regions based on srGS (lowmap; GIAB/mappability/GRCh38_lowmappabilityall.bed.gz). In addition a “notindifficult” union region was also downloaded, representing what we call “easy” sequences for srGS that excludes all of the previous repeat and low complexity regions (easy; GIAB/union/GRCh38_notinalldifficultregions.bed.gz). Each SV was labeled as members of one of these four stratifications based on bedtools intersection of coordinates (regions are not exclusive, so SV could be member of multiple stratifications). To assess overlap of lrGS and srGS across these stratifications, we used the truvari tool to compare our lrGS variants with srGS calls from gnomAD SV v4 using each of the stratification bed files (English et al. 2024) ‘truvari bench -b “$BASE_NORM.gz” -c “$COMP_NORM.gz” -o “$BENCH_DIR” --refdist 500 --pctsize 0.50 --pctseq 0 --sizemin 50 –passonly’. Enrichments were then calculated from a fisher’s exact test over the 200,232 SVs.

### lrGS variant overlap with population databases

To assess overlap of our lrGS-specific with other population resources, we downloaded SV calls from two population resources, Human Genome Structural Variation Consortium version 3 (Logsdon et al. 2025; https://ftp.1000genomes.ebi.ac.uk/vol1/ftp/data_collections/HGSVC3/release/Variant_Calls/1.0/GRCh38/), and 500 genomes sequences from the 1000 Genome Project Long Read Sequencing Consortium (Gustafson et al. 2024; https://s3.amazonaws.com/1000g-ont/ PROCESSED_DATA/ALIGNED_TO_HG38/SNIFFLES_v2.6.2). The 1000G genome’ sample-level sniffles VCF files were merged with jasmine to create a cohort-level SV vcf. Overlap was determined with the truvari bench command requiring 50% reciprocal overlap of SV lengths and start positions within 500bp.

### BIN3 Allele-specific Methylation

To calculate allele-specific methylation at the *BIN3 Alu* insertion, we first used the merged lrGS vcf file to identify all 201 heterozygous individuals for the insertion, and the SV ID of the insertion in each of the het individuals. Then in each individual sample-level vcf, we used the SV ID to find and extract read names for all the supporting reads of the insertion, assuming reads not supporting the insertion were on the reference non-*Alu* haplotype. Then, we subset the raw nanopolish/f5c output to just the enhancer methylation sequence CpGs and labeled reads based on read_ID as reference or Alu+ (read supporting Alu insertion call). Haplotype-specific methylation was plotted across each individual CpG and then averaged per haplotype across all enhancer CpGs in the 201 individuals to calculate a paired two-tailed difference of means t-test for methylation levels between the reference and *Alu* haplotype.

### SV pleiotropy and genome organization

To compare pleiotropy of SVs to SNPs, we used the SuSiE fine-mapping results across our 4 molecular traits. For each SV we counted the number of distinct features in each molecular trait where the variant was fine-mapped into a credible set. We defined pleiotropic variants in more than 1 credible set. For each molecular trait, we calculated percent of fine-mapped variants that are pleiotropic stratified by whether variant was a SNP or SV. Further, in each molecular trait fisher’s exact test was used to calculate pleitropic variant enrichments among fine-mapped SVs or SNPs. To assess if structural variants were disrupting potential genome organization or topologically associated domains (TADs), we downloaded GM12878 lymphoblastoid cell line Hi-C TAD boundaries from Rao et al. 2014, and CTCF ChIP-seq binding peaks from the same cell line from ENCODE (ENCFF017XLW; ENCODE Project Consortium et al. 2012, Kagda et al. 2025). TAD boundaries were extracted and slopped on either side by 20kb to account for the resolution of Hi-C. Variants were labeled as disrupting either TAD boundaries or *CTCF* binding based on coordinate intersection with these annotations. Generalized linear model was then used to regress variant pleiotropy (number of features where SV was fine-mapped across all traits) on whether SV intersected TAD or *CTCF* coordinates, adding SV size as a covariate to control for effects of SV length. Given high correlation of TAD and *CTCF* coordinates, we tested effects of these variables in two separate models rather than modeling together.

## Data Availability

Summary statistics can be downloaded and queried at https://qsushiny.shinyapps.io/adrc_lrs/
Code for manuscript is available at https://github.com/tjense25/ADRC-Flagship-lrGS-Code
Data from specific Stanford cohorts can be requested to the following cohort leaders: ADRC, M.G.D or T.W-C.; SAMS, E.M. or A.D.W.

https://qsushiny.shinyapps.io/adrc_lrs/

## Code and Data Availability

Summary statistics can be downloaded and queried at https://qsushiny.shinyapps.io/adrc_lrs/. Data from specific Stanford cohorts can be requested to the following cohort leaders: ADRC, M.G.D or T.W-C.; SAMS, E.M. or A.D.W. Workflows and scripts for processing of lrGS and multi-omics data, and fine-mapping and colocalizing QTLs are available at https://github.com/tjense25/ADRC-Flagship-lrGS-Code.

## Acknowledgements

We thank members of the Montgomery, Ashley, and Greicius laboratories for helpful discussions and technical assistance. We thank Bohan Ni for help implementing WatershedSV on our ADRC data. We thank the lab of Andreas Keller for their help providing ADRC single-cell RNA-seq data. T.D.J. was supported by U01HG011762, T32HG000044. L.T. is funded by the European Union – NextGenerationEU, Mission 4, Component 2, under Project SOE2024_0000169 (LATENT-AD; CUP: D93C25000390007). This study was supported by grants from the NIH R35AG072290 (M.D.G), E.C., R01AG074339 (E.C.M.), R01AG048076 (A.D.W.). This work utilized computing resources provided by the Stanford Genetics Bioinformatics Service Center, supported by National Institutes of Health Instrumentation Grant S10OD025082, and would not have been possible without the support of the Stanford SCG cluster system administrators. We thank all participants and staff of the Stanford Alzheimer’s Disease Research Center and the Stanford Aging and Memory Study for their essential contributions.

## Author Contribution

T.D.J., Y.L.G, T.W.C., S.B.M., E.A.A., and M.D.G. conceived of the study. T.D.J., Y.L.G., I.F.S., A.P.T., D.N., T.M.A., J.Z., and M.D.G. helped with curation of the genomic and multi-omics data. T.D.J., Y.L.G., L.T., S.Y., and M.D.G. performed formal analysis and statistical modeling. T.D.J., J.E.G., and A.F. performed investigation by performing the long-read sequencing. A.D.W., V.W.H., E.C.M., K.L.P., T.W.C., and M.D.G. performed investigation by coordinating participant recruitment. T.D.J., Y.L.G., L.T., and S.Y. developed bioinformatics analysis pipelines and methodology for this study. D.N., J.L., R.C.P., J.P., and Z.H. provided additional help with methodology and statistical methods. T.D.J., Y.L.G., L.T., S.Y., and A.P.T. generated figures and data visualizations. T.D.J., Y.L.G., L.T., S.Y., S.B.M., M.D.G. wrote original manuscript. All authors helped review and edit work. E.C.M., K.L.P., A.D.W, T.W.C., V.W.H., and M.D.G. obtained project funding for ADRC and SAMS cohorts and long-read sequencing. E.A.A. and S.B.M. provided additional resources. S.B.M., E.A.A., and M.D.G. supervised the study.

## Supplementary Results

### Fine-mapping cell-type–resolved expression

In this study, single-cell RNA-seq data were aggregated across droplets to generate pseudobulk expression profiles for QTL mapping, primarily to maximize statistical power given the study design. The single-cell dataset prioritized breadth of sampling (310 individuals, including longitudinal sampling across multiple timepoints) over per-cell sequencing depth, resulting in a median of 2,639 cells per sample, with median coverage of 1,076 UMIs and 597 genes detected per cell (Grandke et al. 2025). Under these conditions, aggregation across cells increases sensitivity to detect regulatory effects. In addition, a bulk-like representation of expression enabled consistency across molecular modalities, as methylation and proteomic measurements were derived from bulk samples.

Despite this, we were curious to explore potential context-dependent regulatory effects, so we performed additional cell-type–resolved QTL analyses. Pseudobulk expression profiles were generated within annotated PBMC cell types (CD4 T cells, CD8 T cells, NK cells, B cells, and monocytes), requiring a minimum of 30 cells per cell type per sample (**Supplementary figure 9a**). After applying expression and variance filters, the number of retained genes and samples varied across cell types, largely reflecting differences in cell abundances. For example, CD4 T cells retained 310 samples and ∼16k genes, whereas monocytes only retained 182 samples and ∼12k genes. Less abundant cell types exhibited a higher proportion of genes uniquely testable within that context, not meeting inclusion filters in combined tissue bulk profile (49% in monocytes versus 32% in CD4 T cells; **Supplementary figure 9b**). This indicates that cell-type–resolved analyses allowed us to capture regulatory effects in additional genes not observable in bulk.

To map cell-type-specific QTLs, we applied joint SuSiE-based fine-mapping of SVs and SNVs to each cell-type–specific dataset. Across the five PBMC cell types, this analysis identified 910 additional eGenes (genes for which SuSiE identified a credible set) and 432 additional fine-mapped SVs not detected in the bulk analysis. The number of detected credible sets scaled with effective sample size and cell abundance: CD4 T cells yielded 888 credible sets, comparable to bulk (n = 1,046), whereas monocytes yielded 217 (**Supplementary figure 9c**). Notably, most eGenes and fine-mapped SVs were cell-type-specific (**Supplementary Fig. 9d,e**). 27.9% of cell-type-specific eGenes were from uniquely expressed genes not meeting expression filters in the bulk profile, the remaining being expressed in bulk but only identified a fine-mapped variant effect in a specific cell-type. To assess whether transposable element (TE)–derived SVs exhibit cell-type-specific regulatory activity, we compared the distribution of TE classes among fine-mapped SVs across cell types. We did not observe significant differences in TE composition (e.g., Alu/SINE enrichment; chi-squared test p = 0.82), suggesting that, at the resolution of these data, TE-associated regulatory effects are not strongly cell-type-restricted.

Of interest to Alzheimer’s disease, we explored fine-mapped SVs in monocytes because they share the same myeloid lineage as microglia—a cell-type enriched for a substantial proportion of AD heritability. In monocytes, we detected 39 genes containing an SV in their fine-mapped credible set. Among these, we found a 32bp deletion (chr7:100297288-100297320) that was fine-mapped as lead variant for *PILRB* expression—a gene whose protein levels have colocalized with AD risk (Zhan et al. 2025)—in monocytes but not in any other cell type or bulk profile. Further, a 76bp deletion (chr6:32577186 – 32577262) was included in variant credible set for HLA-DQA1 expression, another AD risk gene. This analysis shows mapping QTLs at cell-type resolution, could yield additional Alzheimer’s risk variants; yet these analyses should be interpreted with cautions as limited depth and sample size could yield increased noise, which may both reduce power and increase the likelihood of spurious associations, particularly in less abundant cell types.

We share a gene-level fine-mapping summary table (Supplemental table S8) as well as full variant summary stats from SuSiE-based fine-mapping for each cell type through the accompanying browser.

## Extended Data Tables

**Extended Data Table 1.** Demographics long-read sequencing cohort by diagnosis group.

| Latest diagnosis groups: | HC | AD | MCI | PD.MCI | PD | LBD/PDD | Other |
| --- | --- | --- | --- | --- | --- | --- | --- |
| **N IrGS** | 300 (54.4%) | 67 (12.2%) | 79 (14.3%) | 33 (6.0%) | 39 (7.1%) | 23 (4.2%) | 10 (1.8%) |
| Age at collection, mean (SD) | 71.1 (7.6) | 72.4 (10.8) | 75.8 (7.2) | 69.6 (9.2) | 69.2 (6.5) | 73.6 (6.1) | 76.5 (8.9) |
| Age at latest diagnosis, mean (SD) | 72.9 (8.4) | 74.3 (11.0) | 77.7 (7.7) | 72.5 (9.0) | 72.1 (6.6) | 75.2 (6.4) | 79.0 (9.5) |
| Sex, female n (%) | 176 (58.7%) | 35 (52.2%) | 36 (45.6%) | 11 (33.3%) | 19 (48.7%) | 6 (26.1%) | 6 (60.0%) |
| **Race** |  |  |  |  |  |  |  |
| White | 257 (85.7%) | 50 (74.6%) | 59 (74.7%) | 32 (97.0%) | 39 (100.0%) | 23 (100.0%) | 5 (50.0%) |
| Asian | 22 (7.3%) | 4 (6.0%) | 8 (10.1%) | 1 (3.0%) | 0 (0.0%) | 0 (0.0%) | 1 (10.0%) |
| Black or African American | 2 (0.7%) | 2 (3.0%) | 2 (2.5%) | 0 (0.0%) | 0 (0.0%) | 0 (0.0%) | 0 (0.0%) |
| More Than One Race | 4 (1.3%) | 0 (0.0%) | 0 (0.0%) | 0 (0.0%) | 0 (0.0%) | 0 (0.0%) | 0 (0.0%) |
| American Indian / Alaska Native | 1 (0.3%) | 1 (1.5%) | 0 (0.0%) | 0 (0.0%) | 0 (0.0%) | 0 (0.0%) | 0 (0.0%) |
| Native Hawaiian or Pacific Islander | 0 (0.0%) | 1 (1.5%) | 1 (1.3%) | 0 (0.0%) | 0 (0.0%) | 0 (0.0%) | 0 (0.0%) |
| **Ethnicity** |  |  |  |  |  |  |  |
| Hispanic or Latino | 29 (9.7%) | 8 (11.9%) | 18 (22.8%) | 3 (9.1%) | 2 (5.1%) | 1 (4.3%) | 2 (20.0%) |
| **APOE genotype** |  |  |  |  |  |  |  |
| 33 | 196 (65.3%) | 18 (26.9%) | 36 (45.6%) | 23 (69.7%) | 24 (61.5%) | 10 (43.5%) | 4 (40.0%) |
| 34 | 59 (19.7%) | 34 (50.7%) | 29 (36.7%) | 6 (18.2%) | 9 (23.1%) | 12 (52.2%) | 2 (20.0%) |
| 23 | 24 (8.0%) | 2 (3.0%) | 8 (10.1%) | 3 (9.1%) | 6 (15.4%) | 1 (4.3%) | 2 (20.0%) |
| 44 | 16 (5.3%) | 12 (17.9%) | 5 (6.3%) | 1 (3.0%) | 0 (0.0%) | 0 (0.0%) | 1 (10.0%) |
| 24 | 4 (1.3%) | 1 (1.5%) | 1 (1.3%) | 0 (0.0%) | 0 (0.0%) | 0 (0.0%) | 1 (10.0%) |
| 22 | 1 (0.3%) | 0 (0.0%) | 0 (0.0%) | 0 (0.0%) | 0 (0.0%) | 0 (0.0%) | 0 (0.0%) |
| **srGS N (%)** | 240 (80.0%) | 49 (73.1%) | 60 (75.9%) | 31 (93.9%) | 32 (82.1%) | 21 (91.3%) | 4 (40.0%) |
| **Plasma proteomics N (%)** | 275 (91.7%) | 55 (82.1%) | 63 (79.7%) | 32 (97.0%) | 37 (94.9%) | 21 (91.3%) | 7 (70.0%) |
| **Blood single-cell RNA seq N (%)** | 132 (44.0%) | 42 (62.7%) | 54 (68.4%) | 27 (81.8%) | 31 (79.5%) | 18 (78.3%) | 4 (40.0%) |
| **CSF proteomics N (%)** | 20 (6.7%) | 9 (13.4%) | 3 (3.8%) | 3 (9.1%) | 8 (20.5%) | 1 (4.3%) | 1 (10.0%) |

**Extended Data Table 2.**
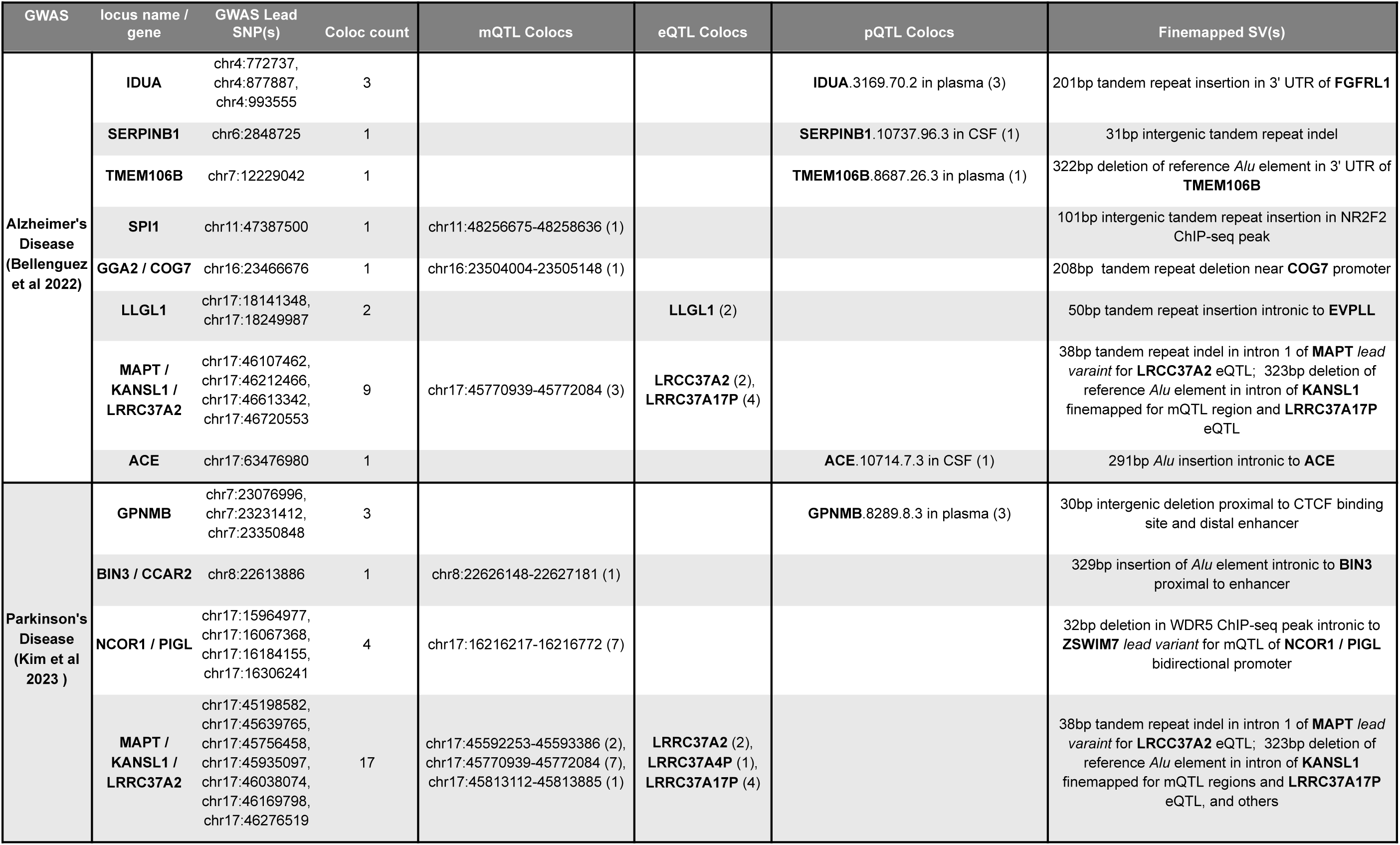
GWAS risk loci colocalized with SV-finemapped QTLs.

| GWAS | locus name / gene | GWAS Lead SNP(s) | Coloc count | mQTL Colocs | eQTL Colocs | pQTL Colocs | Finemapped SV(s) |
| --- | --- | --- | --- | --- | --- | --- | --- |
| Alzheimer's Disease<br>(Bellenguez et al 2022) | IDUA | chr4:772737,<br>chr4:877887,<br>chr4:993555 | 3 |  |  | IDUA.3169.70.2 in plasma (3) | 201bp tandem repeat insertion in 3' UTR of **FGFRL1** |
|  | SERPINB1 | chr6:2848725 | 1 |  |  | SERPINB1.10737.96.3 in CSF (1) | 31bp intergenic tandem repeat indel |
|  | TMEM106B | chr7:12229042 | 1 |  |  | TMEM106B.8687.26.3 in plasma (1) | 322bp deletion of reference *Alu* element in 3' UTR of **TMEM106B** |
|  | SPI1 | chr11:47387500 | 1 | chr11:48256675-48258636 (1) |  |  | 101bp intergenic tandem repeat insertion in NR2F2 ChIP-seq peak |
|  | GGA2 / COG7 | chr16:23466676 | 1 | chr16:23504004-23505148 (1) |  |  | 208bp tandem repeat deletion near **COG7** promoter |
|  | LLGL1 | chr17:18141348,<br>chr17:18249987 | 2 |  | LLGL1 (2) |  | 50bp tandem repeat insertion intronic to **EVPLL** |
|  | MAPT / KANSL1 / LRRC37A2 | chr17:46107462,<br>chr17:46212466,<br>chr17:46613342,<br>chr17:46720553 | 9 | chr17:45770939-45772084 (3) | LRCC37A2 (2),<br>LRRC37A17P (4) |  | 38bp tandem repeat indel in intron 1 of **MAPT** *lead variant* for **LRCC37A2** eQTL; 323bp deletion of reference *Alu* element in intron of **KANSL1** finemapped for mQTL region and **LRRC37A17P** eQTL |
|  | ACE | chr17:63476980 | 1 |  |  | ACE.10714.7.3 in CSF (1) | 291bp *Alu* insertion intronic to **ACE** |
| Parkinson's Disease<br>(Kim et al 2023) | GNPMB | chr7:23076996,<br>chr7:23231412,<br>chr7:23350848 | 3 |  |  | GNPMB.8289.8.3 in plasma (3) | 30bp intergenic deletion proximal to CTCF binding site and distal enhancer |
|  | BIN3 / CCAR2 | chr8:22613886 | 1 | chr8:22626148-22627181 (1) |  |  | 329bp insertion of *Alu* element intronic to **BIN3** proximal to enhancer |
|  | NCOR1 / PIGL | chr17:15964977,<br>chr17:16067368,<br>chr17:16184155,<br>chr17:16306241 | 4 | chr17:16216217-16216772 (7) |  |  | 32bp deletion in WDR5 ChIP-seq peak intronic to **ZSWIM7** *lead variant* for mQTL of **NCOR1** / **PIGL** bidirectional promoter |
|  | MAPT / KANSL1 / LRRC37A2 | chr17:45198582,<br>chr17:45639765,<br>chr17:45756458,<br>chr17:45935097,<br>chr17:46038074,<br>chr17:46169798,<br>chr17:46276519 | 17 | chr17:45592253-45593386 (2),<br>chr17:45770939-45772084 (7),<br>chr17:45813112-45813885 (1) | LRRC37A2 (2),<br>LRRC37A4P (1),<br>LRRC37A17P (4) |  | 38bp tandem repeat indel in intron 1 of **MAPT** *lead variant* for **LRCC37A2** eQTL; 323bp deletion of reference *Alu* element in intron of **KANSL1** finemapped for mQTL regions and **LRRC37A17P** eQTL, and others |

## Extended Data and Supplemental Figures

**Extended Data Figure 1.**
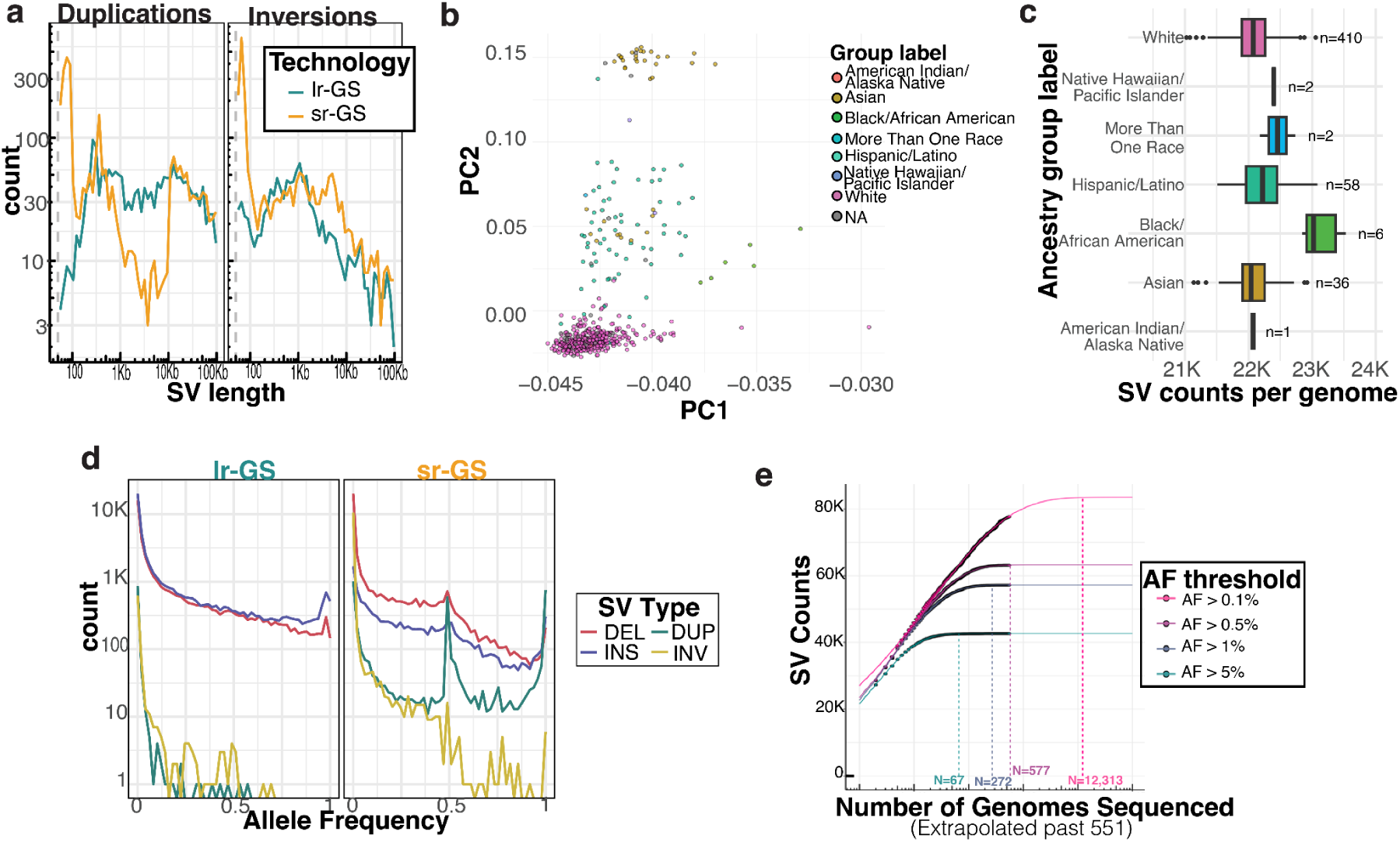
**a,** count of duplications and inversions in 366 matched lrGS and srGS samples stratified by length **b,** principal component of common variant SV genotypes (MAF>5%) showing population stratification, colored by ancestry group. Ancestry group determined from demographic questionnaire surveying participant’s self-reported race and ethnicity **c,** counts of SVs per genome stratified by ancestry group, **d,** count of SVs across frequency spectrum in lrGS compared to srGS **e,** counts of SVs above various MAF thresholds detected as a function of number of genomes sequenced. Counts for each MAF threshold were fit to a Weibull growth model to estimate the asymptote, and the model was used to extrapolate beyond ADRC genomes and find saturation points (the number of genomes needed to acheive 99.9% the value of the predicted asymptote).

**Extended Data Figure 2.**
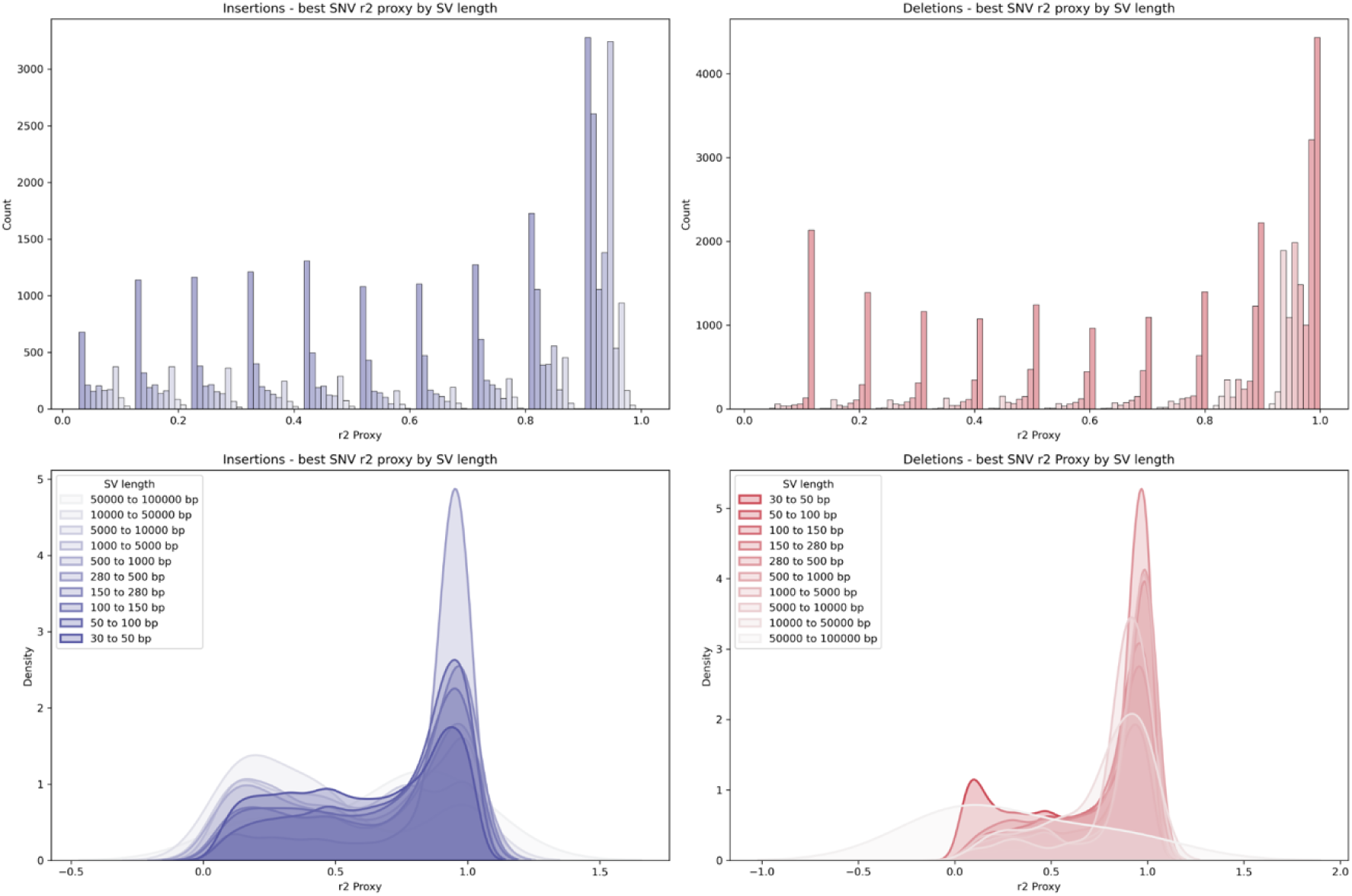
Distribution of maximum linkage disequilibrium (r²) between common structural variants (MAF >2%) and nearby single-nucleotide variants (SNVs) within 1 Mb, stratified by SV type (insertions vs. deletions) and length. Short SVs (<150 bp), particularly insertions, exhibit low r² values, indicating poor tagging by adjacent SNVs, while mid-sized SVs (150–1000 bp) show improved tagging efficiency.

**Extended Data Figure 3.**
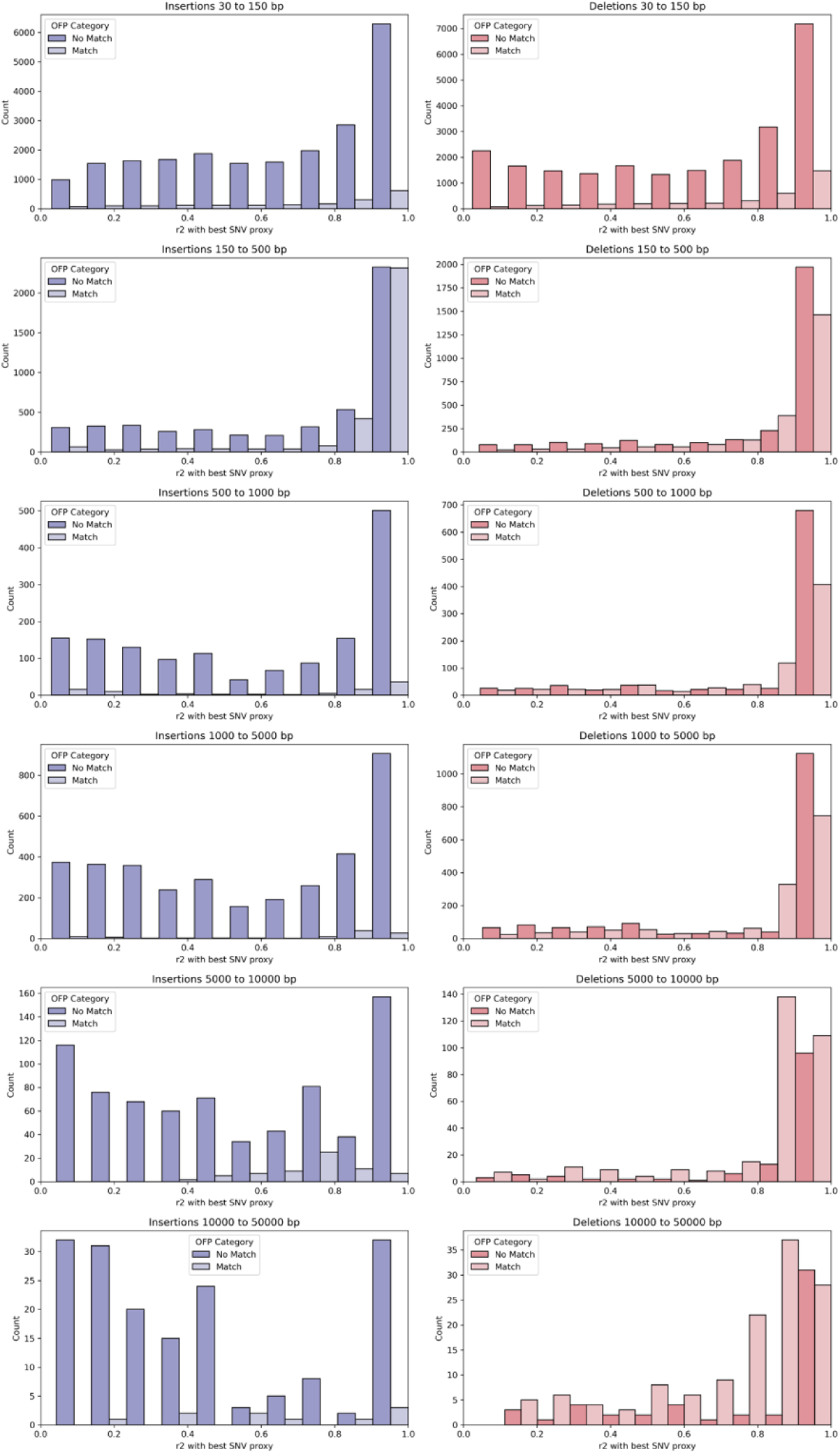
Relationship between SV overlap with gnomAD v4 short-read SVs (overlap fraction, OFP) and maximum r² with nearby SNVs, stratified by SV length. SVs with higher OFP values tend to have stronger LD with adjacent SNVs, with this trend being more pronounced in mid-sized insertions and deletions.

**Extended Data Figure 4.**
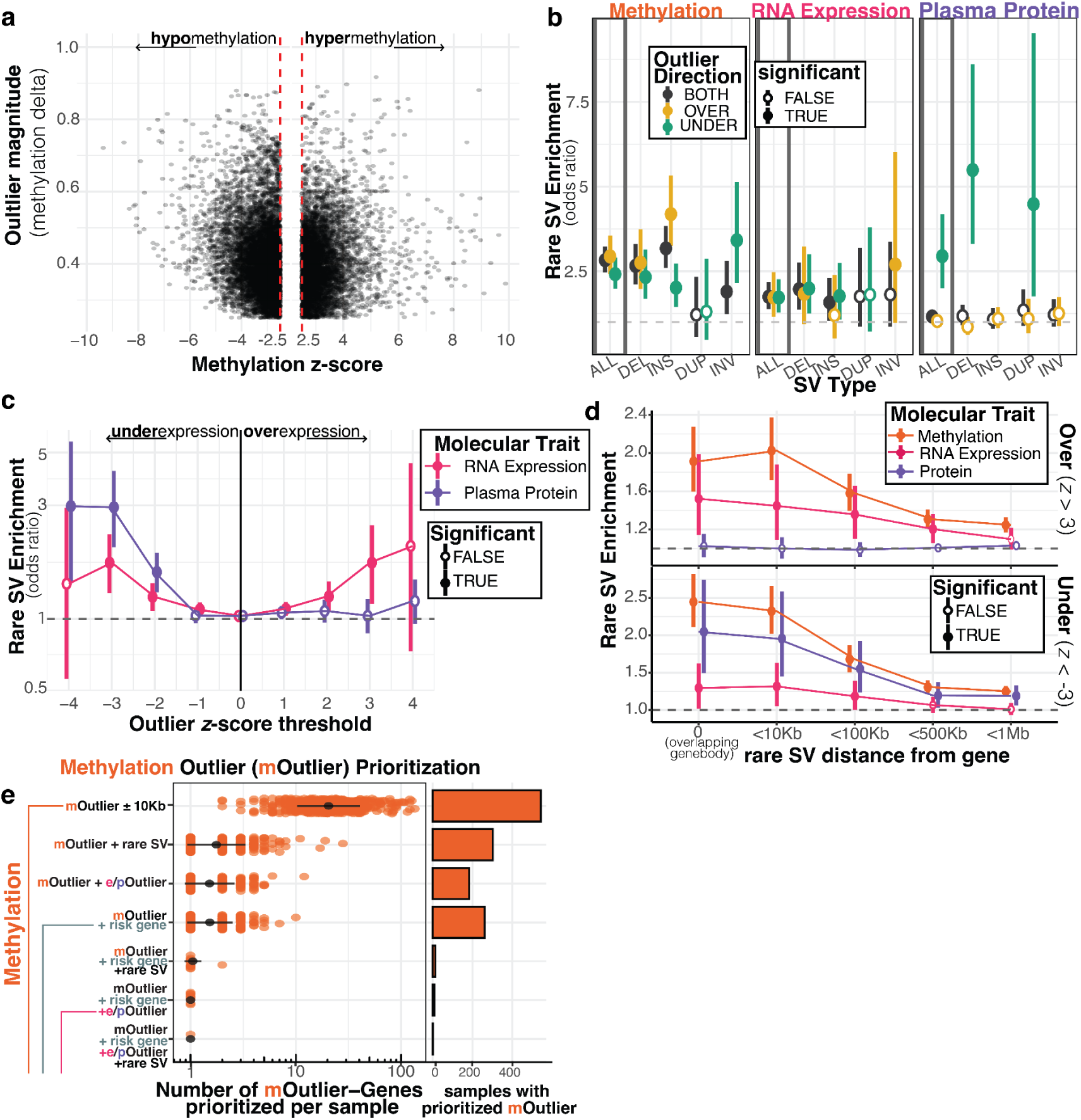
**a,** Methylation outliers called from METAFORA stratified by z-score and methylation delta (absolute difference of observed outlier from population median). **b,** SV-specific enrichment of rare variants near outliers (z-threshold=3) across 3 different molecular traits. Values not reported for examples where fewer than 5 rare variants were observed near outliers. **c,** rare variant enrichment for RNA and protein outliers as a function of z-score threshold used to define outliers. Positive z-score threshold for over-expression (outlier = z > z_threshold), negative z-scores for under-expression (outlier = z < z_threshold). **d,** rare variant enrichment for outliers across molecular traits as a function of distance to the outlier gene. Distance of 0 corresponds to SV overlapping the gene body. **e** Prioritization of methylation outlier (mOutliers) regions per sample. A dot plot displays the number of methylation outliers detected per sample, matching corresponding filters on the y-axis. Filters indicate if rare SV is present within 10kb from mOutlier (rare SV), if gene near mOutlier is a molecular outlier in another ‘ome (ie expression or protein) (e/pOutlier), or if gene near mOutlier has colocalization evidence for AD or PD risk (risk gene). Bar plot displays the number of samples with at least one methylation outlier meeting those filters.

**Extended Data Figure 5.**
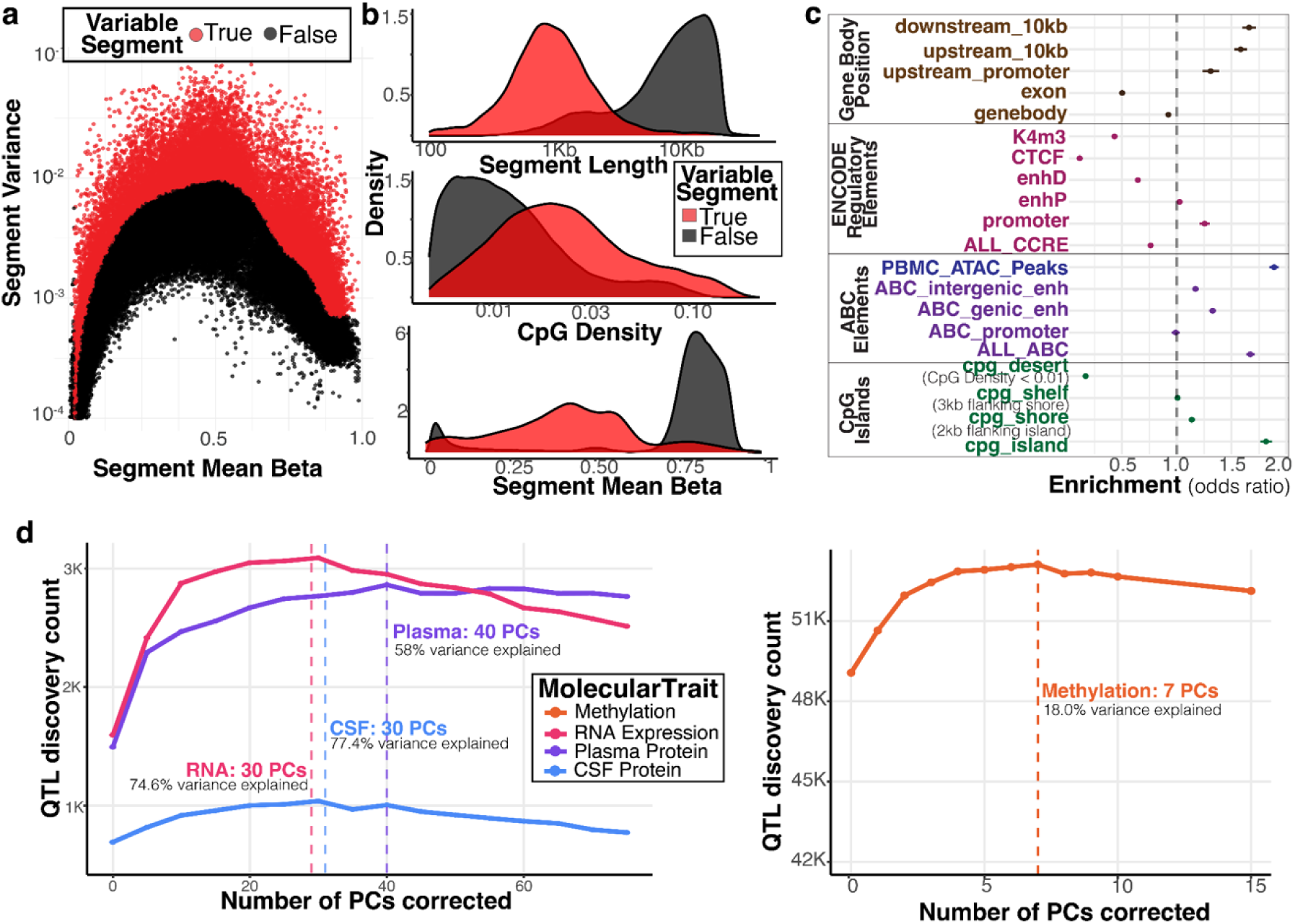
**a,** mean methylation betas and variance of 360K methylation regions detected from segmenting population mean methylation beta profile. Segments in red are defined as variable after modeling the mean-variance relationship across segments. Variable segments have variance above 3 standard errors of predicted variance, given that segments’ mean beta. **b,** distribution of variable methylation segments compared to background segments across 3 different metrics, length of segment (base pairs), CpG density (number of cpg sites per 100 bases), segment mean beta (mean proportion of methylated bases across all CpGs in segment). **c,** enrichment of various genome annotations pulled from ENCODE, ABC models, UCSC cpg island track, and gencode v32 gene coordinates for overlapping variable segments. **d,** number of QTLs detected at an FDR threshold of 5% for a variety of QTL models correcting for an increasing number of phenotype PCs across the 4 molecular traits. The number of PCs achieving a maximum of QTL discoveries is shown with a vertical line, and the percent of variance explained by those PCs is also annotated.

**Extended Data Figure 6.**
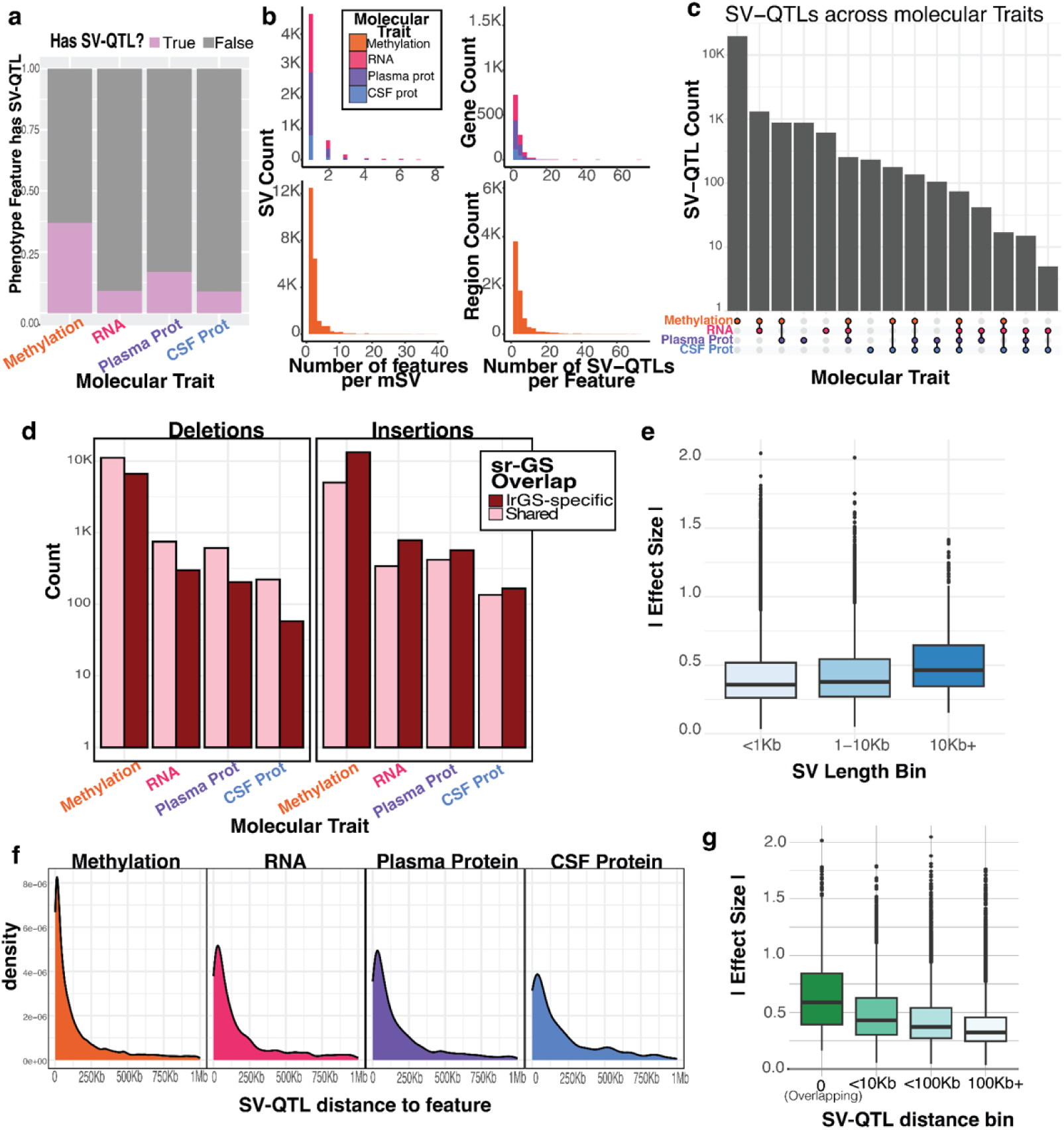
**a,** percent of molecular trait features (genes for expression, SomaLogic aptamers for plasma/csf protein, and variable methylation segments for Methylation) with a detected SV-QTL. **b**, histogram showing the distribution of both the number of features associated with each SV and the number of SVs associated with each feature. Methylation is plotted on a separate axis for clarity, as it has higher counts. **c**, upset plot of the sharing of QTLs across molecular traits for eSVs. Each unique SVID was overlapped across all significant SV-QTLs for each molecular trait. **d**, count of SV-QTLs where the SV was uniquely detected by lr-GS, or also present in sr-GS-based population resources like gnomAD v4. **e**, absolute effect size distribution of SV-QTLs in different length bins shows that larger variants have a higher effect. **f**, distribution of SV-QTLs based on the distance to their target feature (TSS for RNA and protein, and methylation region for Methylation). Methylation had significantly closer associations compared to other omes. **g**, absolute effect size distribution of SV-QTLs binned by how close they are to the target feature. 0 represents an SV that directly overlapped the target feature

**Extended Data Figure 7.**
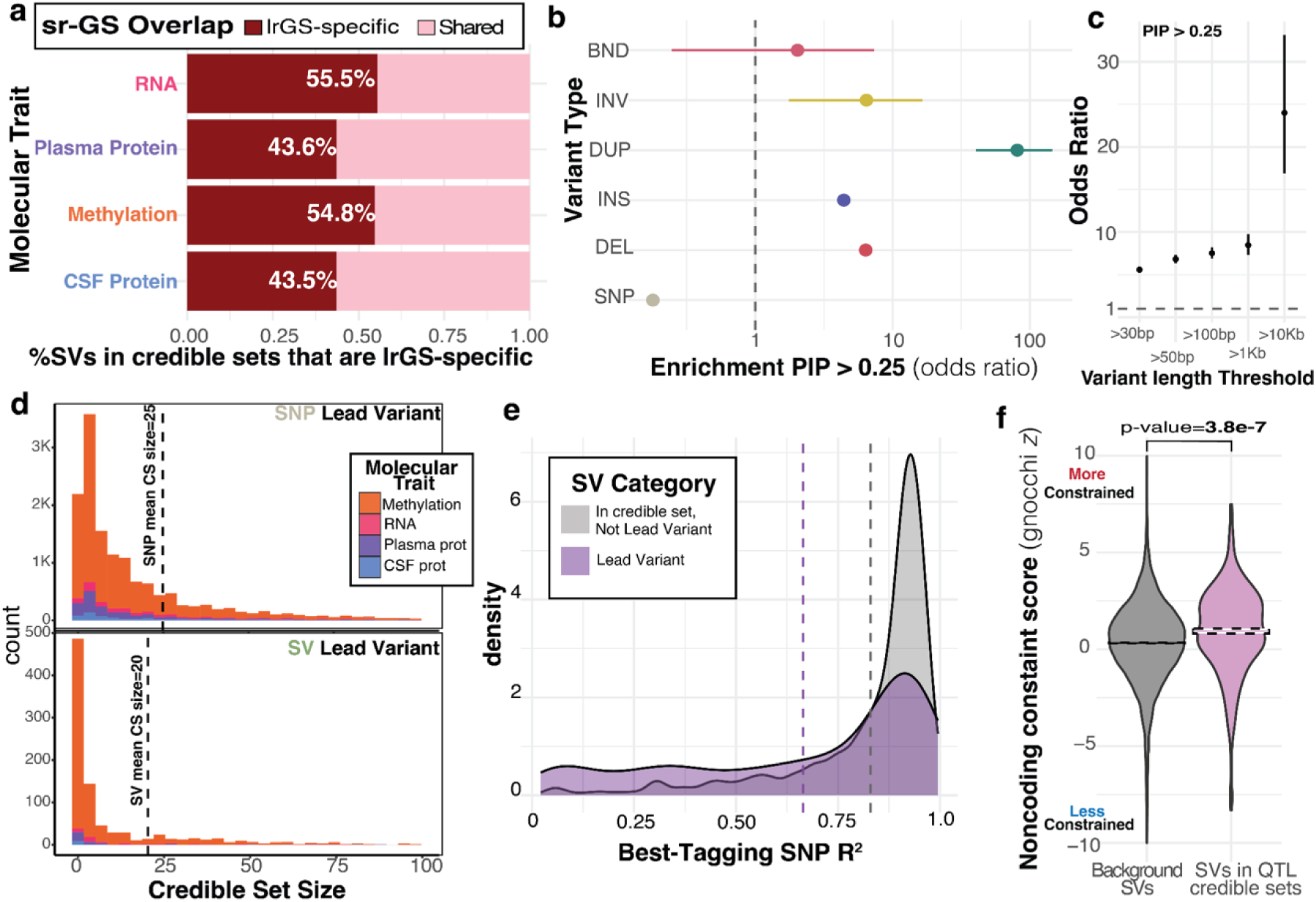
**a,** percent of SVs contained in QTL credible sets that were lrGS-specific (absent from sr-GS based population references) **b,** enrichment across variant types for being fine-mapped with PIP above 0.25. Odds ratio from fisher’s exact test plotted on x-axis **c,** enrichment for variants above various length thresholds being fine-mapped with PIP above 0.25 **d**, number of variants included in credible sets stratified by whether lead variant of credible set (highest pip variant) was a SNP or SV **e,** distribution of the LD of the best-tagging SNP for lead SVs compared to background SV-QTLs **f,** distribution of max non-coding constraint score (gnocchi score) for regions overlapping SV, comparing fine-mapped SVs to all background SVs.

**Extended Data Figure 8.**
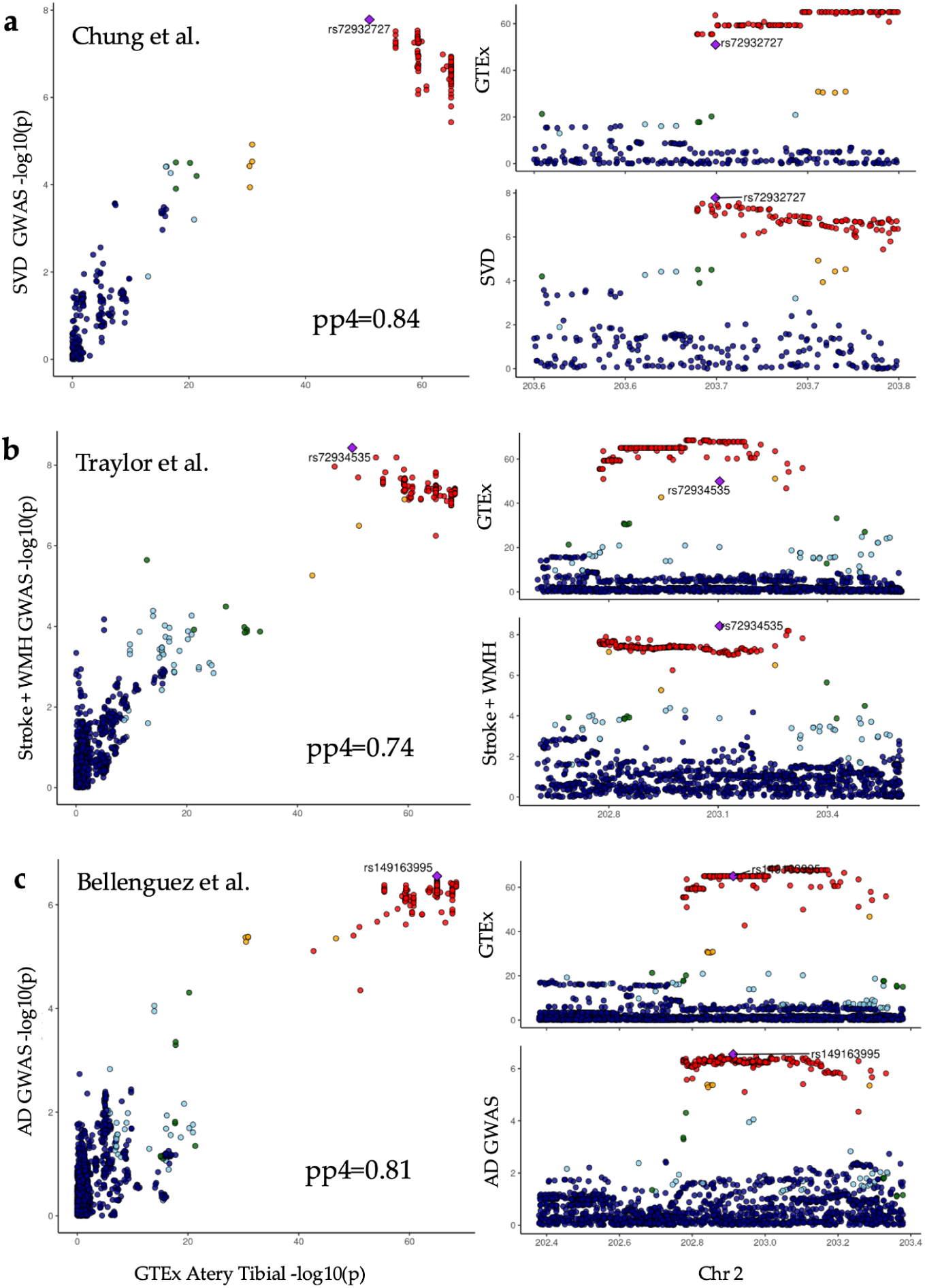
Colocalization of *NBEAL1* expression in GTEx Artery Tibial with Alzheimer’s disease and neurovascular GWAS loci. Colocalization plots showing *NBEAL1* expression in GTEx Artery Tibial and GWAS signals for: **a,)** Small vessel disease (SVD) (Chung et al., 2022 PMID: 31430377), **b,** Stroke and white matter hyperintensities (WMH) (Traylor et al., 2021, PMID: 33773637), **c,** Alzheimer’s disease (AD) (Bellenguez et al., 2024,PMID: 35379992). This locus includes a large intronic deletion in *NBEAL1* (chr2:203034349–203039584; 5,235 bp), in high LD with SNPs previously associated with AD (rs139643391, *WDR12*, R²=0.92), SVD (rs72932727, *ICA1L*, R²=0.89), ischemic stroke, and WMH (rs72934535, *NBEAL1*, R²=0.82). Posterior probabilities of colocalization (pp4) are shown: SVD (PP4 = 0.83), WMH (PP4 = 0.74), and AD (PP4 = 0.81). Lead GWAS SNPs are labeled in the GTEx and GWAS SNPs intersection.

**Supplementary Figure 1.**
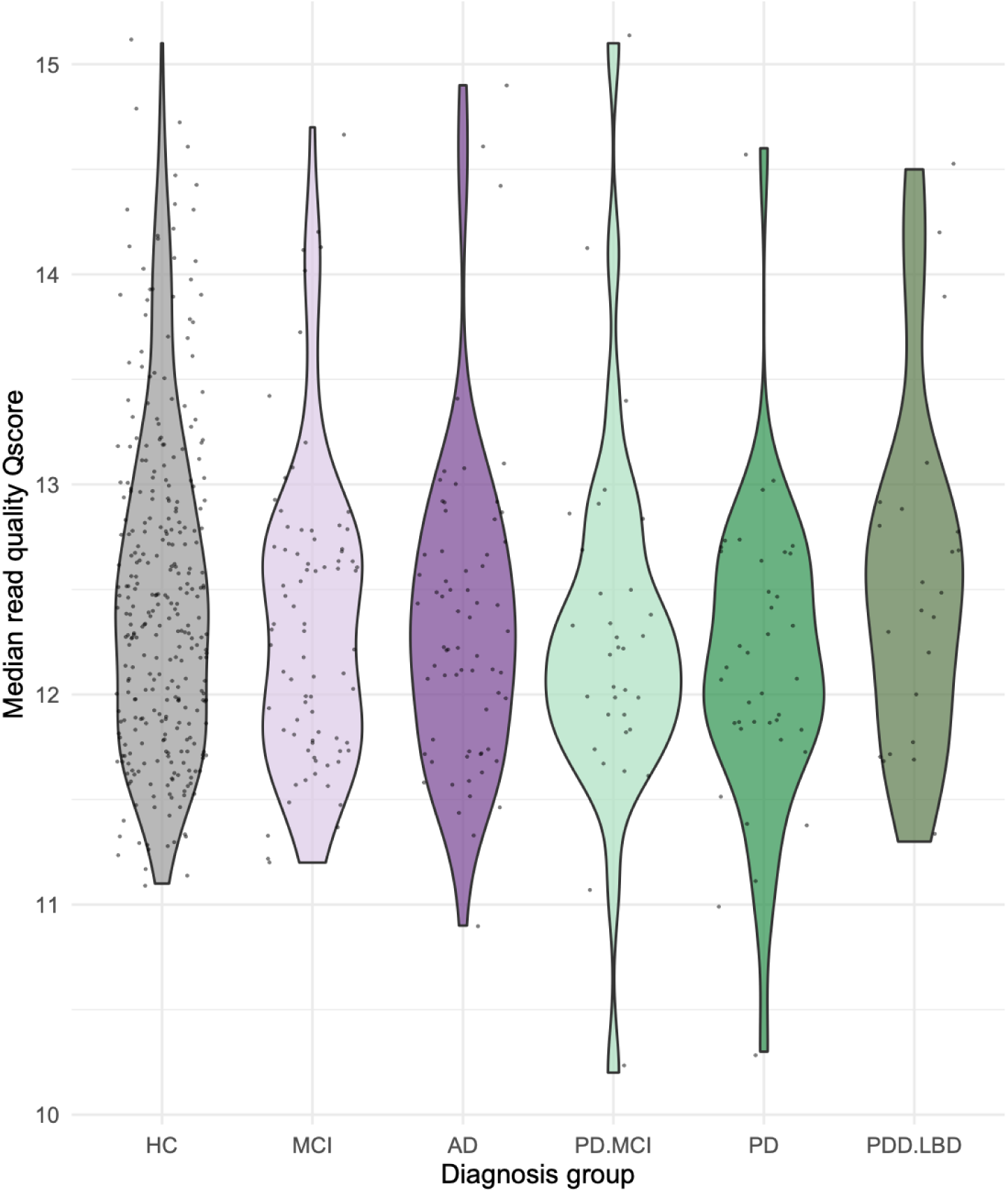
Distribution of median read quality score (Qscore) across all passed reads (Qscore > 10) for ADRC and SAMs participants, stratified by diagnosis group. ONT sequencing on R9 flow cells achieved a median sample Qscore of 12.3, corresponding to 93.6% identity.

**Supplementary Figure 2.**
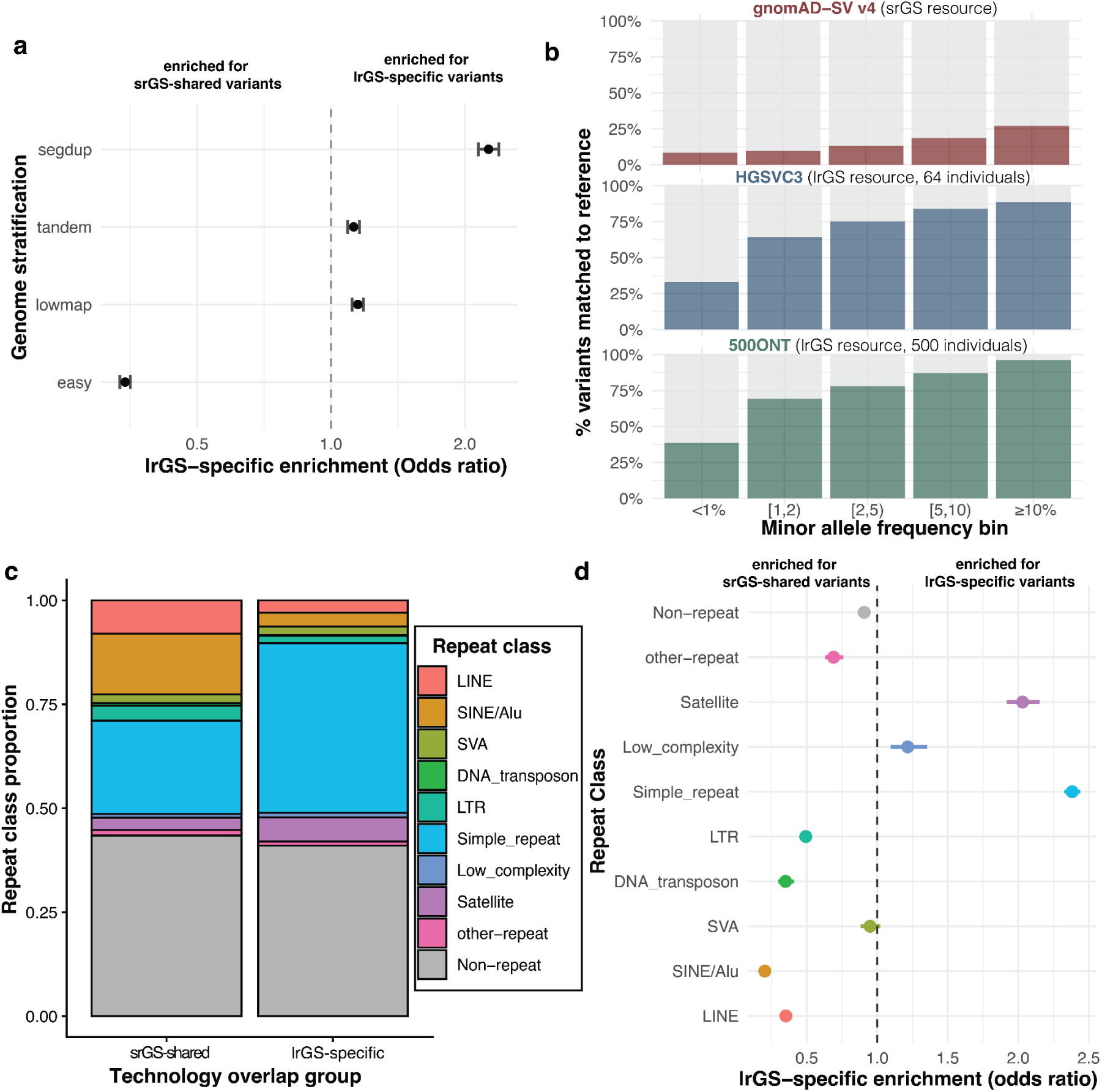
**a,** enrichment of lrGS-specific vs srGS-shared variants across GIAB genome stratifications. Easy refers to “not in difficult regions,” ie not in low mappability, low complexity, or low copy repeat regions. **b,** percent of lrGS SVs in this study that match (reciprocal overlap ≥ 50%) an SV of the same type in three different population SV databases, stratified by minor allele frequency of variant. HGSVC3 is recent release of Human Genome Structural Variation Consortia (Logsdon et al 2025), and 500ONT is first 500 ONT genomes sequenced as part of 1000G Long-read Sequencing Consortia (Gustafson et al 2024). **c,** RepeatMasker defined classes of repetitive insertion/deletion sequences stratified by whether variant was lrGS-specific or shared with srGS. **d,** enrichment of lrGS-specific variants compared to shared variants for different repeat classes.

**Supplementary figure 3.**
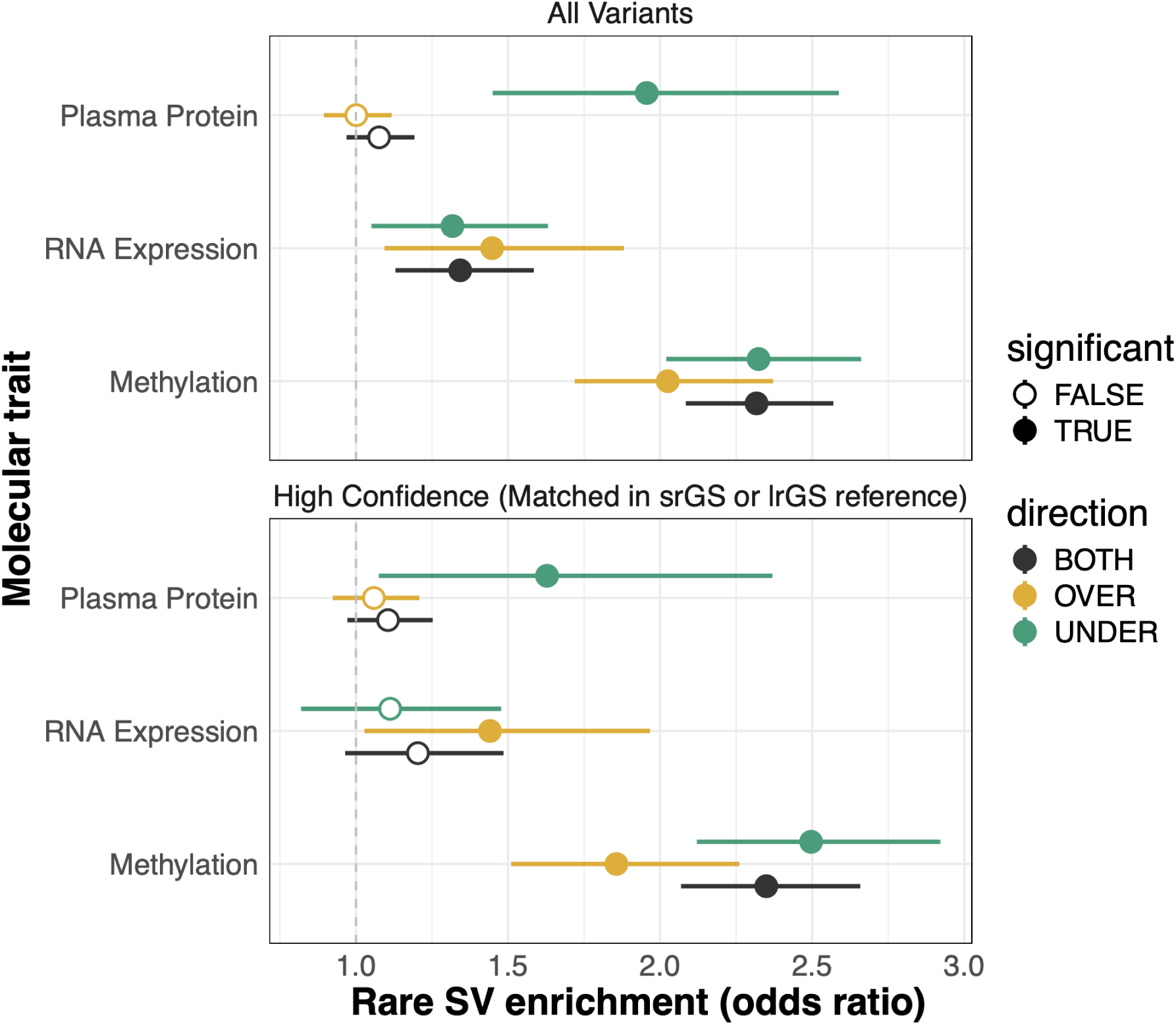
Enrichment of high-confidence rare SVs (those supported by orthogonal evidence) for molecular outliers, compared to enrichment for all rare SVs. High-confidence SVs are defined as those that were observed either in the short-read srGS callset or match a variant seen before in a population SV database (gnomAD-SV v4, HGSVC3, or 500 genomes from 1000G Long-read sequencing consortium). We generally observe more modest enrichments for high-confidence SVs likely due to the rarest SVs–those that typically are most enriched for outliers–being largely absent from population resources.

**Supplementary Figure 4.**
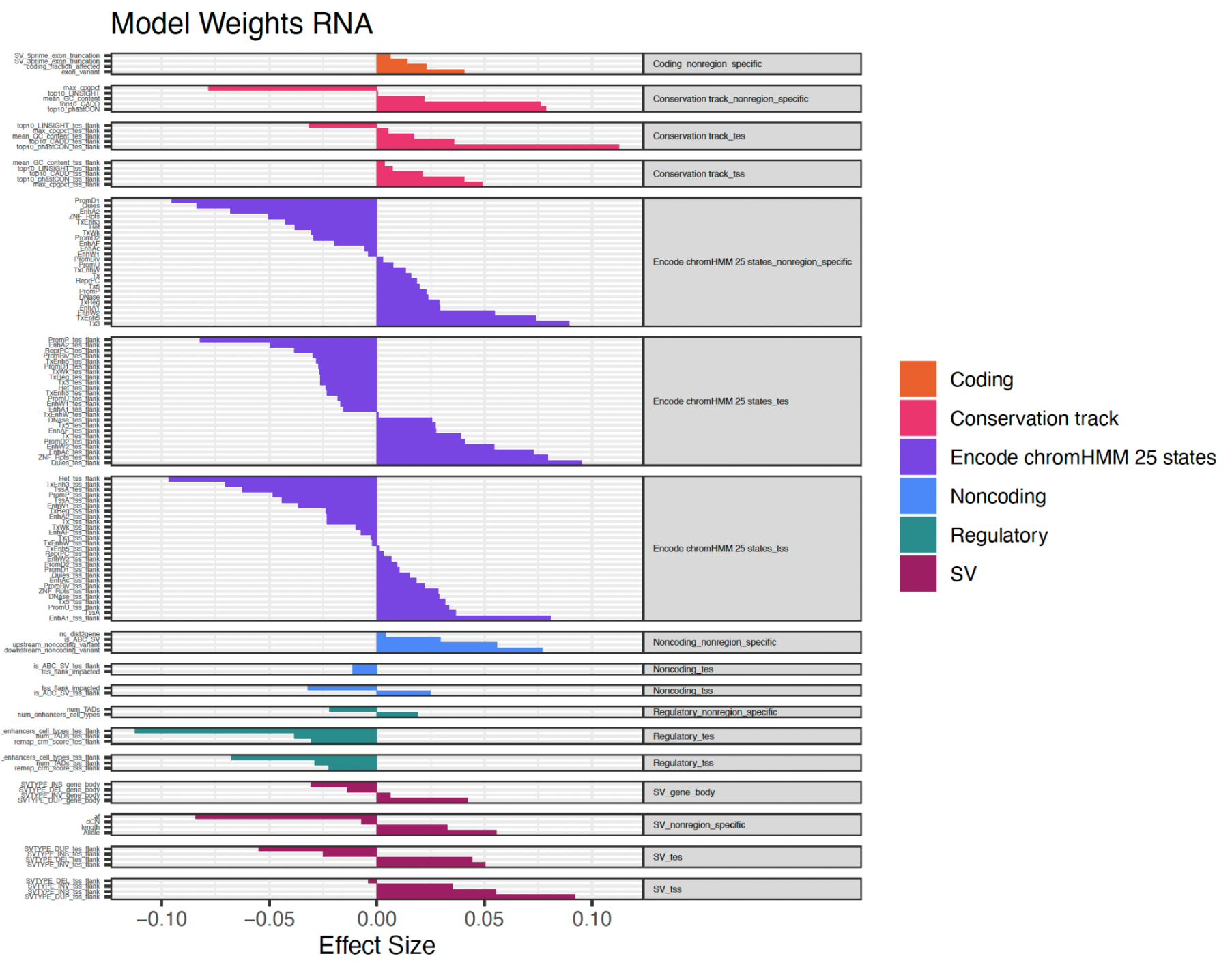
Watershed model weights for RNA expression data were stratified by both genomic regions (including gene body, transcription start site (TSS), and transcription end site (TES)) and gene annotation categories. These categories included: coding (non-region specific), noncoding (non-region specific), regulatory (non-region specific), conservation track (non-region specific), ENCODE ChromHMM25 states (non-region specific), and structural variants (SVs) located within the gene body. The strongest expression-related annotations were: (1) structural variants (SVs) located within exon regions, (2) SVs in proximity to genes that intersect an ABC enhancer assigned to the corresponding gene, and (3) the number of primary tissues in which an SV within an enhancer BED segment is observed.

**Supplementary Figure 5.**
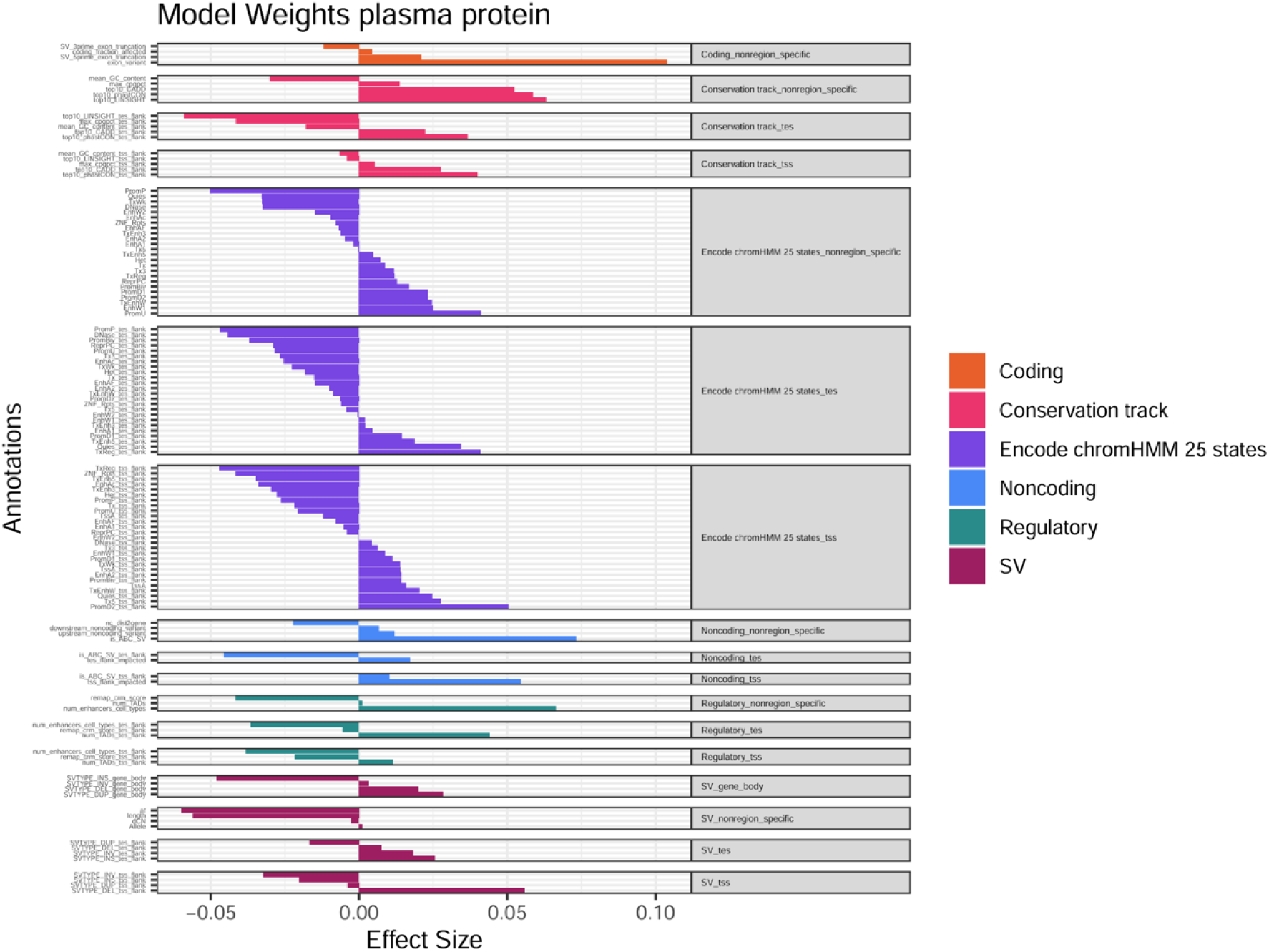
For plasma protein, the top three annotation levels included: (1) SVs situated in exon regions, (2) SVs near genes overlapping with ABC enhancers linked to the gene, and (3) the mean of the top 10 LINSIGHT conservation scores across the SV span.

**Supplementary Figure 6.**
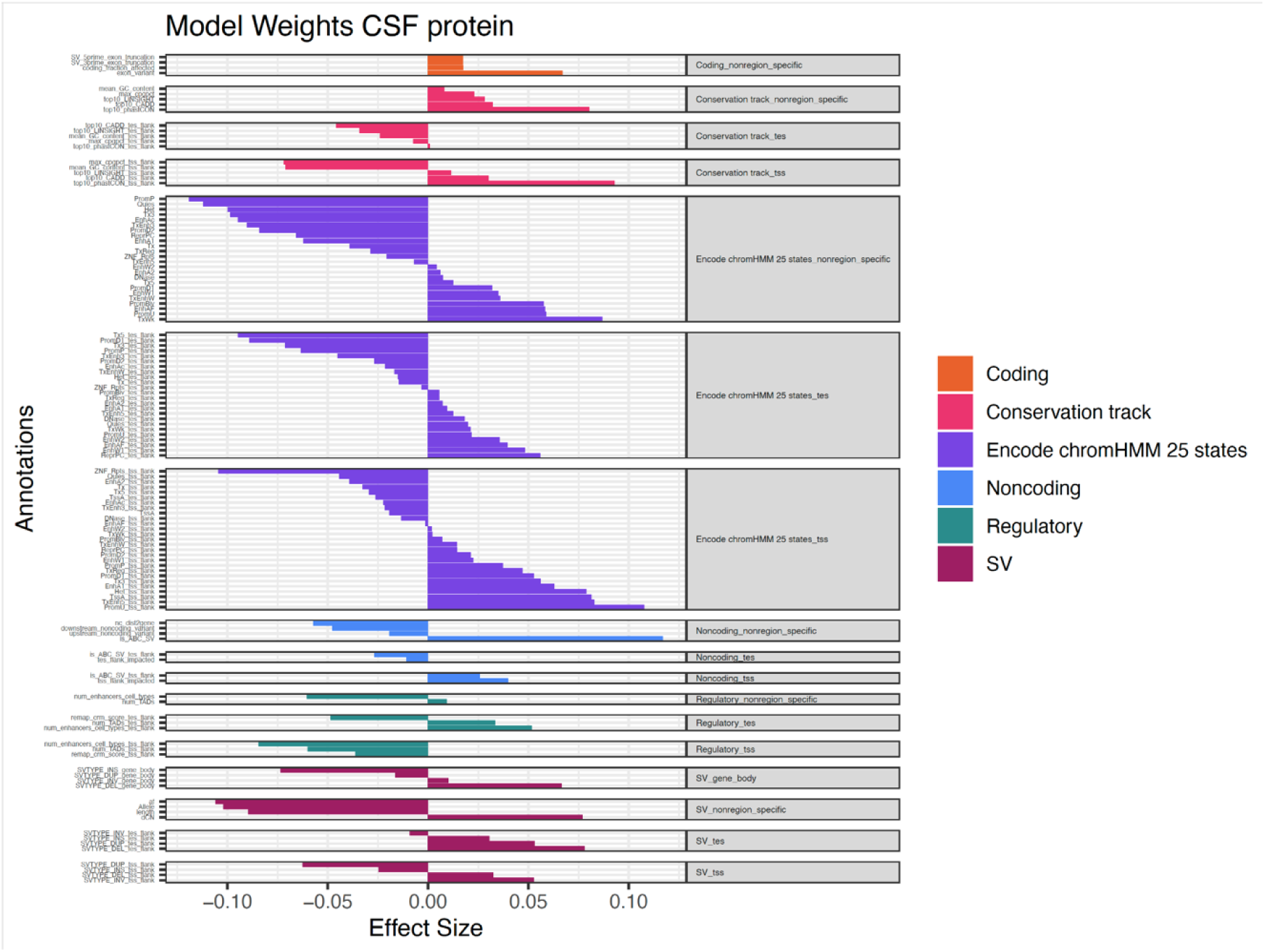
The top annotations for CSF protein levels were: (1) SVs near genes that overlap ABC enhancers mapped to the gene, (2) SVs in the upstream promoter-flanking region of a transcription start site (TSS), and (3) the average of the top 10 phastCons conservation scores across the SV region.

**Supplementary Figure 7.**
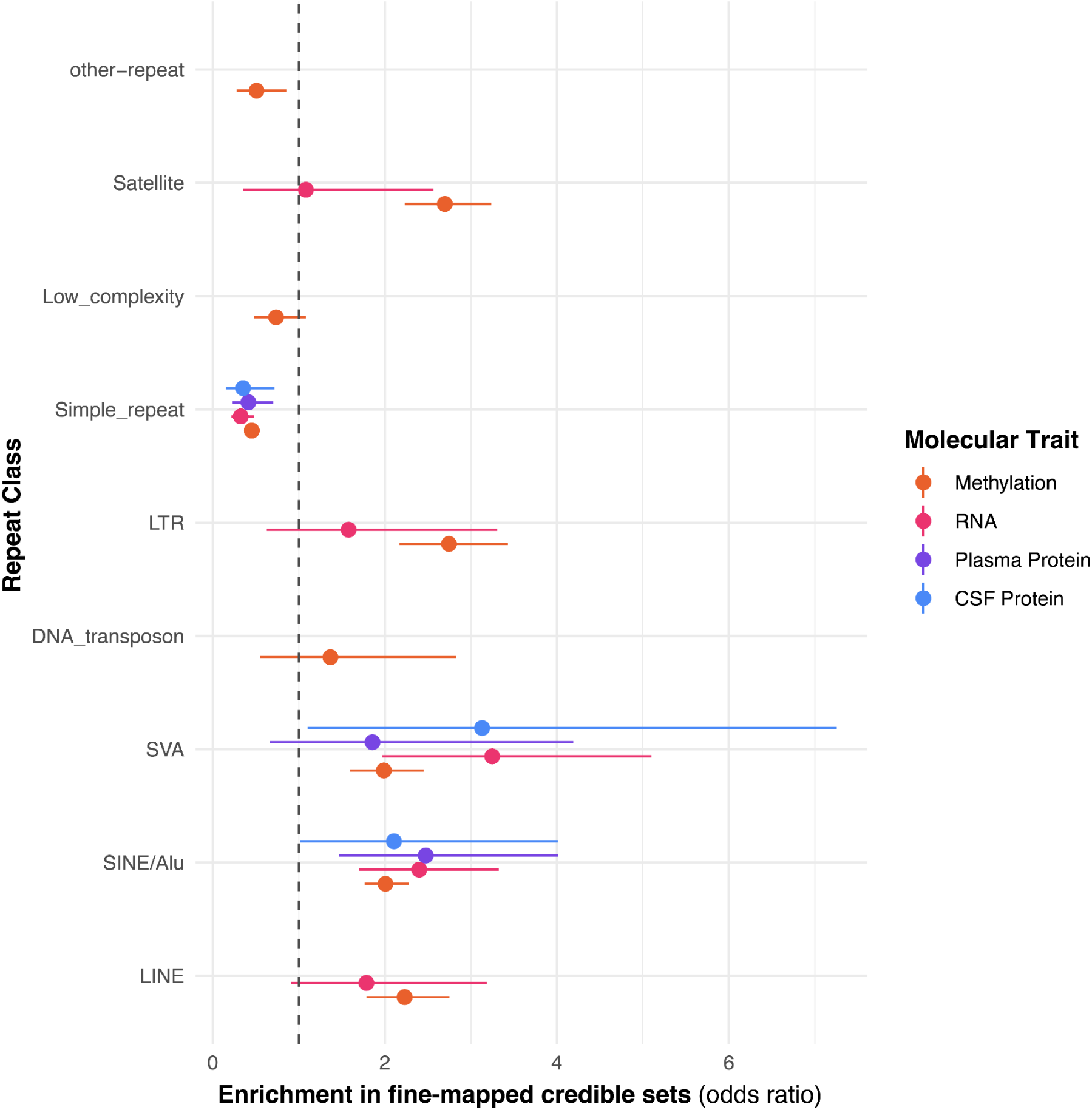
Enrichment of SVs of different repeat classes as defined by RepeatMasker in QTL credible sets across 4 different molecular traits. X-axis plots fisher’s exact test odds ratio estimate and confidence interval. Enrichment not calculated for repeats where there were fewer than 5 examples fine-mapped for that ‘ome.

**Supplementary Figure 8.**
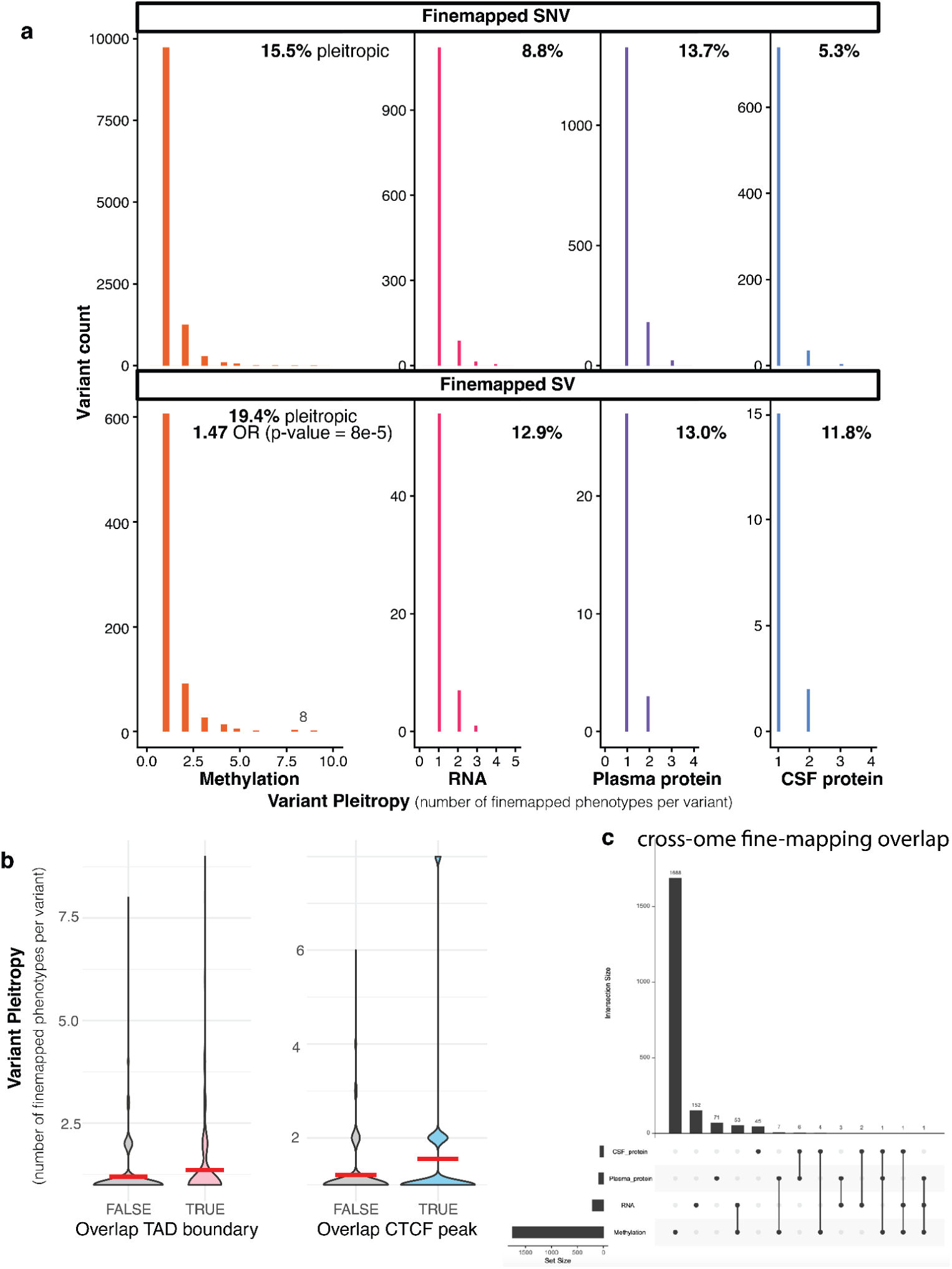
**a,** Distributions of the number of finemapped phenotypes per variant for finemapped SNVs vs SVs across 4 molecular traits. Percent of pleiotropic variants (variants with more than 1 finemapped phenotype) annotated in top right corner. **b,** Distribution of the number of finemapped phenotypes per SV stratified by whether variant overlaps lymphoblastoid cell line TAD boundaries from Hi-C data (Rao et al 2014) or CTCF binding ChIP-seq peak (ENCODE: ENCFF017XLW). Red cross bar represents the group mean. **c,** upset plot displaying SV overlap across the 4 molecular traits for finemapped SVs

**Supplementary Figure 9.**
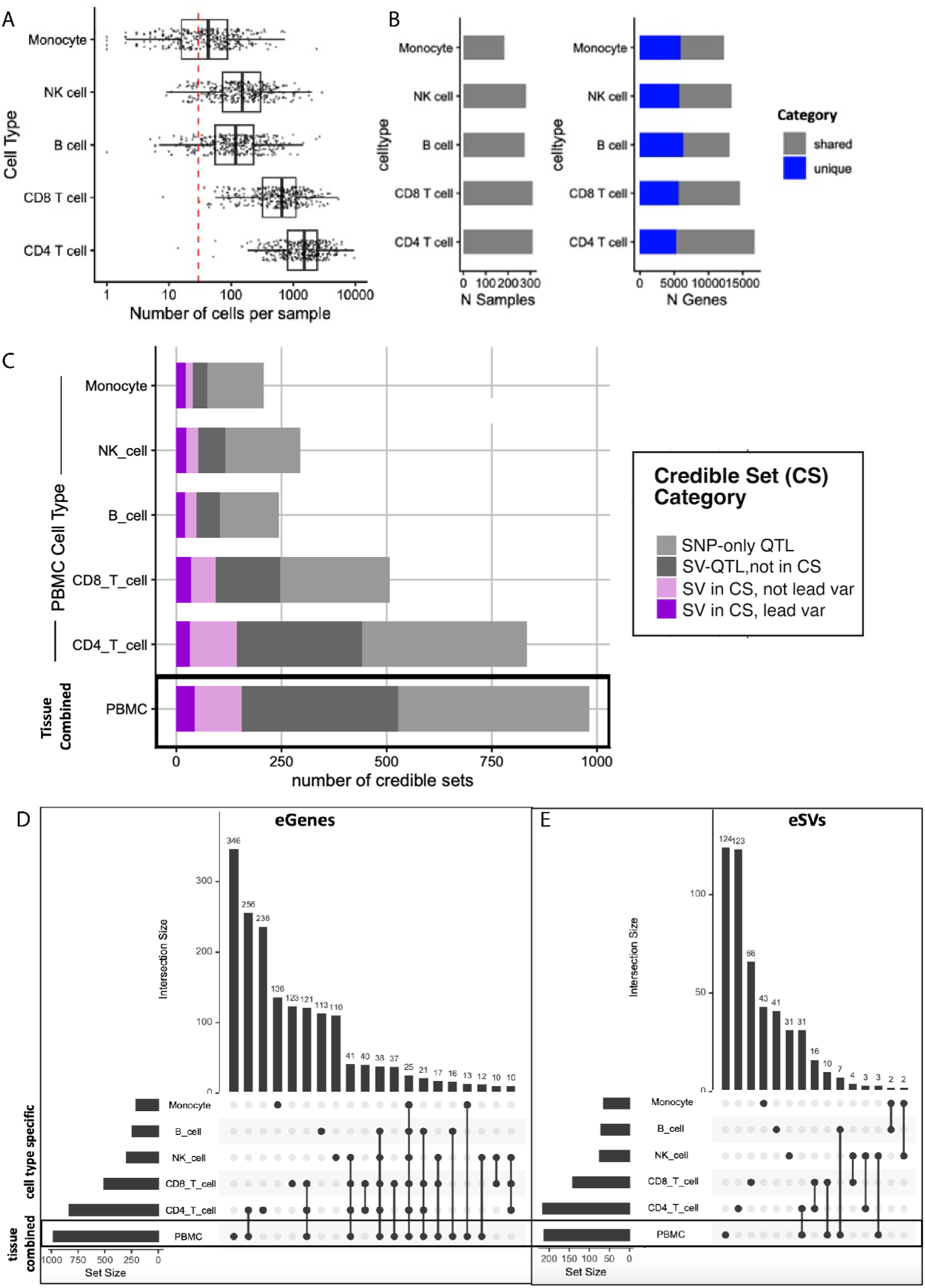
**a,** Count of cells per sample across the 5 most abundant PBMC cell types in scRNA-seq data. Red line represents the minimum number of 30 cells for sample inclusion in psuedobulk aggregation. **b,** Count of samples with at least 30 cells per cell type, and count of genes meeting variance and expression thresholds in that cell type. Genes are colored blue if they are cell-type-unique, ie if they didnt meet inclusion criterion in the bulk tissue aggregation. **c,** Number of credible sets identified from SuSiE finemapping in bulk tissue profile compared to cell-type-specific profiles. Credible sets colored by whether SV was included in credible set and if that SV was the highest pip lead variant. **d,e** Upset plot displaying overlap of across bulk tissue combined profile and cell-type specific profiles. **c** displays overlap of eGenes (genes with at least one credible set finemapped by SuSiE) while **d** displays overlap of eSVs (SVs included in at least one finemapped credible set).

**Supplementary Figure 10.**
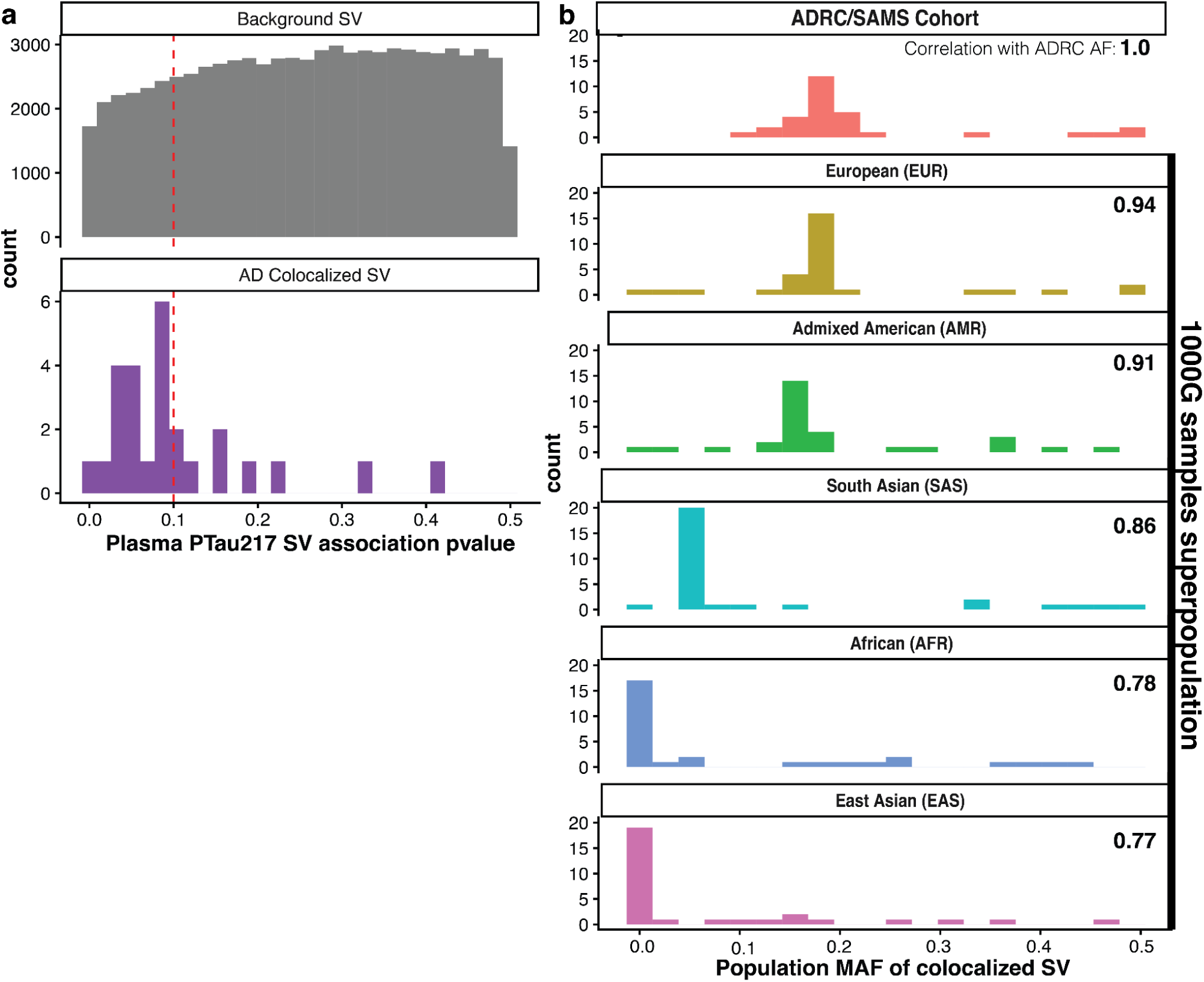
**a,** P-value distribution from linear model of SV genotype on Plasma PTau217 levels, an Alzheimer’s biomarker that captures brain amyloid pathology, stratified by whether SV had colocalization evidence for AD GWAS. Linear model estimates effect of SV genotype on PTau217 levels controlling for age, sex, and APOE genotype. AD Colocalized SVs were 10 times more likely to be significant at alpha threshold of 0.1 (red dashed line) compared to background SVs. **b,** Distribution of minor allele frequencies (MAF) of colocalized SVs as in the ADRC/SAMs cohort (top row), and as ascertained in 5 superpopulations from 1000G samples. AFs were ascertained in 1000G using 500 ONT genomes from the 1000G Long-read sequencing consortium (Gustafson et al 2024). Correlation of population-specific SV AFs to AFs from our ADRC/SAMs samples reported in the top right corner of each histogram, and populations are ordered according to this correlation. Notably, many colocalized SVs with higher frcequency in EUR superpopulation are absent or low frequency in other superpopulations.

**Supplementary Figure 11.**
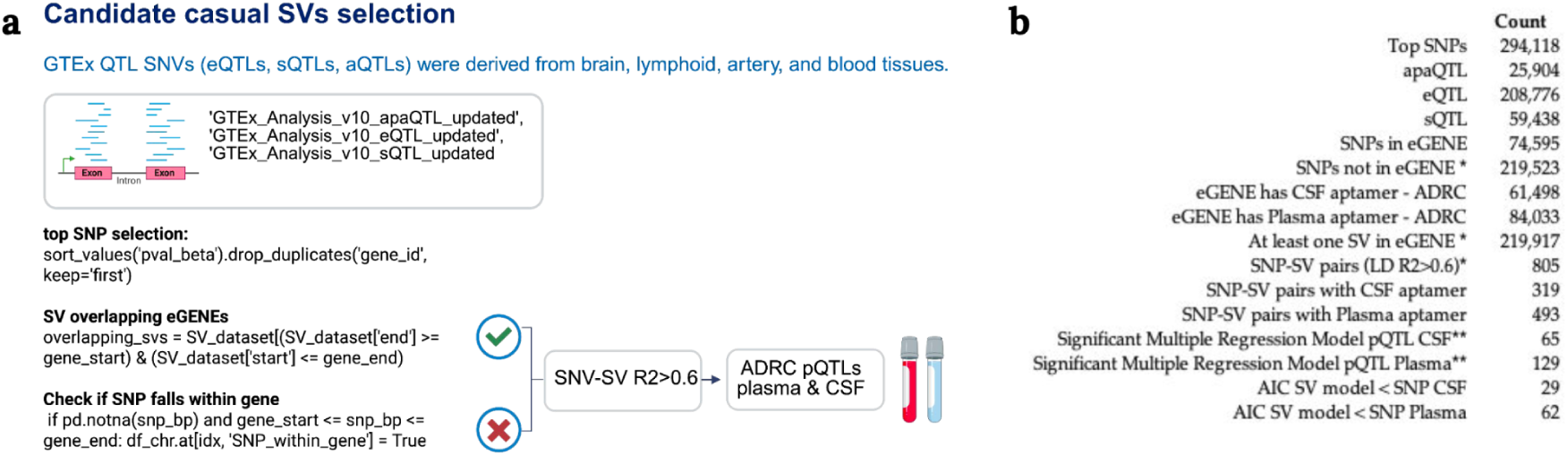
Identification of candidate causal SVs through GTEx QTL colocalization and ADRC proteomic prediction. **a,** Schematic workflow for selecting structural variants (SVs) potentially involved in gene regulation. GTEx QTL datasets—eQTLs (expression QTLs), sQTLs (splicing QTLs), and aQTLs (alternative polyadenylation QTLs)—were derived from brain, lymphoid, artery, and blood tissues. For each QTL type, the top SNP per gene was selected by sorting on p-values. SVs overlapping eGENEs were retained based on genomic coordinates. SNP–SV pairs in high linkage disequilibrium (LD R² > 0.6) were then tested in the ADRC cohort for associations with CSF and plasma protein levels (pQTLs), specifically when the top SNP did not fall within the eGENE body. **b,** Summary of the number of SNPs, QTLs, and SV–gene overlaps at each step of the pipeline. Counts include eGENEs with matching CSF or plasma aptamers in ADRC, and SNP–SV pairs with LD R² > 0.6. Asterisk (*) indicates LD calculation between SNP and SV. Double asterisk (**) indicates that either the SV or SNP showed significance in a multiple regression model (p-value < 0.05). Model fit was evaluated using Akaike Information Criterion (AIC), with a lower AIC indicating better model performance.

**Supplementary Figure 12.**
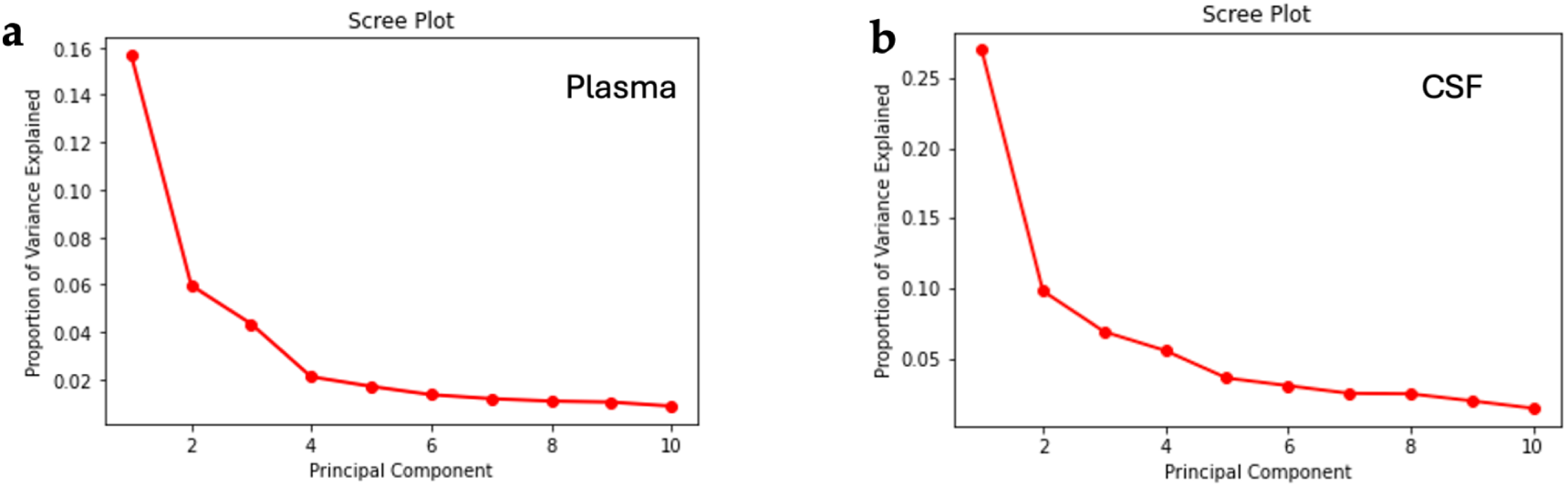
Principal components of ADRC proteomics for GTEx loci multiple regression models. a, Elbow plots showing the principal components derived from plasma proteomics and b, cerebrospinal fluid (CSF) in the ADRC cohort. For the eGENE–proteomics analyses, four principal components were included in the plasma models and six in the CSF models.

**Supplementary Figure 13.**
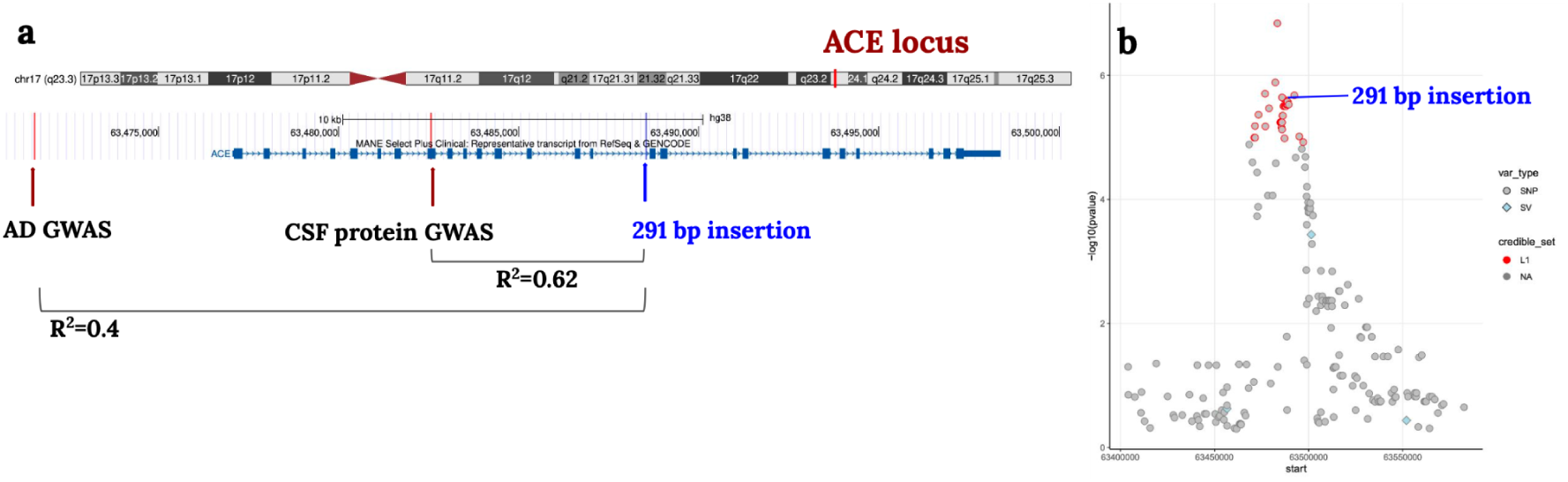
Fine-mapping of the *ACE* Locus for ADRC CSF Proteomics. **a,** Genomic view of the *ACE* locus showing the AD GWAS SNP (rs4277405, Bellenguez et al., 2024), the CSF protein GWAS SNP (rs4309, Cruchaga et al., 2023), and a 291 bp Alu insertion (chr17:63488532). Linkage disequilibrium values (R²) are indicated. **b,** Fine-mapping in the ADRC cohort for *ACE* CSF proteomics (*ACE.10714.7.3.ENSG00000159640.ACE* SomaScan aptamer): –log10(p-values) plotted by genomic position. SNPs are shown as circles, SVs as diamonds. Variants in the L1 credible set are highlighted in red, including the 291 bp insertion.

**Supplementary Figure 14.**
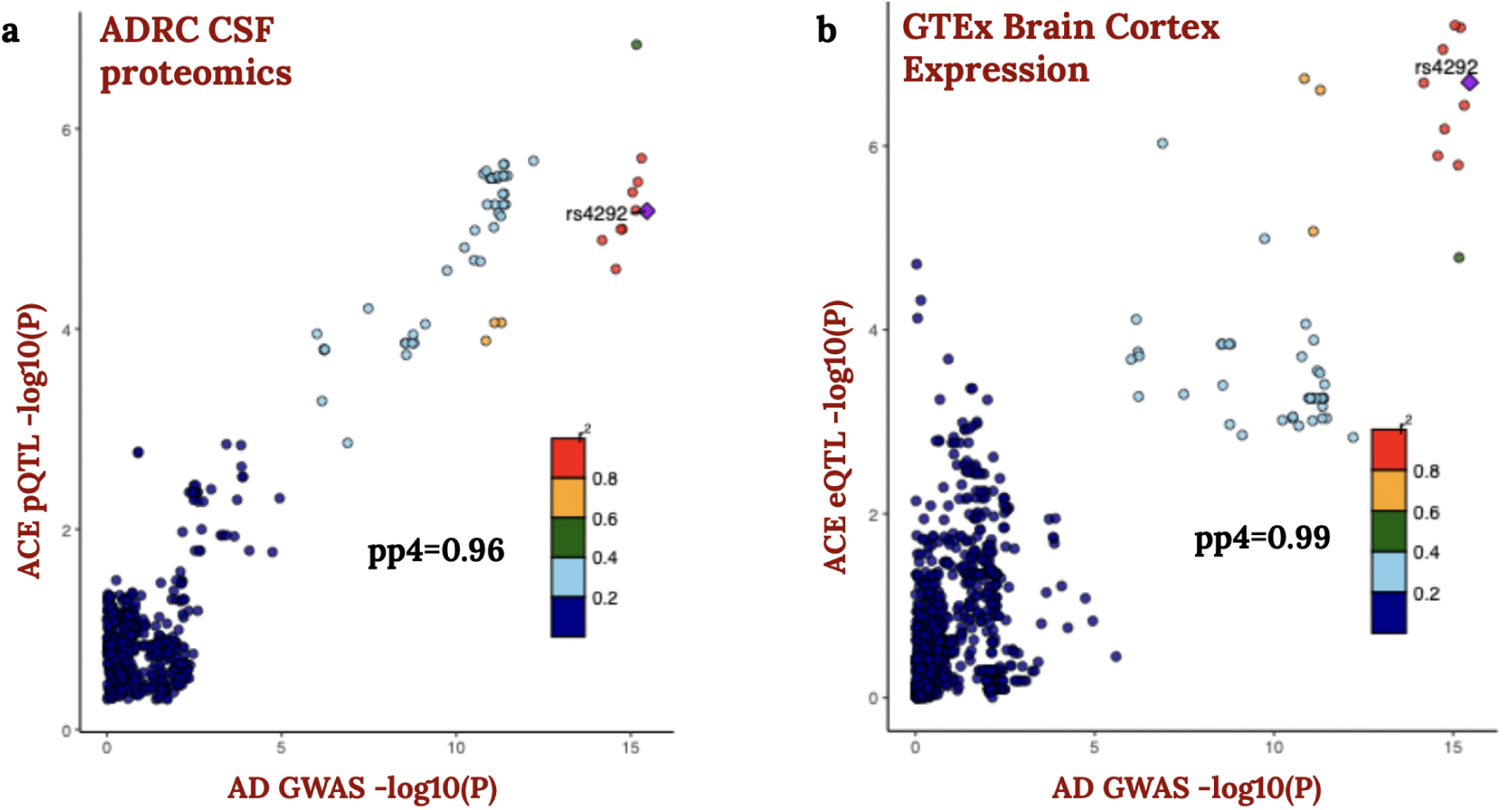
Colocalization of *ACE* molecular traits with Alzheimer’s disease (AD) GWAS in the ADRC and GTEx dataset. Colocalization plots between AD GWAS (Bellenguez et al., 2024, PMID: 35379992):**a,** *ACE* CSF protein levels in the ADRC cohort (pQTL); **b,** *ACE* expression in GTEx Brain Cortex (eQTL). Posterior probabilities (pp4) of colocalization are shown. Lead GWAS SNPs are labeled in the GTEx and GWAS SNPs intersection.

**Supplementary Figure 15.**
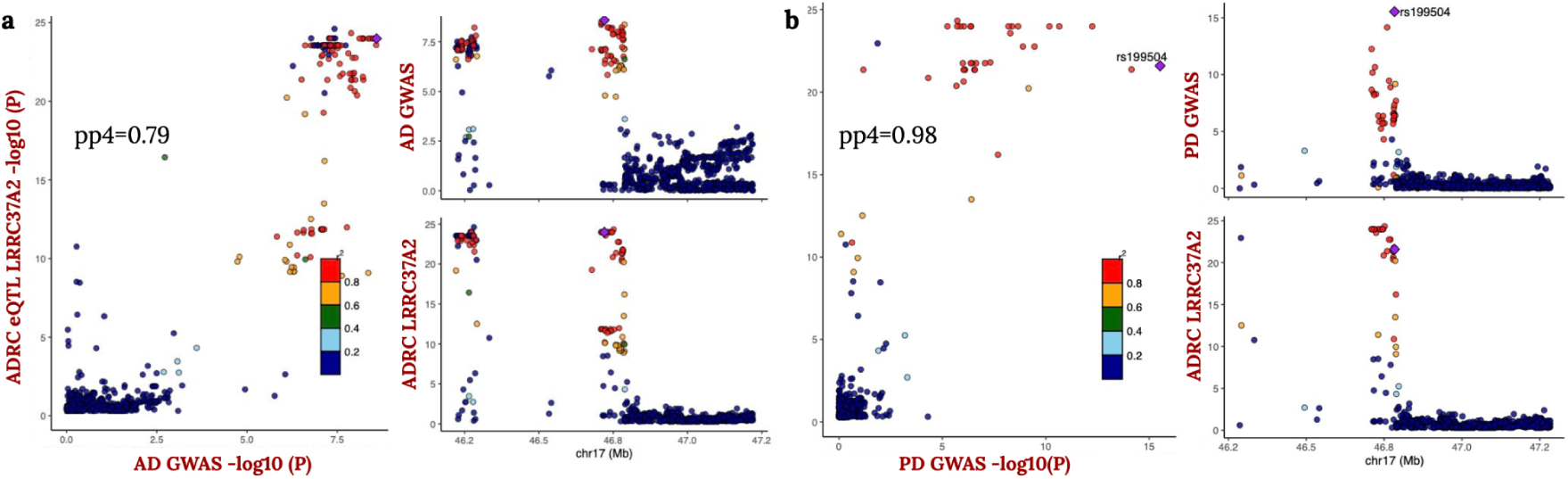
Colocalization of ADRC *LRRC37A2* eQTL with AD and PD GWAS signals. **a,** Colocalization between the *LRRC37A2* eQTL (ENSG00000238083) from the ADRC cohort and Alzheimer’s disease (AD) GWAS (Bellenguez et al., 2024). **b,** Colocalization between the same ADRC *LRRC37A2* eQTL (ENSG00000238083) and Parkinson’s disease (PD) GWAS (Kim et al., 2024).

**Supplementary Figure 16.**
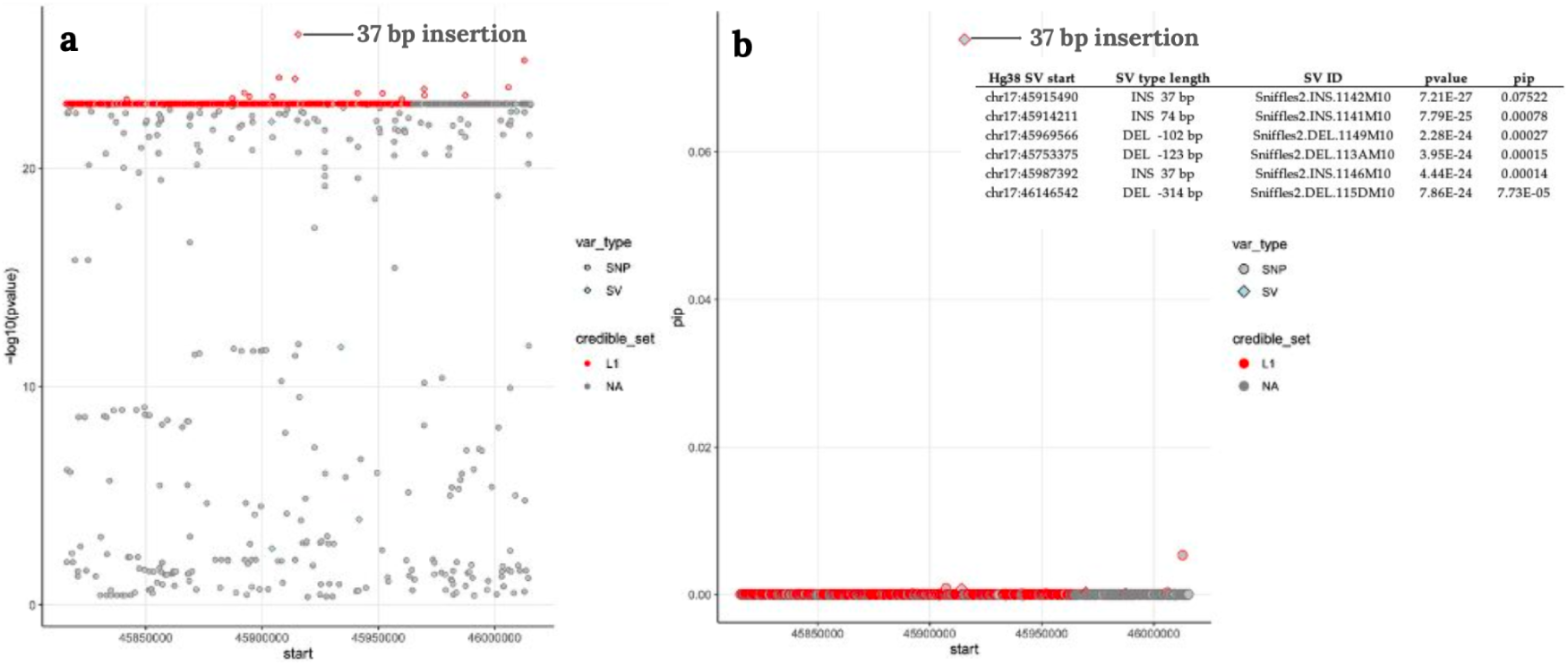
Fine-Mapping of eQTL for *LRRC37A2* in the ADRC Cohort. Example of fine-mapping of the eQTL for LRRC37A2 (ENSG00000238083) in the ADRC dataset, with the lead variant being a 37 bp insertion (Sniffles2.INS.1142M10, chr17:45915490). The L1 cluster includes variants within the credible set. Structural variants (SVs) are represented as diamonds, and single-nucleotide polymorphisms (SNPs) as circles. **a,** shows the –log10(p-value), and **b,**displays the posterior inclusion probability (PIP).

## References

Audano, Peter A., Arvis Sulovari, Tina A. Graves-Lindsay, Stuart Cantsilieris, Melanie Sorensen, Annemarie E. Welch, Max L. Dougherty, et al. 2019. “Characterizing the Major Structural Variant Alleles of the Human Genome.” Cell 176 (3): 663–75.e19.

Bai, Haimeng, Adam C. Naj, Penelope Benchek, Logan Dumitrescu, Timothy Hohman, Kara Hamilton-Nelson, Asha R. Kallianpur, et al. 2023. “A Haptoglobin (HP) Structural Variant Alters the Effect of APOE Alleles on Alzheimer’s Disease.” Alzheimer’s & Dementia: The Journal of the Alzheimer’s Association 19 (11): 4886–95.

Bellenguez, Céline, Fahri Küçükali, Iris E. Jansen, Luca Kleineidam, Sonia Moreno-Grau, Najaf Amin, Adam C. Naj, et al. 2022. “New Insights into the Genetic Etiology of Alzheimer’s Disease and Related Dementias.” Nature Genetics 54 (4): 412–36.

Billingsley, Kimberley J., Jinhui Ding, Pilar Alvarez Jerez, Anastasia Illarionova, Kristin Levine, Francis P. Grenn, Mary B. Makarious, et al. 2023. “Genome-Wide Analysis of Structural Variants in Parkinson Disease.” Annals of Neurology 93 (5): 1012–22.

Billingsley, Kimberley J., Melissa Meredith, Kensuke Daida, Pilar Alvarez Jerez, Shloka Negi, Laksh Malik, Rylee M. Genner, et al. 2024. “Long-Read Sequencing of Hundreds of Diverse Brains Provides Insight into the Impact of Structural Variation on Gene Expression and DNA Methylation.” bioRxivorg. 10.1101/2024.12.16.628723.

Boettger, Linda M., Rany M. Salem, Robert E. Handsaker, Gina M. Peloso, Sekar Kathiresan, Joel N. Hirschhorn, and Steven A. McCarroll. 2016. “Recurring Exon Deletions in the HP (haptoglobin) Gene Contribute to Lower Blood Cholesterol Levels.” Nature Genetics 48 (4): 359–66.

Bowles, Kathryn R., Derian A. Pugh, Yiyuan Liu, Tulsi Patel, Alan E. Renton, Sara Bandres-Ciga, Ziv Gan-Or, et al. 2022. “17q21.31 Sub-Haplotypes Underlying H1-Associated Risk for Parkinson’s Disease Are Associated with LRRC37A/2 Expression in Astrocytes.” Molecular Neurodegeneration 17 (1): 48.

Chaisson, Mark J. P., Ashley D. Sanders, Xuefang Zhao, Ankit Malhotra, David Porubsky, Tobias Rausch, Eugene J. Gardner, et al. 2019. “Multi-Platform Discovery of Haplotype-Resolved Structural Variation in Human Genomes.” Nature Communications 10 (1): 1784.

Chemparathy, Augustine, Yann Le Guen, Yi Zeng, John Gorzynski, Tanner D. Jensen, Chengran Yang, Nandita Kasireddy, et al. 2024. “A 3’UTR Insertion Is a Candidate Causal Variant at the TMEM106B Locus Associated with Increased Risk for FTLD-TDP.” Neurology. Genetics 10 (1): e200124.

Chen, Siwei, Laurent C. Francioli, Julia K. Goodrich, Ryan L. Collins, Masahiro Kanai, Qingbo Wang, Jessica Alföldi, et al. 2024. “A Genomic Mutational Constraint Map Using Variation in 76,156 Human Genomes.” Nature 625 (7993): 92–100.

Chiang, Colby, Alexandra J. Scott, Joe R. Davis, Emily K. Tsang, Xin Li, Yungil Kim, Tarik Hadzic, et al. 2017. “The Impact of Structural Variation on Human Gene Expression.” Nature Genetics 49 (5): 692–99.

Chui, Martin Man-Chun, Anna Ka-Yee Kwong, Hiu Yu Cherie Leung, Chingyiu Pang, Ines F. Scheller, Sheila Suet-Na Wong, Cheuk-Wing Fung, et al. 2025. “An Outlier Approach: Advancing Diagnosis of Neurological Diseases through Integrating Proteomics into Multi-Omics Guided Exome Reanalysis.” Npj Genomic Medicine 10 (1): 36.

Chung, Jaeyoon, Sandro Marini, Joanna Pera, Bo Norrving, Jordi Jimenez-Conde, Jaume Roquer, Israel Fernandez- Cadenas, et al. 2019. “Genome-Wide Association Study of Cerebral Small Vessel Disease Reveals Established and Novel Loci.” Brain: A Journal of Neurology 142 (10): 3176–89.

Collins, Ryan L., Harrison Brand, Konrad J. Karczewski, Xuefang Zhao, Jessica Alföldi, Laurent C. Francioli, Amit V. Khera, et al. 2020. “A Structural Variation Reference for Medical and Population Genetics.” Nature 581 (7809): 444–51.

Costa-Mallen, Paola, Cyrus P. Zabetian, Pinky Agarwal, Shu-Ching Hu, Dora Yearout, Ali Samii, James B. Leverenz, John W. Roberts, and Harvey Checkoway. 2015. “Haptoglobin Phenotype Modifies Serum Iron Levels and the Effect of Smoking on Parkinson Disease Risk.” Parkinsonism & Related Disorders 21 (9): 1087–92.

Cruchaga, Carlos, Dan Western, Jigyasha Timsina, Lihua Wang, Ciyang Wang, Chengran Yang, Muhammad Ali, et al. 2023. “Proteogenomic Analysis of Human Cerebrospinal Fluid Identifies Neurologically Relevant Regulation and Informs Causal Proteins for Alzheimer’s Disease.” Res. Sq. 10.21203/rs.3.rs-2814616/v1.

Dwarshuis, Nathan, Divya Kalra, Jennifer McDaniel, et al. 2024. “The GIAB Genomic Stratifications Resource for Human Reference Genomes.” Nature Communications 15 (1): 9029.

Ebert, Peter, Peter A. Audano, Qihui Zhu, Bernardo Rodriguez-Martin, David Porubsky, Marc Jan Bonder, Arvis Sulovari, et al. 2021. “Haplotype-Resolved Diverse Human Genomes and Integrated Analysis of Structural Variation.” Science (New York, N.Y.) 372 (6537): eabf7117.

Eichler, Evan E. 2019. “Genetic Variation, Comparative Genomics, and the Diagnosis of Disease.” The New England Journal of Medicine 381 (1): 64–74.

ENCODE Project Consortium. 2012. “An Integrated Encyclopedia of DNA Elements in the Human Genome.” Nature 489 (7414): 57–74.

English, Adam C., Vipin K. Menon, Richard A. Gibbs, Ginger A. Metcalf, and Fritz J. Sedlazeck. 2022. “Truvari: Refined Structural Variant Comparison Preserves Allelic Diversity.” Genome Biology 23 (1): 271.

Ferkingstad, Egil, Patrick Sulem, Bjarni A. Atlason, Gardar Sveinbjornsson, Magnus I. Magnusson, Edda L. Styrmisdottir, Kristbjorg Gunnarsdottir, et al. 2021. “Large-Scale Integration of the Plasma Proteome with Genetics and Disease.” Nature Genetics 53 (12): 1712–21.

Ferraro, Nicole M., Benjamin J. Strober, Jonah Einson, Nathan S. Abell, Francois Aguet, Alvaro N. Barbeira, Margot Brandt, et al. 2020. “Transcriptomic Signatures across Human Tissues Identify Functional Rare Genetic Variation.” Science (New York, N.Y.) 369 (6509). 10.1126/science.aaz5900.

Fulco, Charles P., Joseph Nasser, Thouis R. Jones, Glen Munson, Drew T. Bergman, Vidya Subramanian, Sharon R. Grossman, et al. 2019. “Activity-by-Contact Model of Enhancer-Promoter Regulation from Thousands of CRISPR Perturbations.” Nature Genetics 51 (12): 1664–69.

Gamaarachchi, Hasindu, Chun Wai Lam, Gihan Jayatilaka, Hiruna Samarakoon, Jared T. Simpson, Martin A. Smith, and Sri Parameswaran. 2020. “GPU Accelerated Adaptive Banded Event Alignment for Rapid Comparative Nanopore Signal Analysis.” BMC Bioinformatics 21 (1): 343.

Grandke, Friederike, Tobias Fehlmann, Fabian Kern, David M. Gate, Tobias William Wolff, Olivia Leventhal, Divya Channappa, et al. 2025. “A Single-Cell Atlas to Map Sex-Specific Gene-Expression Changes in Blood upon Neurodegeneration.” Nature Communications 16 (1): 1965.

GTEx Consortium. 2013. “The Genotype-Tissue Expression (GTEx) Project.” Nature Genetics 45 (6): 580–85.

Gustafson, Jonas A., Sophia B. Gibson, Nikhita Damaraju, Miranda P. G. Zalusky, Kendra Hoekzema, David Twesigomwe, Lei Yang, et al. 2024. “High-Coverage Nanopore Sequencing of Samples from the 1000 Genomes Project to Build a Comprehensive Catalog of Human Genetic Variation.” Genome Research 34 (11): 2061–73.

Huang, Jie, Jennifer E. Huffman, Yunfeng Huang, Ítalo Do Valle, Themistocles L. Assimes, Sridharan Raghavan, Benjamin F. Voight, et al. 2022. “Genomics and Phenomics of Body Mass Index Reveals a Complex Disease Network.” Nature Communications 13 (1): 7973.

Jensen, Tanner D., Bohan Ni, Chloe M. Retuer, John E. Gorzynski, et al. 2025 “Integration of transcriptomics and long-read genomics prioritizes structural variants in rare disease.” Genome research vol. 35,4 914–928

Jensen, Tanner D., Rhina Kaur, et al. “Population-scale detection of methylation outliers from long-read genome sequencing.” medRxiv [Preprint]. 2026 doi: 10.64898/2026.06.09.26355279.

Kagda, Meenakshi S., Bonita Lam, Casey Litton, et al. 2025. “Data Navigation on the ENCODE Portal.” Nature Communications 16 (1): 9592.

Kim, Jonggeol Jeffrey, Dan Vitale, Diego Véliz Otani, Michelle Mulan Lian, Karl Heilbron, 23andMe Research Team, Hirotaka Iwaki, et al. 2024. “Multi-Ancestry Genome-Wide Association Meta-Analysis of Parkinson’s Disease.” Nature Genetics 56 (1): 27–36.

Lei, Haixin, Ian N. M. Day, and Igor Vorechovský. 2005. “Exonization of AluYa5 in the Human ACE Gene Requires Mutations in Both 3’ and 5’ Splice Sites and Is Facilitated by a Conserved Splicing Enhancer.” Nucleic Acids Research 33 (12): 3897–3906.

Li, Taibo, Nicole Ferraro, Benjamin J. Strober, Francois Aguet, Silva Kasela, Marios Arvanitis, Bohan Ni, et al. 2023. “The Functional Impact of Rare Variation across the Regulatory Cascade.” Cell Genomics 3 (10): 100401.

Liang, Liang, Changchang Cao, Lei Ji, et al. 2023. “Complementary Alu Sequences Mediate Enhancer-Promoter Selectivity.” Nature 619 (7971): 868–875.

Liao, Wen-Wei, Mobin Asri, Jana Ebler, et al. 2023. “A Draft Human Pangenome Reference.” Nature 617 (7960): 312– 324.

Liu, Boxiang, Michael J. Gloudemans, Abhiram S. Rao, Erik Ingelsson, and Stephen B. Montgomery. 2019. “Abundant Associations with Gene Expression Complicate GWAS Follow-Up.” Nature Genetics 51 (5): 768–69.

Logsdon, Glennis A., Mitchell R. Vollger, and Evan E. Eichler. 2020. “Long-Read Human Genome Sequencing and Its Applications.” Nature Reviews. Genetics 21 (10): 597–614.

Logsdon, Glennis A., Peter Ebert, Peter A. Audano, et al. 2025. “Complex Genetic Variation in Nearly Complete Human Genomes.” Nature 644 (8076): 430–441.

Lun, Aaron T. L., Davis J. McCarthy, and John C. Marioni. 2016. “A Step-by-Step Workflow for Low-Level Analysis of Single-Cell RNA-Seq Data with Bioconductor.” F1000Research 5 (August):2122.

McCreight, Alexander, Yanghyeon Cho, Ruixi Li, et al. 2025. “SuSiE 2.0: Improved Methods and Implementations for Genetic Fine-Mapping and Phenotype Prediction.” In bioRxivorg. BioRxiv, December 25.

Mahmoud, M., Y. Huang, K. Garimella, P. A. Audano, W. Wan, N. Prasad, R. E. Handsaker, et al. 2024. “Utility of Long-Read Sequencing for All of Us.” Nature Communications 15 (1): 837.

Nalls, Mike A., Cornelis Blauwendraat, Costanza L. Vallerga, Karl Heilbron, Sara Bandres-Ciga, Diana Chang, Manuela Tan, et al. 2019. “Identification of Novel Risk Loci, Causal Insights, and Heritable Risk for Parkinson’s Disease: A Meta-Analysis of Genome-Wide Association Studies.” Lancet Neurology 18 (12): 1091–1102.

Nurk, Sergey, Sergey Koren, Arang Rhie, Mikko Rautiainen, Andrey V. Bzikadze, Alla Mikheenko, Mitchell R. Vollger, et al. 2022. “The Complete Sequence of a Human Genome.” Science (New York, N.Y.) 376 (6588): 44–53.

Oh, Hamilton Se-Hwee, Jarod Rutledge, Daniel Nachun, Róbert Pálovics, Olamide Abiose, Patricia Moran-Losada, Divya Channappa, et al. 2023. “Organ Aging Signatures in the Plasma Proteome Track Health and Disease.” Nature 624 (7990): 164–72.

Olson, Nathan D., Justin Wagner, Jennifer McDaniel, et al. 2022. “PrecisionFDA Truth Challenge V2: Calling Variants from Short and Long Reads in Difficult-to-Map Regions.” Cell Genomics 2 (5): 100129.

Rao, Suhas S. P., Miriam H. Huntley, Neva C. Durand, et al. 2014. “A 3D Map of the Human Genome at Kilobase Resolution Reveals Principles of Chromatin Looping.” Cell 159 (7): 1665–1680.

Rasooly, Danielle, Gina M. Peloso, and Claudia Giambartolomei. 2022. “Bayesian Genetic Colocalization Test of Two Traits Using Coloc.” Current Protocols 2 (12): e627.

Robinson, Mark D., Davis J. McCarthy, and Gordon K. Smyth. 2010. “edgeR: A Bioconductor Package for Differential Expression Analysis of Digital Gene Expression Data.” Bioinformatics (Oxford, England) 26 (1): 139–40.

Schloissnig, Siegfried, Samarendra Pani, Jana Ebler, et al. 2025. “Structural Variation in 1,019 Diverse Humans Based on Long-Read Sequencing.” Nature 644 (8076): 442–452.

Sheng, Jintao, Alexandra N. Trelle, America Romero, Jennifer Park, Tammy T. Tran, Sharon J. Sha, Katrin I. Andreasson, Edward N. Wilson, Elizabeth C. Mormino, and Anthony D. Wagner. 2025. “Top-down Attention and Alzheimer’s Pathology Affect Cortical Selectivity during Learning, Influencing Episodic Memory in Older Adults.” Science Advances 11 (24): eads4206.

Simpson, Jared T., Rachael E. Workman, P. C. Zuzarte, Matei David, L. J. Dursi, and Winston Timp. 2017. “Detecting DNA Cytosine Methylation Using Nanopore Sequencing.” Nature Methods 14 (4): 407–10.

Smit, AFA, Hubley, R & Green, P. 2015. RepeatMasker Open-4.0. http://www.repeatmasker.org

Sun, Benjamin B., Joshua Chiou, Matthew Traylor, Christian Benner, Yi-Hsiang Hsu, Tom G. Richardson, Praveen Surendran, et al. 2023. “Plasma Proteomic Associations with Genetics and Health in the UK Biobank.” Nature 622 (7982): 329–38.

Traylor, Matthew, Elodie Persyn, Liisa Tomppo, Sofia Klasson, Vida Abedi, Mark K. Bakker, Nuria Torres, et al. 2021. “Genetic Basis of Lacunar Stroke: A Pooled Analysis of Individual Patient Data and Genome-Wide Association Studies.” Lancet Neurology 20 (5): 351–61.

Trelle, Alexandra N., Valerie A. Carr, Edward N. Wilson, Michelle S. Swarovski, Madison P. Hunt, Tyler N. Toueg, Tammy T. Tran, et al. 2021. “Association of CSF Biomarkers with Hippocampal-Dependent Memory in Preclinical Alzheimer Disease.” Neurology 96 (10): e1470–81.

Vialle, Ricardo A., Katia de Paiva Lopes, Yan Li, Bernard Ng, Julie A. Schneider, Aron S. Buchman, Yanling Wang, et al. 2025. “Structural Variants Linked to Alzheimer’s Disease and Other Common Age-Related Clinical and Neuropathologic Traits.” Genome Medicine 17 (1): 20.

Wang, Gao, Abhishek Sarkar, Peter Carbonetto, and Matthew Stephens. 2020. “A Simple New Approach to Variable Selection in Regression, with Application to Genetic Fine Mapping.” Journal of the Royal Statistical Society. Series B, Statistical Methodology 82 (5): 1273–1300.

Wang, Hui, Timothy S. Chang, Beth A. Dombroski, Po-Liang Cheng, Vishakha Patil, Leopoldo Valiente-Banuet, Kurt Farrell, et al. 2024. “Whole-Genome Sequencing Analysis Reveals New Susceptibility Loci and Structural Variants Associated with Progressive Supranuclear Palsy.” Molecular Neurodegeneration 19 (1): 61.

Wang, Hui, Beth A. Dombroski, Po-Liang Cheng, Albert Tucci, Ya-Qin Si, John J. Farrell, Jung-Ying Tzeng, et al. 2025. “Structural Variation Detection and Association Analysis of Whole-Genome-Sequence Data from 16,543 Alzheimer’s Disease Sequencing Project Subjects.” Alzheimer’s & Dementia: The Journal of the Alzheimer’s Association 21 (6): e70277.

Western, Daniel, Jigyasha Timsina, Lihua Wang, Ciyang Wang, Chengran Yang, Bridget Phillips, Yueyao Wang, et al. 2024. “Proteogenomic Analysis of Human Cerebrospinal Fluid Identifies Neurologically Relevant Regulation and Implicates Causal Proteins for Alzheimer’s Disease.” Nature Genetics 56 (12): 2672–84.

Winer, Joseph R., Melanie J. Plastini, America Romero, Hillary Vossler, Isha Sai, Divya Channappa, Carla Abdelnour, et al. 2025. “Effects of α-Synuclein Pathology in Normal Aging and Alzheimer’s Disease.” medRxiv. 10.1101/2025.08.06.25333152.

Wu, Shyh-Jong, Tusty-Jiuan Hsieh, Mei-Chuan Kuo, Mei-Lan Tsai, Ke-Li Tsai, Chun-Hung Chen, and Yuan-Han Yang. 2013. “Functional Regulation of Alu Element of Human Angiotensin-Converting Enzyme Gene in Neuron Cells.” Neurobiology of Aging 34 (7): 1921.e1–7.

Xu, Xiaolu, Peng Zhang, Zhizheng Zhuo, Yunyun Duan, Liying Qu, Dan Cheng, Ting Sun, et al. 2024. “Prediction of H3K27M Alteration Status in Brainstem Glioma Using Multi-Shell Diffusion MRI Metrics.” Journal of Magnetic Resonance Imaging 60 (2): 576–85.

Zhan, Hui, Davis Cammann, Jeffrey L. Cummings, Xianjun Dong, and Jingchun Chen. 2025. “Biomarker Identification for Alzheimer’s Disease through Integration of Comprehensive Mendelian Randomization and Proteomics Data.” Journal of Translational Medicine 23 (1): 278.

